# Clinical evaluation of the Xpert MTB/XDR assay for rapid detection of isoniazid, fluoroquinolone, ethionamide and second-line drug resistance: A cross-sectional multicentre diagnostic accuracy study

**DOI:** 10.1101/2021.05.06.21256505

**Authors:** Adam Penn-Nicholson, Sophia B. Georghiou, Nelly Ciobanu, Mubin Kazi, Manpreet Bhalla, Anura David, Francesca Conradie, Morten Ruhwald, Valeriu Crudu, Camilla Rodrigues, Vithal Prasad Myneedu, Lesley Scott, Claudia M Denkinger, Samuel G Schumacher, Xpert XDR Trial Consortium

**Author notes:** **Corresponding author:** Adam Penn-Nicholson, Campus Biotech, 9 Chemin des Mines, 1202 Geneva, Switzerland. Authors contributed equally (Adam Penn-Nicholson, Sophia B. Georghiou). The members of the Xpert XDR Trial Consortium are listed at the end of this paper.

## Abstract

**Background:** The WHO End TB Strategy requires universal drug susceptibility testing and treatment of all people with tuberculosis. However, available second-line diagnostic tools are cumbersome and require sophisticated laboratory infrastructure, and ultimately less than half of those with drug-resistant tuberculosis receive appropriate treatment. Xpert MTB/XDR was developed to help overcome these limitations.

**Methods:** We assessed the diagnostic accuracy of sputum-based Xpert MTB/XDR for isoniazid, fluoroquinolone, ethionamide and second-line injectable resistance detection in adults with an Xpert MTB/RIF or Ultra *Mycobacterium tuberculosis*-positive result against a composite reference standard of phenotypic drug-susceptibility testing and whole genome sequencing (NCT03728725). Participants with pulmonary tuberculosis symptoms and ≥1 risk factor for drug resistance were consecutively enrolled between four clinical sites in India, Moldova and South Africa.

**Findings:** Between 31 July 2019 and 21 March 2020, we enrolled 710 patients, of which 611 (86.1%) had results from index and composite reference standard tests and were included in analysis. The sensitivity of Xpert MTB/XDR was 94% for isoniazid, 95% for fluoroquinolones, 54% for ethionamide, 73% for amikacin, 86% for kanamycin, and 61% for capreomycin resistance detection. Specificity was 98-100% for all drugs. Performance was equivalent to line-probe assays. The non-determinate rate of Xpert MTB/XDR was 2·96%.

**Interpretation:** This first prospective, multicentre clinical study of the Xpert MTB/XDR assay demonstrated high diagnostic test accuracy, meeting target product profile criteria for a next-generation drug susceptibility test.

**Funding:** German Federal Ministry of Education and Research through KfW, Dutch Ministry of Foreign Affairs, and Australian Department of Foreign Affairs and Trade.

**Research in context:** *Evidence before this study:* The World Health Organization (WHO) has highlighted the development of expanded, rapid molecular drug susceptibility tests as a key priority to tackle drug-resistant tuberculosis (TB). Prior to the Cepheid Xpert MTB/XDR assay, the only WHO-recommended rapid molecular assay for second-line resistance detection was the Bruker-Hain GenoType MTBDR*sl* line probe assay. However, the high complexity of DNA-based hybridization assays limits their use to central reference or regional-level laboratories where the appropriate infrastructure and user expertise can be ensured. In this context, the Xpert MTB/XDR assay is the only lower complexity, automated molecular assay for broader resistance detection suitable for use at lower levels of the laboratory network. We searched PubMed databases for articles evaluating the performance of the sputum-based assay using search terms (tuberculosis or TB) AND (Xpert OR GeneXpert OR cartridge) AND (second-line OR XDR OR “extensively drug-resistant”) AND (sputum OR sputa) AND (test OR assay OR diagnostic OR “point of care”) AND (performance OR accuracy OR sensitivity OR specificity OR diagnos*). The search was done on 8 Feb 2021 with no search date or language restrictions. Our search yielded 51 studies, of which only three reported on a sputum-based cartridge for expanded resistance detection. The first two studies described the assay design approach and reported initial performance metrics for a prototype version of the Xpert MTB/XDR assay. In the third study, the manufacturer demonstrated high performance for the final assay (sensitivity of 94–100% and a specificity of 100% for all drugs except for ethionamide) when compared with sequencing for 314 sputum specimens and sediments. This study was the first assessment of Xpert MTB/XDR assay diagnostic accuracy in a prospective patient cohort.

*Added value of this study:* This is the first clinical study of the diagnostic performance of the Xpert MTB/XDR assay for expanded drug resistance detection. This study was performed independent of the manufacturer in settings of intended use utilizing prospectively collected primary clinical samples. The study employed a comprehensive reference standard using both phenotypic drug susceptibility testing and whole genome sequencing separately, as well as in combination, which allowed a differentiated view of performance of the assay for resistance detection. Moreover, we compared the performance of the assay on direct sputum samples with the performance of the WHO-recommended Bruker-Hain MTBDR*plus* and MTBDR*sl* line probe assays on culture.

*Implications of all the available evidence:* The WHO End TB Strategy calls for early diagnosis and universal access to drug susceptibility testing. With a 10-color calibration to GeneXpert instruments, the Xpert MTB/XDR assay expands upon the drug detection landscape of the Xpert MTB/RIF and Ultra assays and overcomes the limitations of the Bruker-Hain line probe assays to offer an option for lower-level health care centres to conduct rapid, expanded drug susceptibility testing in follow-up to a TB-positive result using existing laboratory infrastructure. Data from this study demonstrates that the Xpert MTB/XDR assay has high performance for a diverse clinical population in the intended setting of use, providing national TB programmes with a valuable tool for expanded drug susceptibility testing for all persons with signs and symptoms of TB.

## Background

In 2010, the World Health Organization (WHO) recommended the use of the Xpert MTB/RIF® assay (“Xpert”, Cepheid, Sunnyvale, CA), an integrated, automated, cartridge-based system for multidrug-resistant tuberculosis (MDR-TB) diagnosis that uses the GeneXpert instrument platform.^1^ The assay and its more sensitive WHO-recommended successor, Xpert MTB/RIF Ultra® (“Ultra”), have since been widely adopted in tuberculosis (TB) programmes.^2^ Both assays identify *Mycobacterium tuberculosis* if present in the specimen. However, both assays detect rifampicin (RIF) resistance only, and thus do not provide sufficient information regarding a given strain’s resistance profile necessary to diagnose *M. tuberculosis* resistance to additional drug compounds, including the first-line drug isoniazid (INH), fluoroquinolones (FQ) and any of the injectable compounds [amikacin (AMK), kanamycin (KAN) and/or capreomycin (CAP)]. The rapid diagnosis and appropriate treatment of drug-resistant TB (DR-TB) is essential to prevent significant morbidity, mortality and further transmission of TB disease. Given the importance of INH and the FQs, in particular, in current TB and DR-TB patient treatment regimens,^3–6^ it is critical to rule-out resistance to these compounds prior to treatment.

The novel Xpert MTB/XDR assay is a rapid, sputum-based assay that is indicated for use as a reflex test to any *M. tuberculosis* positive result to test for resistance to INH, FQ, ethionamide (ETH), and second-line injectable drugs (SLI).^7^ A prototype version of the assay showed promising performance for INH and second-line resistance detection in a blinded study of 24 clinical sputum samples (sensitivity 75–100%; specificity 94–100%), as well as in a clinical evaluation study (sensitivity 92·7–98·1%; specificity 94·3–99·6%).^8,9^ The assay has since been updated with additional gene targets and the time-to-result shortened to 90 minutes. The final assay detects INH resistance-associated mutations in *katG*, the *inhA* promoter, *fabG1* and the *oxyR-ahpC* intergenic gene region, FQ resistance-associated mutations in *gyrA* and *gyrB*, ETH resistance-associated mutations in the *inhA* promoter, and SLI (AMK, KAN and CAP) resistance-associated mutations in *rrs* and the *eis* promoter. In a retrospective study of 314 clinical sputum samples and sediments in two laboratories, the final assay demonstrated high performance compared with sequencing (99·7%, 97·5%, 100%, 96·5%, 94·1% and 88·5% for detection of INH, FLQ, AMK, KAN, CAP and ETH resistance respectively, with a specificity of 100% for all the drugs except for ETH, for which the assay demonstrated a specificity of 97·3%).^10^

We conducted a cross-sectional, multicentre, diagnostic accuracy study in which the performance of the Xpert MTB/XDR assay on sputum samples was assessed in four reference laboratories in settings of intended use against Mycobacteria Growth Indicator Tube (MGIT) culture phenotypic drug susceptibility testing (pDST) and whole genome sequencing (WGS) as a composite reference standard for the diagnosis of INH, FQ, ETH, AMK, KAN and CAP resistance.

## Methods

### Study design and participants

This prospective, multicentre, cross-sectional diagnostic accuracy study (NCT03728725) was sponsored by FIND and conducted with the Phthisiopneumology Institute “Chiril Draganiuc”, in Chisinau, Moldova, P.D. Hinduja Hospital and Medical Research Centre in Mumbai, India, the National Institute of TB and Respiratory Diseases (NITRD) in New Delhi, India, and University of the Witwatersrand in Johannesburg, South Africa. The study was conducted in accordance with the 1964 Helsinki declaration and later amendments and approved by the relevant institutional review boards and independent ethics committees. All participants provided informed consent.

Individuals were recruited at outpatient clinic settings and inpatient hospital settings. Interested individuals were referred to study personnel for additional information and screening. Screening criteria included age 18 years or above, symptoms suggesting pulmonary tuberculosis and at least one risk factor for DR-TB. Participants were consecutively enrolled in the study if they met inclusion criteria, provided informed consent, had a *M. tuberculosis*-positive Xpert or Ultra result with either a RIF-resistant or -sensitive result, and provided at least 3 mL of sputum. Information regarding patient sex, age, weight, height, infection site, previous TB treatment start date and drug regimen, pulmonary symptoms, and HIV status were collected at enrolment. Clinical information and reference test result were not available to operators or readers of the index test at the time of data generation. All data entry was validated with source document verification. Xpert MTB/XDR test results were not shared with clinical staff and did not influence patient treatment options.

The full study protocol is provided as Supplement S1, and at clinicaltrials.gov (NCT03728725).

### Procedures

Patients meeting eligibility criteria were asked to provide ≥3 mL of sputum as either a single sputum or two consecutively collected and pooled sputa. Samples were homogenized with glass beads and split for testing. Xpert MTB/XDR and acid-fast bacilli smear were performed directly on the homogenized sputum sample. Liquid (MGIT) and solid (LJ) culture were performed using 2 mL of decontaminated sputum. MGIT DST was performed for all culture-positive samples for INH, RIF, FQs (moxifloxacin and levofloxacin), ETH, AMK, KAN, and CAP at the WHO-recommended critical concentrations for MGIT DST.^11,12^ Cultured samples underwent subsequent testing by Bruker-Hain GenoType MTBDR*plus*, MTBDR*sl* LPA, and WGS to report high-confidence resistance mutations in relevant gene regions (*katG, inhA, fabG1, ahpC, gyrA, gyrB, rrs, eis*, and *tlyA*),^13^ as well as a second Xpert MTB/XDR assay. DNA for WGS was extracted on site from 300 µL positive MGIT culture using Molzym Ultra-Deep Microbiome Prep kits and extracts were sent to MedGenome (Bangalore, India) for WGS (Supplement S2).

### Statistical analysis

We estimated that 760 total samples would need to be tested for this study to yield 600 specimens with full results (accounting for around 20% of culture and molecular tests with invalid and indeterminate results). We estimated that the analysis of 600 specimens would allow us to assess INH, FQ, AMK, KAN, and CAP sensitivity with 95% confidence intervals (based on the Wilson score method), of 7%, 13%, 23%, 16%, and 23%, respectively, and specificity with 95% confidence intervals of 3–4%, based upon knowledge of past drug resistance rates for the given sites or regions.^14^ More detail on the sample size calculations is provided in the protocol (Supplement S1).

Primary analyses focused on estimating accuracy (clinical sensitivity and specificity) for INH, ETH, FQ (moxifloxacin or levofloxacin), AMK, KAN, and CAP resistance detection using a composite reference standard. Specimens were labelled as drug-resistant if either pDST or WGS suggested drug-resistance; specimens were labelled as drug-sensitive if both pDST and WGS suggested drug-sensitivity. For each drug, clinical sensitivity was defined as the proportion resistant based on the composite reference standard that tested resistant by Xpert MTB/XDR. Clinical specificity was defined as the proportion susceptible based on the composite reference standard that tested susceptible by Xpert MTB/XDR. For simple proportions (sensitivity and specificity), 95% confidence intervals were computed using the Wilson score method. Primary diagnostic accuracy analyses were carried out across all study sites. All TB samples that generated both Xpert MTB/XDR (resistant or susceptible) and composite reference standard results contributed to the analysis of the diagnostic test accuracy for INH, FQ, ETH, and SLI resistance detection. Non-determinate rates of the Xpert MTB/XDR assay, defined as invalid *M. tuberculosis* detection, as well as indeterminate rates, defined as indeterminate results for drug resistance among valid *M. tuberculosis* detection results, were also calculated across study sites as a percentage of all Xpert MTB/XDR tests performed on patient sputa.

Secondary analyses investigated assay diagnostic accuracy between important subgroups, including: Xpert MTB/XDR performance on direct sputum (“on sputum”) versus cultured isolates (“on isolates”), performance by smear result, performance between clinical sites, performance by patient HIV status and TB pre-treatment status, and performance compared with the Bruker-Hain MTBDR*plus* and MTBDR*sl* assays. Detailed definitions for diagnostic test results are given in Supplement S3.

## Results

Between 31 July 2019 and 21 March 2020, 714 patients were screened for eligibility (Figure 1). A total of 710 patients were enrolled, of whom 99 were excluded, mainly as a result of negative cultures for *M. tuberculosis* (n=89) and the remaining 10 for missing or invalid reference or index test results. The initial population of 710 participants had a smear positivity rate of 69% and a culture positivity rate of 87%. The final, analysed population of 611 participants, for which results from both Xpert MTB/XDR and reference standard were available, had a median age of 37 years (18–77). Of these participants, 35% were women and 16% were HIV-positive (Figure 1, Table 1); 81% were Xpert MTB/RIF or Ultra RIF-resistant, while 76% were smear-positive for *M. tuberculosis*.

**Table 1.**
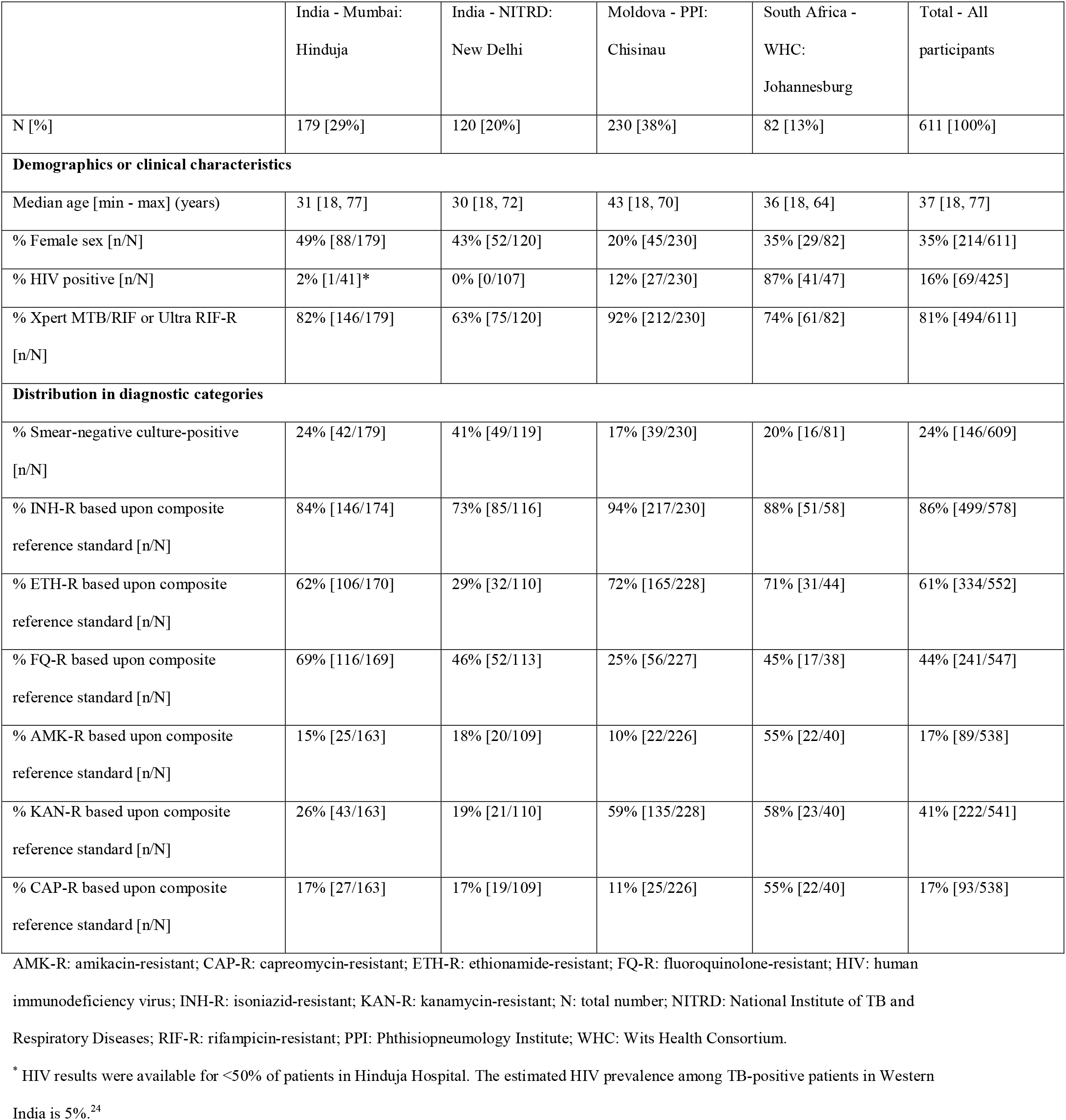
Demographics and clinical characteristics of analysed patient cohort.

|  | India - Mumbai:<br>Hinduja | India - NITRD:<br>New Delhi | Moldova - PPI:<br>Chisinau | South Africa -<br>WHC:<br>Johannesburg | Total - All<br>participants |
| --- | --- | --- | --- | --- | --- |
| N [%] | 179 [29%] | 120 [20%] | 230 [38%] | 82 [13%] | 611 [100%] |
| **Demographics or clinical characteristics** |  |  |  |  |  |
| Median age [min - max] (years) | 31 [18, 77] | 30 [18, 72] | 43 [18, 70] | 36 [18, 64] | 37 [18, 77] |
| % Female sex [n/N] | 49% [88/179] | 43% [52/120] | 20% [45/230] | 35% [29/82] | 35% [214/611] |
| % HIV positive [n/N] | 2% [1/41]* | 0% [0/107] | 12% [27/230] | 87% [41/47] | 16% [69/425] |
| % Xpert MTB/RIF or Ultra RIF-R<br>[n/N] | 82% [146/179] | 63% [75/120] | 92% [212/230] | 74% [61/82] | 81% [494/611] |
| **Distribution in diagnostic categories** |  |  |  |  |  |
| % Smear-negative culture-positive<br>[n/N] | 24% [42/179] | 41% [49/119] | 17% [39/230] | 20% [16/81] | 24% [146/609] |
| % INH-R based upon composite<br>reference standard [n/N] | 84% [146/174] | 73% [85/116] | 94% [217/230] | 88% [51/58] | 86% [499/578] |
| % ETH-R based upon composite<br>reference standard [n/N] | 62% [106/170] | 29% [32/110] | 72% [165/228] | 71% [31/44] | 61% [334/552] |
| % FQ-R based upon composite<br>reference standard [n/N] | 69% [116/169] | 46% [52/113] | 25% [56/227] | 45% [17/38] | 44% [241/547] |
| % AMK-R based upon composite<br>reference standard [n/N] | 15% [25/163] | 18% [20/109] | 10% [22/226] | 55% [22/40] | 17% [89/538] |
| % KAN-R based upon composite<br>reference standard [n/N] | 26% [43/163] | 19% [21/110] | 59% [135/228] | 58% [23/40] | 41% [222/541] |
| % CAP-R based upon composite<br>reference standard [n/N] | 17% [27/163] | 17% [19/109] | 11% [25/226] | 55% [22/40] | 17% [93/538] |

AMK-R: amikacin-resistant; CAP-R: capreomycin-resistant; ETH-R: ethionamide-resistant; FQ-R: fluoroquinolone-resistant; HIV: human
immunodeficiency virus; INH-R: isoniazid-resistant; KAN-R: kanamycin-resistant; N: total number; NITRD: National Institute of TB and
Respiratory Diseases; RIF-R: rifampicin-resistant; PPI: Phthisiopneumology Institute; WHC: Wits Health Consortium.
\* HIV results were available for <50% of patients in Hinduja Hospital. The estimated HIV prevalence among TB-positive patients in Western
India is 5%.<sup>24</sup>

**Figure 1.**
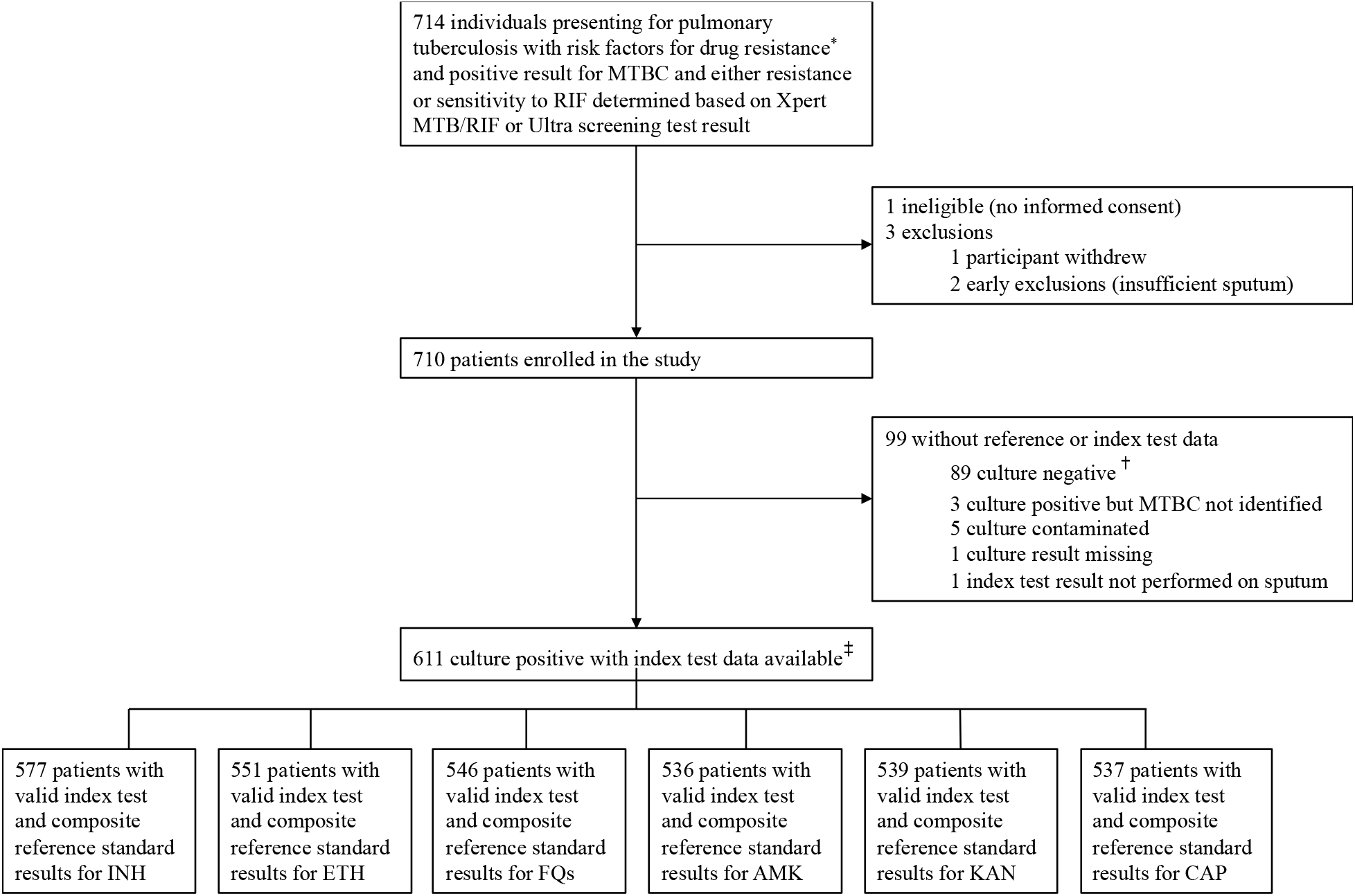
Participant enrolment and exclusions. AMK: amikacin; CAP: capreomycin; ETH: ethionamide; FQ: fluoroquinolones; INH: isoniazid; KAN: kanamycin; MTBC: *Mycobacterium tuberculosis* complex. ^*^ Drug-resistant tuberculosis risk factors included: previously received >1 month of treatment for a prior tuberculosis episode (n=286); failing tuberculosis treatment as demonstrated by a positive sputum smear or culture after ≥3 months of standard tuberculosis treatment (n=134); close contact with a known DR-TB case (n=59); newly diagnosed with MDR-TB within the last 30 days (n=305); or previously diagnosed with MDR-TB and failed tuberculosis treatment as demonstrated by a positive sputum smear or culture after ≥3 months of a standard MDR-TB treatment regimen (n=103). Many patients reported more than one risk factor for drug-resistant tuberculosis. ^†^ Culture-negative test results were observed in this study as the screening test (Xpert MTB/RIF or Ultra) was performed prior to enrolment on a different sputum sample than the sputum sample that was used for direct Xpert MTB/XDR and culture. ^‡^ The sum for individual drugs does not correspond to 611 as some samples had no reference standard (phenotypic drug susceptibility testing and/or whole genome sequencing) result for certain drugs.

The Xpert MTB/XDR assay had high diagnostic accuracy for the detection of INH, FQ, AMK, KAN, and CAP resistance in the clinical study (Figure 2). For Xpert MTB/XDR performed directly on sputum, sensitivity and specificity were 94% and 100% for INH resistance detection, 94% and 99% for FQ resistance detection, 54% and 100% for ETH resistance detection, 73% and 100% for AMK resistance detection, 86% and 98% for KAN resistance detection, and 61% and 100% for CAP resistance detection compared with a composite reference standard of pDST and WGS.

**Figure 2.**
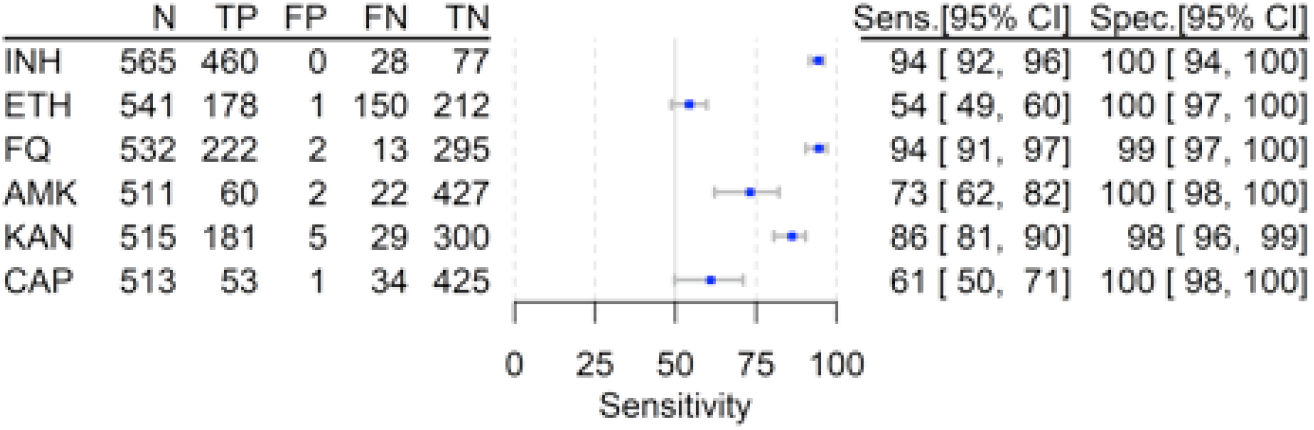
Sensitivity and specificity of the sputum-based Xpert MTB/XDR assay for resistance detection compared with a composite reference standard of phenotypic DST and WGS. AMK: amikacin; CAP: capreomycin; ETH: ethionamide; FN: false negatives; FP: false positives; F : fluoroquinolones; INH: isoniazid; KAN: kanamycin; Sens: sensitivity; Spec: specificity; TN: tru negatives; TP, true positives.

There was no notable difference of Xpert MTB/XDR performance when using sputum compared with isolates for the assay sample (Supplement S3). Xpert MTB/XDR invalid rate (i.e. unsuccessful test) was notably higher for smear negative samples than for smear positive samples (6·2% compared with 0·2%), though we detected no notable differences in performance for drug resistance detection based upon smear status (S3). Additionally, there were no performance differences by patient HIV or pre-treatment status (S3).

There were a few notable Xpert MTB/XDR performance differences by site (Figure 3). The sensitivity of the assay for INH resistance detection was lower in New Delhi (80%; 95% CI 70– 88%) than other sites, with WGS revealing the presence of INH resistance mutations that are not included in the Xpert MTB/XDR assay (e.g. *katG* W135^*^ stop codon), though specificity was high (100%) across all sites. Likewise, the sensitivity of the assay for ETH resistance detection was low in New Delhi and Mumbai compared with Moldova and South Africa, though specificity was high across and between sites. For SLI resistance detection, the Xpert MTB/XDR assay sensitivity was also notably lower in New Delhi (26–33%) than for other sites. The sensitivity of the assay for CAP resistance detection was also notably low in Moldova (40%). However, it should be noted that the sensitivity estimates for Xpert MTB/XDR detection of SLI resistance detection in New Delhi and CAP resistance detection in Moldova greatly improved if only pDST was used as a reference standard (67–75% and 56%, respectively, see Supplement S3).

**Figure 3.**
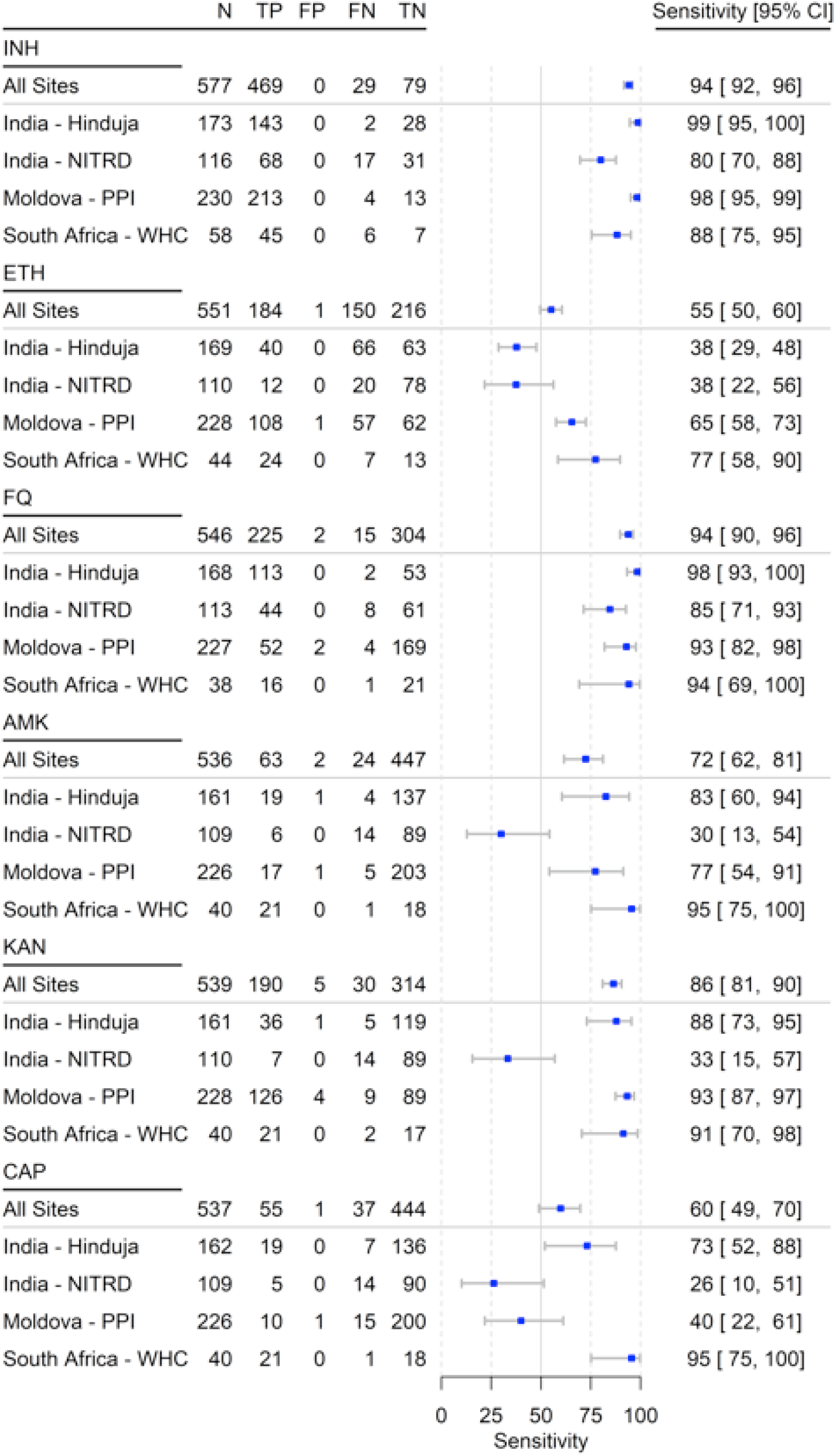
Sensitivity of the Xpert MTB/XDR assay for resistance detection by clinical site, compared with a composite reference standard of phenotypic DST and WGS. AMK: amikacin; CAP: capreomycin; ETH: ethionamide; FN: false negatives; FP: false positives; FQ: fluoroquinolones; INH: isoniazid; KAN: kanamycin; NITRD: National Institute of TB and Respiratory Diseases; PPI: Phthisiopneumology Institute; TN: true negatives; TP, true positives; WHC: Wits Health Consortium. Note: Specificity was above 98% for all drugs and all sites. Note: a table of Xpert MTB/XDR and reference standard discordant results is available in Supplement S3.

The overall non-determinate rate for the Xpert MTB/XDR assay was 2.96% (Table 2). Drug resistance indeterminate rates, among valid Xpert MTB/XDR *M. tuberculosis*-detected results, were ≤3.5% for all drugs included in the assay.

**Table 2.**
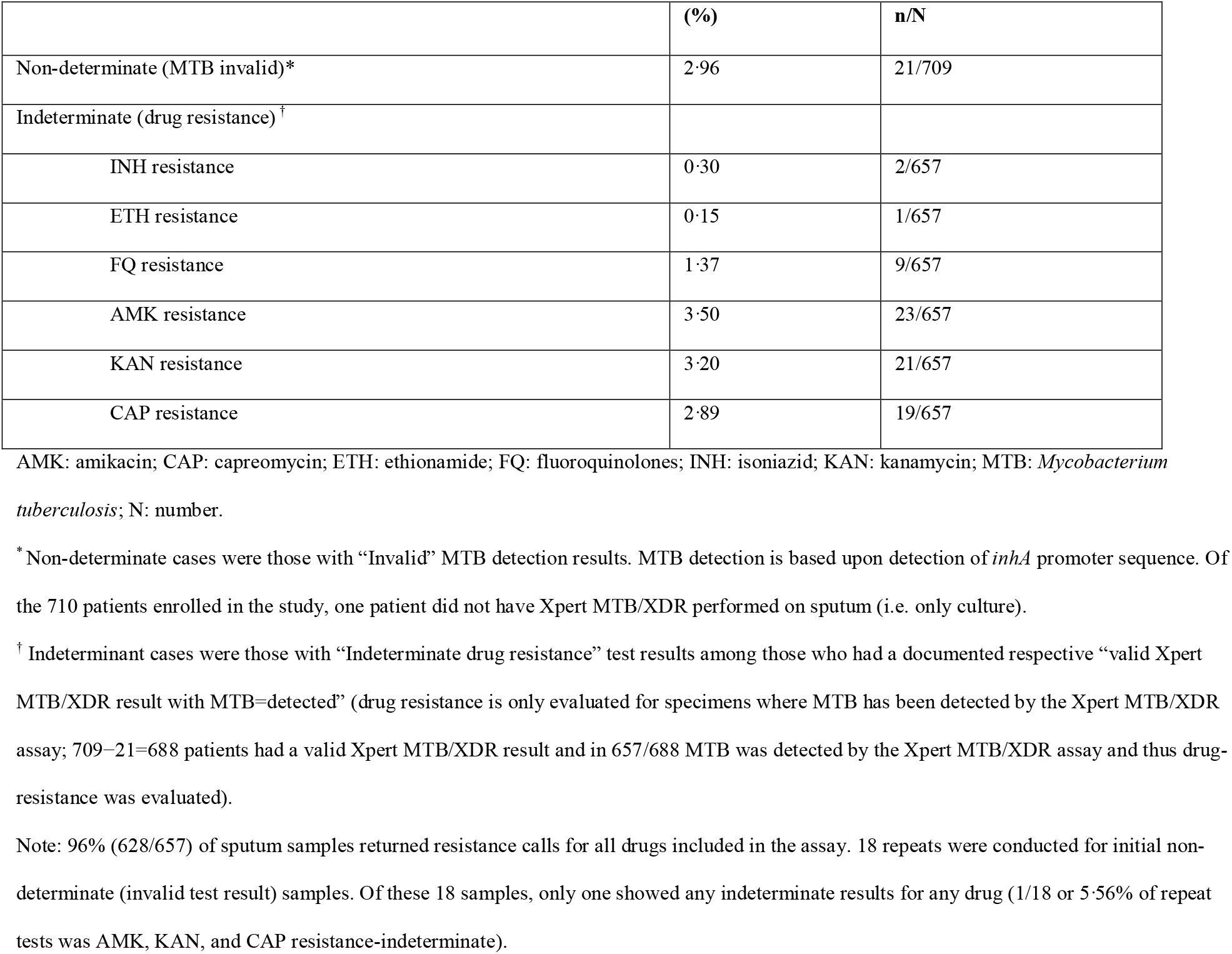
Overall Xpert MTB/XDR non-determinate indeterminate rates from testing of unprocessed sputum.

|  | (%) | n/N |
| --- | --- | --- |
| Non-determinate (MTB invalid)* | 2.96 | 21/709 |
| Indeterminate (drug resistance)<sup>†</sup> |  |  |
| INH resistance | 0.30 | 2/657 |
| ETH resistance | 0.15 | 1/657 |
| FQ resistance | 1.37 | 9/657 |
| AMK resistance | 3.50 | 23/657 |
| KAN resistance | 3.20 | 21/657 |
| CAP resistance | 2.89 | 19/657 |

AMK: amikacin; CAP: capreomycin; ETH: ethionamide; FQ: fluoroquinolones; INH: isoniazid; KAN: kanamycin; MTB: *Mycobacterium tuberculosis*; N: number.
\*Non-determinate cases were those with “Invalid” MTB detection results. MTB detection is based upon detection of *inhA* promoter sequence. Of the 710 patients enrolled in the study, one patient did not have Xpert MTB/XDR performed on sputum (i.e. only culture).
<sup>†</sup> Indeterminant cases were those with “Indeterminate drug resistance” test results among those who had a documented respective “valid Xpert MTB/XDR result with MTB=detected” (drug resistance is only evaluated for specimens where MTB has been detected by the Xpert MTB/XDR assay; 709–21=688 patients had a valid Xpert MTB/XDR result and in 657/688 MTB was detected by the Xpert MTB/XDR assay and thus drug-resistance was evaluated).
Note: 96% (628/657) of sputum samples returned resistance calls for all drugs included in the assay. 18 repeats were conducted for initial non-determinate (invalid test result) samples. Of these 18 samples, only one showed any indeterminate results for any drug (1/18 or 5.56% of repeat tests was AMK, KAN, and CAP resistance-indeterminate).

The Xpert MTB/XDR assay performed similarly to the Bruker-Hain line probe assays (LPAs) for INH, FQ, AMK, KAN, and CAP resistance detection, even though the Xpert MTB/XDR assay was performed directly on sputum samples while the LPAs were performed only on cultured samples (Table 3). The only difference was for INH resistance detection, as the Xpert MTB/XDR assay demonstrated slightly higher sensitivity, likely attributed to the two additional gene targets in Xpert MTB/XDR that are not found in the MTBDR*plus* assay (i.e. *fabG1* and the *ahpC-oxyR* intergenic region). Of the eight samples that were Xpert MTB/XDR INH-resistant, but MTBDR*plus* INH-susceptible in this study, WGS confirmed that two had *ahpC-oxyR* mutations (i.e. c-10t and c-15t) and three had the *fabG1* L203L mutation in the absence of other mutations. Two of the 8 samples did not have WGS available, though WGS confirmed that the 8th sample had a katG S315T mutation that was not detected by the LPA.

**Table 3.** Xpert MTB/XDR performance on sputum compared with MTBDR*plus* and MTBDR*sl* from culture isolates.

|  | N* | TP | FP | FN | TN | Sensitivity % (95% CI) | Specificity % (95% CI) |
| --- | --- | --- | --- | --- | --- | --- | --- |
| **<u>INH-R detection</u>** |  |  |  |  |  |  |  |
| MTBDRplus | 575 | 461 | 0 | 36 | 78 | 93 (90 to 95) | 100 (94 to 100) |
| Xpert MTB/XDR | 575 | 469 | 0 | 28 | 78 | 94 (92 to 96) | 100 (94 to 100) |
| Diff. [Xpert MTB/XDR - MTBDRplus] | 575 |  |  |  |  | +1.6 (+0.2, +3.4) | 0.0 (−4.7, +4.7) |
| **<u>FQ-R detection</u>** |  |  |  |  |  |  |  |
| MTBDRsl | 532 | 222 | 2 | 13 | 295 | 95 (91, 97) | 99 (97, 100) |
| Xpert MTB/XDR | 532 | 222 | 2 | 13 | 295 | 95 (91, 97) | 99 (97, 100) |
| Diff. [Xpert MTB/XDR - MTBDRplus] | 532 |  |  |  |  | 0.0 (−1.6, +1.6) | 0.0 (-1.3, +1.3) |
| **<u>AMK-R detection</u>** |  |  |  |  |  |  |  |
| MTBDRsl | 511 | 60 | 2 | 22 | 427 | 73 (62 to 82) | 100 (98 to 100) |
| Xpert MTB/XDR | 511 | 60 | 2 | 22 | 427 | 73 (62 to 82) | 100 (98 to 100) |
| Diff. [Xpert MTB/XDR - MTBDRplus] | 511 |  |  |  |  | 0.0 (−4.5, +4.5) | 0.0 (-0.9, +0.9) |
| **<u>KAN-R detection</u>** |  |  |  |  |  |  |  |
| MTBDRsl | 515 | 181 | 5 | 29 | 300 | 86 (81 to 90) | 98 (96 to 99) |
| Xpert MTB/XDR | 515 | 181 | 5 | 29 | 300 | 86 (81 to 90) | 98 (96 to 99) |
| Diff. [Xpert MTB/XDR - MTBDRplus] | 515 |  |  |  |  | 0.0 (−1.8, +1.8) | 0.0 (−1.2, +1.2) |
| **<u>CAP-R detection</u>** |  |  |  |  |  |  |  |
| MTBDRsl | 513 | 53 | 1 | 34 | 425 | 61 (50 to 71) | 100 (99 to 100) |
| Xpert MTB/XDR | 513 | 53 | 1 | 34 | 425 | 61 (50 to 71) | 100 (99 to 100) |
| Diff. [Xpert MTB/XDR - MTBDRplus] | 513 |  |  |  |  | 0.0 (−4.2, +4.2) | 0.0 (−0.9, +0.9) |

AMK-R: amikacin-resistant; CAP-R: capreomycin-resistant; CI: confidence interval; diff: difference; ETH-R: ethionamide-resistant; FN: false
negatives; FP: false positives; FQ-R: fluoroquinolone-resistant; INH-R: isoniazid-resistant; KAN-R: kanamycin-resistant; N: total number; TN:
true negatives; TP: true positives.
Presented data for line probe assays is from culture isolates of matched patients.
\*Analysis is limited to patients with results on both Xpert MTB/XDR and the respective Hain line probe assay to provide a direct head-to-head comparison. An analysis comparing performance when testing for both Xpert MTB/XDR and LPA was done from culture isolates is available in Supplement S3.

## Discussion

This is the first prospective, clinical diagnostic accuracy study of the Xpert MTB/XDR assay for INH, FQ, ETH, and second-line TB drug resistance detection. We found that the Xpert MTB/XDR assay, performed directly on smear-positive and smear-negative sputum specimens, demonstrated high diagnostic accuracy for the detection of INH, FQ, AMK, KAN, and CAP resistance among patients from the three WHO regions of Africa, Europe, and South-East Asia. Importantly, direct Xpert MTB/XDR testing of sputum samples produced results for resistance on all drugs in 96% of tested *M. tuberculosis*-positive specimens with a single, easy-to-use assay. The assay demonstrated slightly better performance for INH resistance detection compared with the Bruker-Hain MTBDR*plus* assay (1·6%, 95% CI 0·2-3·4), as the assay includes two additional gene targets not found in the LPA (i.e. *fabG1* and the *ahpC-oxyR* intergenic region), and equivalent performance for second-line resistance detection compared with the Bruker-Hain MTBDR*sl* assay.

Notably, specificity was high (>98%) for the detection of all drugs included in the Xpert MTB/XDR assay. Sensitivity of the assay varied by drug target, ranging from 54% for ETH resistance detection to 94% for FQ resistance detection against the composite reference standard. For INH, FQ, and SLI resistance detection, performance estimates met minimal criteria of the WHO target product profile for a next-generation DST assay to be used at peripheral microscopy centres compared with pDST (no criteria have been defined to date for ETH or for a composite reference standard).^15^ Furthermore, no performance differences were seen by smear status or patient HIV or pre-treatment status, highlighting the suitability of the test for a diverse patient population.

Some notable diagnostic performance variations were identified by site. In particular, assay sensitivity for INH resistance detection was lower in New Delhi (80%, 95% CI 70–88%) than for other sites. Most of the Xpert MTB/XDR INH false-negative samples in New Delhi were phenotypically INH-resistant with no high confidence resistance mutation(s) in the gene regions covered by the Xpert MTB/XDR assay. For example, three phenotypically INH-resistant samples from NITRD had *ahpC* t-34a/g mutations that are outside of the coverage of the Xpert MTB/XDR assay. Additionally, two different Indian isolates had stop codons in *katG* (W135^*^ and E607^*^) that were not identified by the Xpert MTB/XDR assay. Importantly, the MTBDR*plus* LPA also did not identify INH resistance for any of these samples, further suggesting that INH resistance mechanisms outside of those covered by the Xpert MTB/XDR assay and most other current molecular assays might play a role in resistance in this geographical context. Assay sensitivity for ETH resistance detection, in contrast, was low both overall and between sites, likely due to the fact that the Xpert MTB/XDR assay is only capable of detecting ETH resistance-associated mutations in one gene region (i.e. the *inhA* promoter), and so other phenotypic ETH resistance mechanisms, such as *ethA* resistance mutations,^16^ were missed by Xpert MTB/XDR, since they are not targeted by the assay. As no graded list of *ethA* resistance mutations currently exists for ETH, and ETH pDST is unreliable,^17^ we are unable to comment upon potential improvements in diagnostic performance if *ethA* resistance mutations were included in the assay. The low sensitivity for ETH suggests a role of the assay for ruling-in but not for ruling-out resistance to ETH.

For SLI resistance detection, Xpert MTB/XDR sensitivity was notably lower in New Delhi (26– 33%, depending on the drug) than for the other sites. Upon closer examination, it was determined that 11 of the 14 false-negative results in New Delhi were from specimens that were phenotypically SLI-susceptible but heteroresistant by WGS, with *rrs* c1402a and g1484t mutations present in only a fraction of WGS reads at levels likely undetectable by the Xpert MTB/XDR assay (the *rrs* c1402a mutation was detected in <50% of reads for nine of the 11 samples).^10,18^ Ultimately, the clinical relevance of these minor resistant populations is unclear given that these samples were phenotypically SLI-susceptible. If only pDST was considered as a reference standard in this study, the sensitivity of the Xpert MTB/XDR assay would improve to 67–75% for the detection of SLI resistance in New Delhi and 75–92% overall. The sensitivity of the assay for CAP resistance detection was also low in Moldova (40%, 95% CI 22–61%), likely due to the fact that certain CAP resistance mutations were present in only a low fraction of generated WGS reads for Xpert MTB/XDR false-negative samples (e.g. the *rrs* c1402a mutation was detected in ≤25% of reads for the five CAP-resistant samples that were not detected by Xpert MTB/XDR). Furthermore, a few additional phenotypically CAP-resistant samples did not have any resistance mechanisms detected by WGS, including *tlyA* mutations, suggesting that additional resistance mechanisms outside of those covered by the Xpert MTB/XDR assay might play a role in CAP resistance in Moldova.^19^

The strengths of this study included the enrolment of patients in four diverse sites in three WHO regions, the use of WHO-recommended molecular comparators (Bruker-Hain GenoType MTBDR*plus* and MTBDR*sl*), and the use of a composite reference standard to fully characterize assay performance. To investigate the performance of Xpert MTB/XDR in real-world populations, an effort was made to include diverse clinical populations across different geographical regions widely representative of the TB epidemic. However, differences in local patient populations, laboratories, and *M. tuberculosis* strains may have also contributed to performance variations seen between sites and reflect a limitation of studies including diverse populations. It is important to note that the study did not allow for evaluation of Xpert MTB/XDR assay performance for TB detection. As the study was designed to assess the Xpert MTB/XDR assay as a reflex test to any TB positive result,^7^ only Xpert MTB/RIF and Ultra *M. tuberculosis*-positive patients were enrolled in our study, and so no data were available to confirm Xpert MTB/XDR specificity for TB detection. Although previous studies have demonstrated the limit of detection of the Xpert MTB/XDR assay to be equivalent to Xpert MTB/RIF,^10,18^ additional prospective, clinical data would be helpful to support this finding and define best use cases. Furthermore, the study was designed to evaluate a high number of drug-resistant specimens to provide accurate sensitivity estimates, and this was achieved by testing mostly RIF resistant samples, but also meant that sample size was relatively low to assess Xpert MTB/XDR specificity for INH resistance detection. However, given that no false positive results were obtained in this study, overall confidence is high in the specificity of the assay for INH resistance detection.

As a reflex test to an Xpert MTB/RIF or Ultra TB-positive result, the Xpert MTB/XDR assay allows for rapid extended drug resistance profiling directly from sputum, using existing infrastructure and sample processing procedures, thereby providing a decentralized option for optimized TB treatment. Given the importance of INH in TB treatment regimens,^3–6^ as well as the growing concerns regarding INH mono-resistant strains of TB,^20^ the availability of a sputum-based assay with an expanded landscape for INH resistance detection may be of great value to best direct appropriate TB patient treatment in many contexts. Although the definition of XDR-TB was redefined by WHO in 2021, the assay still presents a crucial diagnostic to identify and direct treatment of INH-resistant TB, FQ-resistant TB, and pre-XDR TB, now defined as *M. tuberculosis* strains that fulfil the definition of multidrug-resistant/rifampicin-resistant TB and which are also resistant to any FQ.^21^ The relevance of the assay is underscored given the importance of INH and FQ in current drug-susceptible and drug-resistant treatment regimens,^3–6^and the assay fills the need for rapid FQ DST to all-oral, short course drug-resistant TB regimens. While the importance of determining resistance to SLIs has decreased given changing TB treatment guidelines, there is still value in having this information as SLIs remain in widespread use as countries make the switch to all-oral regimens, as well as for patients who fail on these novel regimens. Additional field studies will be necessary to determine the optimal placement of the assay in existing diagnostic and laboratory algorithms and the acceptability and robustness of the GeneXpert instruments with10-color calibration needed to performr the assay, especially in high burden countries with varying infrastructure capacities.^22^

Overall, the Xpert MTB/XDR assay met the minimal criteria set by the WHO target product profile for a next-generation DST, although sensitivity of the assay for ETH resistance detection was low and performance variations were identified between sites.^15,23^ The introduction and rollout of this rapid sputum-based assay has the potential to greatly improve TB patient diagnosis and management worldwide. Additional studies, including demonstration studies of various implementation approaches, will be critical to define best use cases for the Xpert MTB/XDR assay and ensure optimal impact in improving outcomes for patients with drug-resistant TB.

## Supporting information

Supplement 1 - Protocol

Supplement 2 - WGS protocol

STARD checklist

## Data Availability

Individual, de-identified participant data will be shared, including data dictionaries. Other documents that have been made available include the study protocol and statistical analysis plan. Templates of the informed consent forms may be shared upon request. The data will be available immediately following publication with no end date. The data will be shared with anyone who wishes to access the data. The data will be available for any purpose of analyses. For data, please contact the corresponding author.

## Xpert XDR Trial Consortium members

Catharina Boehme, Sergio Carmona, Sarabjit Chadha, Megha Dhalla, Christine Hoogland, Aurélien Macé, Shweta Mall, Stefano Ongarello, Shakir Reza, Sanjay Sarin, Shubhada Shenai (Foundation for Innovative New Diagnostics, Geneva, Switzerland); Trish Kahamba, Wendy Stevens, Lyndel Singh, Xabisa Makeleni (University of the Witwatersrand, Johannesburg, South Africa); Ritu Singhal; Jyoti Arora; Rohit Sarin (National Institute of TB and Respiratory Diseases, New Delhi, India).

## Acknowledgements

The authors would like to thank the study participants and their families for generously volunteering to participate in this study, as well as the study sites for their time and effort in conducting the study, and assisting with the analysis of the operational data. The authors also thank ACOMED for conduction statistical analysis, MedGenome for WGS and Cepheid for their technical expertise towards the study. Medical writing support was provided by Talya Underwood, MPhil, of Anthos Communications Ltd, UK, funded by the Foundation for Innovative New Diagnostics, according to Good Publication Practice guidelines.

## Contributors

APN, SBG, CMD, and SGS designed the study; APN, SBG, MR, LS, VPM, CR, VC, CMD, SGS oversaw the study. NC, MK, MB, AD, and FC coordinated the individual study sites. Statistical analysis was undertaken by ACOMED. The manuscript drafts were developed by SBG, APN, CMD, and SGS with input from the authors. All authors contributed to interpretation of data and editing of the article and approved the final version of the manuscript.

