## Supplement 1 - Protocol for "Clinical evaluation of the Xpert MTB/XDR assay for rapid detection of isoniazid, fluoroquinolone, ethionamide and second-line drug resistance: A cross-sectional multicentre diagnostic accuracy study"

CONFIDENTIAL

### STUDY PROTOCOL

#### Phase II: Multicentre Clinical Study to Assess the Performance of the Xpert MTB/XDR Assay for INH- and Second-line Resistance Detection

#### Xpert MTB/XDR Clinical Evaluation

Version: v1.1

Date: 05 NOV 2018

Sponsor Name: FIND

Funder: FIND (KFW & NL)

Disease Programme: TB

Regulatory Agency Identifying Number(s):

Clinicaltrials.gov Registration Number: NCT03728725

Foundation for Innovative New Diagnostics
Campus Biotech
Chemin des Mines 9
1202 Geneva
Switzerland
T: +41 (0)22 710 05 90
F: +41 (0)22 710 05 99

#### **Confidentiality Statement**:

The information contained in this document, especially unpublished data, is the property of FIND (or under its control) and may not be reproduced, published or disclosed to others without prior written authorization from FIND.

CONFIDENTIAL

CONFIDENTIAL

#### Institutions/Partners Involved in the Trial

The main institutions/organizations/partners are listed below. More information can be found in the Study Contact List for the names, telephone, e-mail address, title and role on the study.

| Organization/Institution/Company/Partner | Role in the Study |
|---|---|
| FIND, Geneva, Switzerland | Sponsor |
| Hinduja Hospital and Medical Research Centre, Mumbai, India | Principal investigator 1 |
| Phthisiopneumology Institute, Chisinau, Moldova | Principal investigator 2 |
| Wits Health Consortium, Johannesburg, South Africa | Principal investigator 3 |
| National Institute of Tuberculosis and Respiratory Diseases, New Delhi, India | Principal investigator 4 |
| Cepheid | IVD Manufacturer |

**FIND**
Contact person: Claudia Denkinger


Campus Biotech
Chemin des Mines 9
1202 Geneva
Switzerland

**Cepheid**
Contact person: Nova Via


904 E Caribbean Drive
Sunnyvale, CA 94089
United States

**Hinduja Hospital and Medical Research Centre**
Contact person: Camilla Rodrigues


Veer Savarkar Marg
Mahim, Mumbai
Maharashtra 400016, India

CONFIDENTIAL

**Phthisiopneumology Institute**
Contact person: Valeriu Crudu


13 Varnav Str
MD-2025 Chisinau, Moldova

**National Health Laboratory Services**
**Contact person: Professor Wendy Stevens**
****

**31 Princess of Wales Terrace**
**Parktown**
**2193**

**National Institute of Tuberculosis and Respiratory Diseases**
Contact person: Manpreet Bhalla


Sri Aurobindo Marg, Near Qutub Minar, Seth Sarai,
Mehrauli, New Delhi,
Delhi 110030, India

CONFIDENTIAL

*Terms of references and nature of agreements are available from the sponsor on request.

#### Signature Page (Sponsor)

We, the undersigned, have reviewed and approved this Protocol, including Appendices. We will supervise and coordinate the clinical study as described and ensure adherence to GCP/GCLP, the principles outlined in the Declaration of Helsinki and applicable regulatory requirements.

HEAD OF TB PROGRAMME

Name: Claudia Denkinger
Institution: FIND

Signature: ______________________ Date:_______________
DD/MMM/YYYY

HEAD OF CLINICAL & REGULATORY AFFAIRS

Name: Jennifer Kealy
Institution: FIND

Signature: ______________________ Date:_______________
DD/MMM/YYYY

PROJECT MANAGER

Name: Sophia Georghiou
Institution: FIND

Signature: _______________________ Date:_______________
DD/MMM/YYYY

TRIAL MANAGER

Name: Adam Penn-Nicholson
Institution: FIND

Signature: _______________________ Date:_______________
DD/MMM/YYYY

TRIAL STATISTICIAN

Name: Stefano Ongarello
Institution:

Signature: ________________________ Date:_______________
DD/MMM/YYYY

CONFIDENTIAL

DATA MANAGER

Name: Aurélien Macé
Institution: FIND

Signature: ________________________ Date:________________
DD/MMM/YYYY

CONFIDENTIAL

### Statement of Hinduja Hospital and Medical Research Centre Principal Investigator

*All studies sponsored by FIND will be conducted in accordance with ICH-GCP Guidelines, to the extent possible in the research setting.*

In signing this page, I, the undersigned, agree to conduct the study in compliance with all applicable regulations and guidelines as stated in the protocol and other information supplied to me.

I will ensure that the requirements relating to obtaining Institutional Review Board (IRB)/Independent Ethics Committee (IEC) review and approval are met. I will promptly report to the IRB/IEC any and all changes in the research activities covered by this protocol.

I have sufficient time to properly conduct and complete the trial within the agreed trial period and I have adequate resources (staff and facilities) for the foreseen duration of the trial.

I am responsible for supervising any individual or party to whom I delegate trial related duties and functions conducted at the trial site. Further, I will ensure this individual or party is qualified to perform those trial-related duties and functions.

I certify that Individuals involved with the conduct of this trial have completed GCP training within the past 3 years and, if applicable, Human Subjects Protection Training.

I understand that all information obtained during the conduct of the study with regard to the subjects' state of health will be regarded as confidential. No subject's names or personal identifying information may be disclosed. All subject data will be anonymized and identified by assigned numbers on all Case Report Forms, laboratory samples and source documents forwarded to the sponsor. Monitoring and auditing by the sponsor, and inspection by the appropriate regulatory authority(ies), will be permitted.

I will maintain confidentiality of this protocol and all other related investigational materials. Information taken from the study protocol may not be disseminated or discussed with a third party without the express consent of the sponsor.

Name of Principal Investigator: ________________________

(Print)

Site: _____________________________

Signature: _________________________ Date: ____________________

DD/MMM/YYYY

CONFIDENTIAL

### Statement of Phthisiopneumology Institute Principal Investigator

*All studies sponsored by FIND will be conducted in accordance with ICH-GCP Guidelines, to the extent possible in the research setting.*

In signing this page, I, the undersigned, agree to conduct the study in compliance with all applicable regulations and guidelines as stated in the protocol and other information supplied to me.

I will ensure that the requirements relating to obtaining Institutional Review Board (IRB)/Independent Ethics Committee (IEC) review and approval are met. I will promptly report to the IRB/IEC any and all changes in the research activities covered by this protocol.

I have sufficient time to properly conduct and complete the trial within the agreed trial period and I have adequate resources (staff and facilities) for the foreseen duration of the trial.

I am responsible for supervising any individual or party to whom I delegate trial related duties and functions conducted at the trial site. Further, I will ensure this individual or party is qualified to perform those trial-related duties and functions.

I certify that Individuals involved with the conduct of this trial have completed GCP training within the past 3 years and, if applicable, Human Subjects Protection Training.

I understand that all information obtained during the conduct of the study with regard to the subjects' state of health will be regarded as confidential. No subject's names or personal identifying information may be disclosed. All subject data will be anonymized and identified by assigned numbers on all Case Report Forms, laboratory samples and source documents forwarded to the sponsor. Monitoring and auditing by the sponsor, and inspection by the appropriate regulatory authority(ies), will be permitted.

I will maintain confidentiality of this protocol and all other related investigational materials. Information taken from the study protocol may not be disseminated or discussed with a third party without the express consent of the sponsor.

Name of Principal Investigator: ________________________

(Print)

Site: _____________________________

Signature: _________________________ Date: ____________________

DD/MMM/YYYY

CONFIDENTIAL

### Statement of Wits Health Consortium Principal Investigator

*All studies sponsored by FIND will be conducted in accordance with ICH-GCP Guidelines, to the extent possible in the research setting.*

In signing this page, I, the undersigned, agree to conduct the study in compliance with all applicable regulations and guidelines as stated in the protocol and other information supplied to me.

I will ensure that the requirements relating to obtaining Institutional Review Board (IRB)/Independent Ethics Committee (IEC) review and approval are met. I will promptly report to the IRB/IEC any and all changes in the research activities covered by this protocol.

I have sufficient time to properly conduct and complete the trial within the agreed trial period and I have adequate resources (staff and facilities) for the foreseen duration of the trial.

I am responsible for supervising any individual or party to whom I delegate trial related duties and functions conducted at the trial site. Further, I will ensure this individual or party is qualified to perform those trial-related duties and functions.

I certify that Individuals involved with the conduct of this trial have completed GCP training within the past 3 years and, if applicable, Human Subjects Protection Training.

I understand that all information obtained during the conduct of the study with regard to the subjects' state of health will be regarded as confidential. No subject's names or personal identifying information may be disclosed. All subject data will be anonymized and identified by assigned numbers on all Case Report Forms, laboratory samples and source documents forwarded to the sponsor. Monitoring and auditing by the sponsor, and inspection by the appropriate regulatory authority(ies), will be permitted.

I will maintain confidentiality of this protocol and all other related investigational materials. Information taken from the study protocol may not be disseminated or discussed with a third party without the express consent of the sponsor.

Name of Principal Investigator: ________________________

(Print)

Site: ______________________________

Signature: __________________________ Date: ____________________

DD/MMM/YYYY

CONFIDENTIAL

### Statement of National Institute of Tuberculosis and Respiratory Diseases Principal Investigator

*All studies sponsored by FIND will be conducted in accordance with ICH-GCP Guidelines, to the extent possible in the research setting.*

In signing this page, I, the undersigned, agree to conduct the study in compliance with all applicable regulations and guidelines as stated in the protocol and other information supplied to me.

I will ensure that the requirements relating to obtaining Institutional Review Board (IRB)/Independent Ethics Committee (IEC) review and approval are met. I will promptly report to the IRB/IEC any and all changes in the research activities covered by this protocol.

I have sufficient time to properly conduct and complete the trial within the agreed trial period and I have adequate resources (staff and facilities) for the foreseen duration of the trial.

I am responsible for supervising any individual or party to whom I delegate trial related duties and functions conducted at the trial site. Further, I will ensure this individual or party is qualified to perform those trial-related duties and functions.

I certify that Individuals involved with the conduct of this trial have completed GCP training within the past 3 years and, if applicable, Human Subjects Protection Training.

I understand that all information obtained during the conduct of the study with regard to the subjects' state of health will be regarded as confidential. No subject's names or personal identifying information may be disclosed. All subject data will be anonymized and identified by assigned numbers on all Case Report Forms, laboratory samples and source documents forwarded to the sponsor. Monitoring and auditing by the sponsor, and inspection by the appropriate regulatory authority(ies), will be permitted.

I will maintain confidentiality of this protocol and all other related investigational materials. Information taken from the study protocol may not be disseminated or discussed with a third party without the express consent of the sponsor.

Name of Principal Investigator: ________________________

(Print)

Site: _____________________________

Signature: _________________________ Date: ____________________

DD/MMM/YYYY

CONFIDENTIAL

### Protocol History/Amendment Summary*

| Version number | Release date | Comments |
|---|---|---|
| 1.0 | 15 October 2018 | Initial version (updated timeline from 15 Sep 2018 version) |
| 1.1 | 05 November 2018 | Added ETH (ethionamide) to MGIT DST testing, throughout, and updated objectives to reflect testing.<br>Revised study flow language and figure to better illustrate Xpert MTB/RIF screening steps as part of routine diagnostic testing. As such, sputum collection volume need only be ≥3ml |
| 1.1 | 05 February 2019 | Changed NHLS to Wits Health Consortium throughout.<br>Added Wendy Stevens name as WHC PI. |

*Refer to Appendix 3 for Protocol Amendment History

CONFIDENTIAL

### List of Abbreviations and Acronyms

| Abbreviation/acronym | Meaning |
|---|---|
| AE | Adverse Event |
| AMK | Amikacin |
| BSL | Biosafety Level |
| CAP | Capreomycin |
| CRF | Case Report Form |
| DMC | Data Monitoring Committee |
| DR-TB | Drug-resistant TB |
| DST | Drug Susceptibility Testing |
| eCRF | Electronic Case Report Form |
| ETH | Ethionamide |
| GCP | Good Clinical Practice |
| GCLP | Good Clinical Laboratory Practice |
| GDP | Good Documentation Practice |
| ICF | Informed Consent Form |
| ICH | International Council on Harmonisation |
| IDMC | Independent Data-Monitoring Committee |
| IEC | Independent Ethics Committee |
| INH | Isoniazid |
| IRB | Institutional Review Board |
| ISF | Investigator Site File |
| ISO | International Organization for Standardization |
| IUO | Investigation Use Only |
| IVD | *in vitro* Diagnostic |
| KAN | Kanamycin |
| LOD | Limit of Detection |
| LPA | Line Probe Assay |
| MDR | Multidrug-resistant |

CONFIDENTIAL

| | |
|---|---|
| MTB | *Mycobacterium tuberculosis* |
| NGS | Next Generation Sequencing |
| NTP | National TB Programme |
| QA | Quality Assurance |
| QC | Quality Control |
| QMS | Quality Management System |
| RA | Regulatory Authority |
| RBM | Risk Based Monitoring |
| RM | Risk Management |
| RIF | Rifampicin |
| SAE | Serious Adverse Event |
| SAP | Statistical Analysis Plan |
| SNRL | Supranational Reference Laboratory |
| SOP | Standard Operating Procedure |
| TB | Tuberculosis |
| TMF | Trial Master File |
| WHO | World Health Organization |
| XDR | Extensively Drug-resistant |

CONFIDENTIAL

### Protocol Synopsis

| | |
|---|---|
| **Title** | Phase II: Multicentre Clinical Study to Assess the Performance of the Xpert MTB/XDR Assay for INH- and Second-line Resistance Detection |
| **Short title** | Xpert MTB/XDR Clinical Evaluation |
| **Protocol version and date** | v1.1 05-NOV-2018 |
| **Background and rationale** | Evidence of the clinical diagnostic accuracy and operational characteristics of the Xpert MTB/XDR assay is needed to comprehensively evaluate Xpert MTB/XDR validity and inform global and national policy decision-making. |
| **Objectives** | Primary objectives<br>1.1 Estimate the diagnostic accuracy of the Xpert MTB/XDR assay for INH and ETH resistance detection<br>1.2 Estimate the diagnostic accuracy of the Xpert MTB/XDR assay for fluoroquinolone resistance detection<br>1.3 Estimate the diagnostic accuracy of the Xpert MTB/XDR assay for second-line injectable resistance detection<br>1.4 Assess Xpert MTB/XDR technical performance, including non-determinant rates, ease of use and other systems operational characteristics.<br>Secondary objectives<br>2.1 Assess additional Xpert MTB/XDR performance characteristics, including direct performance versus performance on cultured samples, performance between sites, by smear result, by gene target and compared to Hain MTBDRplus and MTBDRsl |
| **Study design** | Multicentre, cross-sectional diagnostic accuracy study |
| **Study sites/setting** | Clinical sites with high rates of drug-resistant TB in India, Moldova and South Africa |
| **Study population** | Patients with pulmonary TB symptoms and at least one DR-TB risk factor will be screened by Xpert MTB/RIF or Ultra. Patients with a clear TB-positive and RIF-resistant or RIF-sensitive result by Xpert MTB/RIF or Ultra and who consent to study procedures will be tested by Xpert MTB/XDR. An anticipated 284 TB-positive and 316 additional RIF-resistant patients will be enrolled in this study. |
| **Eligibility criteria** | Screening criteria:<br>• Age 18 years and above<br>• Symptoms suggesting pulmonary TB, i.e. persistent cough (generally ≥2 weeks or as per local definition of TB suspect) and at least one risk factor for DR-TB<br>Inclusion criteria:<br>• A clear TB-positive and RIF-resistant or -sensitive result by Xpert MTB/RIF or Xpert MTB/RIF Ultra<br>• Provision of Informed Consent<br>• Production of an adequate quantity (≥3mL) of sputum |
| **Primary endpoints** | 1.1 Sensitivity and specificity estimates for INH and ETH resistance detection<br>1.2 Sensitivity and specificity estimates for fluoroquinolone resistance detection<br>1.3 Sensitivity and specificity estimates for second-line injectable resistance detection |

CONFIDENTIAL

| | |
|---|---|
| | 1.4 Assessment of Xpert MTB/XDR technical performance |
| **Secondary endpoints** | 2.1 Assessment of additional Xpert MTB/XDR performance characteristics |
| **Study duration** | 10 weeks for IVD testing, with an additional 15 weeks to complete culture, DST, and NGS |
| **Time schedule** | IRB/IEC Approval: June-December 2018<br>Shipment: January-March 2019<br>Training: March 2019<br>All Testing: April-September 2019<br>Data Cleaning: September-October 2019<br>Preliminary Analysis: October 2019 |
| **GCP statement** | This study will be conducted in compliance with the protocol, the Declaration of Helsinki, ICH-GCP, ISO 13485: 2016 (if applicable) as well as all national legal and regulatory requirements. |

CONFIDENTIAL

### Project Timeline

| | | | |
|---|---|---|---|
| ▼ **Clinical Studies** | **528 d** | **Oct 25, 2017 at…** | **Nov 1, 2019 at…** |
| ▼ **Study Conception and Preparation** | **310 d** | **Oct 25, 2017 at…** | **Jan 1, 2019 at…** |
| Write and Review Clinical Protocols | 24 w | Oct 25, 2017 at… | Apr 10, 2018 at… |
| Site Selection and Contracts | 24 w | Apr 11, 2018 at… | Sep 25, 2018 at… |
| Preparation of TMF | 24 w | Apr 11, 2018 at… | Sep 25, 2018 at… |
| IRB Approvals | 14 w | Sep 26, 2018 at… | Jan 1, 2019 at 5… |
| ▼ **Study Conduct** | **330 d** | **Jul 30, 2018 at…** | **Nov 1, 2019 at…** |
| ▼ **Phase I: Lab Validation** | **170 d** | **Jul 30, 2018 at…** | **Mar 22, 2019 at…** |
| Strain Selection | 12 w | Jul 30, 2018 at… | Oct 19, 2018 at… |
| Shipment and Customs Clearance | 8 w | Oct 22, 2018 at… | Dec 14, 2018 at… |
| Site Training and Initiation | 1 w | Jan 21, 2019 at… | Jan 25, 2019 at… |
| Testing, Data Entry and Cleaning | 4 w | Jan 28, 2019 at… | Feb 22, 2019 at… |
| Analysis | 2 w | Feb 25, 2019 at… | Mar 8, 2019 at… |
| Data Review and Report | 2 w | Mar 11, 2019 at… | Mar 22, 2019 at… |
| ▼ **Phase II: Clinical Trial** | **240 d** | **Dec 3, 2018 at…** | **Nov 1, 2019 at…** |
| Import Permits | 8 w | Dec 3, 2018 at… | Jan 25, 2019 at… |
| Shipment and Customs Clearance | 7 w | Jan 28, 2019 at… | Mar 15, 2019 at… |
| Site Training and Initiation | 2 w | Mar 18, 2019 at… | Mar 29, 2019 at… |
| Testing of Samples | 10 w | Apr 1, 2019 at 8… | Jun 7, 2019 at 5… |
| Completion of Culture/DST/NGS | 15 w | Jun 10, 2019 at… | Sep 20, 2019 at… |
| Data Entry and Cleaning | 2 w | Sep 23, 2019 at… | Oct 4, 2019 at 5… |
| Data Analysis | 2 w | Oct 7, 2019 at 8… | Oct 18, 2019 at… |
| Data Review and Report | 2 w | Oct 21, 2019 at… | Nov 1, 2019 at… |

CONFIDENTIAL

### 1 Introduction

FIND and partners intend to address the need for a multi- and extensively drug-resistant tuberculosis (M/XDR-TB) diagnostic solution for patients in settings with a high burden of drug-resistant tuberculosis (DR-TB) though the development, evaluation and introduction of an Xpert MTB/XDR assay.

#### 1.1 Study Rationale

Evidence of the clinical diagnostic accuracy and operational characteristics of the Xpert MTB/XDR assay is needed to comprehensively evaluate assay validity in settings of intended use and inform global and national policy decision-making.

#### 1.2 Potential Risks and Benefits

This study has no known medical risks for participants. Sample inclusion in this study will not affect TB or M/XDR-TB patient routine care and treatment. No clinical decisions will be made from Xpert MTB/XDR results. The possibility of unknown or unforeseen risk from specimen collection and diagnostic testing is minimal. Knowledge gained from this study may benefit patient communities by improving DR-TB diagnosis. Given the minimal risks associated with this study and the potential benefits to society and individuals, the benefits outweigh the aggregated risks.

If unforeseen risks are recognized during the study, then FIND, study partners, Institutional Review Boards (IRBs)/Independent Ethics Committees (IECs), and participants must be provided with relevant information.

#### 1.3 Background

In recent years, tuberculosis (TB) control efforts have been complicated by the rise and spread of MDR-TB, or TB that is resistant to the first-line drugs isoniazid (INH) and rifampicin (RIF), and XDR-TB, or MDR-TB that has developed additional resistance to a fluoroquinolone and any of the injectable compounds [amikacin (AMK), kanamycin (KAN) and/or capreomycin (CAP)][1,2]. The rapid diagnosis and appropriate treatment of M/XDR-TB is essential to prevent significant morbidity, mortality and further transmission of disease. For treatment of uncomplicated MDR-TB, the World Health Organization (WHO) recently endorsed a 6-9 month treatment regimen, thereby replacing conventional 18-24 month regimens[3]. The fluoroquinolones and second-line injectables are key components of the 6-9 month regimen, and so it is necessary to rule-out resistance to these compounds prior to treating patients with the shorter regimen.

CONFIDENTIAL

The Xpert MTB/RIF assay (Cepheid, Sunnyvale, CA) is an integrated, automated, cartridge-based system for MDR-TB diagnosis that uses the GeneXpert instrument platform. WHO confirmed evidence to support the widespread use of the Xpert MTB/RIF assay in 2010 and the assay has since been widely used in TB programs[4], but it is only capable of identifying *Mycobacterium tuberculosis* (Mtb) and detecting RIF resistance, and so it does not provide the comprehensive resistance profile necessary to determine if the shorter TB treatment regimen is appropriate for certain patients. Conventional diagnosis of TB drug resistance relies upon the slow growth of Mtb in solid or liquid media, only then followed by resistance testing to determine phenotypic drug resistance. These conventional drug susceptibility testing (DST) methods can take several weeks to yield results[5-6], require significant laboratory infrastructure and training, and are potentially biohazardous. In view of the inadequacy of these conventional tests for resistance detection, the development of rapid tests for INH and second-line resistance detection has become a research and implementation priority.

Rapid diagnostics for second-line drug resistance detection identify mutations in the *gyrA* and *gyrB* genes[7], associated with fluoroquinolone resistance, and *rrs* and *eis* gene regions, associated with second-line injectable resistance (with *eis* promoter mutations exclusively associated with KAN resistance)[8]. Recently, a novel GeneXpert cartridge was developed to detect mutations occurring in these genes, as well as in the *katG* and *inhA* gene regions, associated with INH resistance[9]. A Research Use Only (RUO) version of the Xpert MTB/XDR cartridge showed promising performance for INH and second-line resistance detection in a clinical evaluation study (sensitivity 92.7-98.1%; specificity 94.3-99.6%)[10]. The assay chemistry has since been improved and additional gene targets have been added to the assay to improve the detection of INH resistance (i.e. *fabG1* and *ahpC*).

Evidence of assay accuracy from an external laboratory validation using a well-characterized set of Mtb strains as well as from large-scale, multicentre clinical studies is needed to confirm the validity of the most recent Xpert MTB/XDR assay for INH and second-line resistance detection, and to recommend its use in diverse clinical settings. The focus of this protocol is the multicentre clinical evaluation.

CONFIDENTIAL

### 2 Study Objectives and Endpoints

| Objectives | Data Collected | Endpoint |
|---|---|---|
| Primary | | |
| 1.1 **Estimate the diagnostic accuracy of the Xpert MTB/XDR assay for INH and ETH resistance detection**, for a clinically diverse set of TB samples, with phenotypic DST and *katG, fabG1*, *ahpC* and *inhA* | 284 Xpert MTB/RIF TB-positive and 316 RIF-resistant clinical samples tested by Xpert MTB/XDR, phenotypic DST and NGS | 1.1 Sensitivity and specificity estimates for INH and ETH resistance detection |

| | | |
|---|---|---|
| sequencing results as a composite reference standard.<br>**1.2 Estimate the diagnostic accuracy of the Xpert MTB/XDR assay for fluoroquinolone resistance detection**, for a clinically diverse set of TB samples, with phenotypic DST and *gyrA* and *gyrB* sequencing results as a composite reference standard.<br>**1.3 Estimate the diagnostic accuracy of the Xpert MTB/XDR assay for second-line injectable resistance detection**, for a clinically diverse set of TB samples, with phenotypic DST and *rrs* and *eis* sequencing results as a composite reference standard. | | 1.2 Sensitivity and specificity estimates for fluoroquinolone resistance detection<br>1.3 Sensitivity and specificity estimates for second-line injectable resistance detection |
| **1.4 Assess Xpert MTB/XDR operational aspects and ease of use,** including Xpert MTB/XDR non-determinant rates. | Completion of laboratory CRFs for all samples and completion of user-appraisal questionnaires by at least two operators per site at two different timepoints (after training and after study completion) | 1.4 Assessment of Xpert MTB/XDR operational aspects and ease of use |
| Secondary | | |
| **2.1 Assess additional Xpert MTB/XDR performance characteristics:**<br>o Direct performance versus performance on cultured samples<br>o Performance between sites<br>o Performance by smear result<br>o Performance by gene target<br>o Performance compared to Hain MTBDR*plus/sl*<br>o Performance by patient HIV status<br>o Performance by patient pre-treatment status<br>o Performance as reflex test to Ultra or MTB/RIF assay | Smear, culture, NGS, MTBDR*plus*, MTBDR*sl* and two Xpert MTB/XDR results (direct from sputum and from cultured strain) for each clinical sample tested | 2.1 Assessment of additional Xpert MTB/XDR performance characteristics |

### 3 Study Design

CONFIDENTIAL

#### 3.1 General Design

Multicentre clinical diagnostic accuracy study.

#### 3.2 Scientific Rationale for Study Design

The current study is part of a two-phase project.

- Phase 1 (the current study) is an analytical study to validate and expand upon the Xpert MTB/XDR analytical data that the manufacturer (Cepheid, Sunnyvale, California) has compiled for internal validation, CE-IVD marking and/or other regulatory purposes.
- Phase 2 is a clinical study to determine the diagnostic accuracy at intended settings of use.

The two phases will provide complementary pieces of evidence to address the study objectives.

The main purpose of this clinical study is to confirm accuracy estimates from the external laboratory validation and ensure that Xpert MTB/XDR diagnostic performance characteristics will be consistent at sites of intended use. Assay performance when testing sputum samples (direct testing) and and cultured isolates (indirect testing), assay performance compared to the Hain MTBDR*plus* and MTBDR*sl* (Hain Lifesciences GmbH) line probe assays (LPAs), and assay error and failure rates and device operational characteristics will also be assessed during this phase of the study.

In order to address clinical study objectives quickly and efficiently, this study will first assess Xpert MTB/XDR performance for Xpert MTB/RIF MTB-positive samples from patients prospectively enrolled in the study. Focusing the study on Xpert MTB/RIF MTB-positive samples is in line with the intended use of the assay as a reflex, or follow-up, test to any Xpert MTB/RIF or Ultra MTB-positive result. This will also allow for an assessment of assay performance for INH resistance detection among all MTB-positive samples including INH mono-resistant, as this population might be different from a largely RIF-resistant population. However, to ensure numbers are adequate for an assessment of Xpert MTB/XDR assay performance for second-line resistance detection, once sufficient MTB-positive, RIF-sensitive samples have been tested, a subset of solely Xpert RIF-resistant samples will be tested until enrolment targets are met (see section 9.1 and Appendix 4: Sample Requirement to Achieve Target Sample Size).

#### 3.3 Study Setting

Study sites will be geographically diverse and representative of regional epidemics (i.e. TB and resistance prevalence, HIV co-infection, lineage distribution). Laboratories will represent the intended setting of use for the Xpert MTB/XDR assay,

CONFIDENTIAL

i.e. laboratories that routinely process a large number of DR-TB samples and wherein second-line drug resistance may be suspected. Site preference will be given to countries/sites that are able to be early adopters (in terms of both political will and financial means) and sites with strong National TB Programme (NTP) relationship and global reach in order to gain visibility and encourage buy-in to facilitate national and global policy uptake. Participating sites must also have a sufficiently high throughput of TB clinical samples, and have access to phenotypic DST and NGS testing to allow for the efficient assessment of Xpert MTB/XDR assay performance.

**Regional distributions and country options** (to be confirmed):

- Africa (South Africa)
- Asia (India)
- Eastern Europe (Moldova)

#### 3.4 End of Study Definition

Phase II will be considered completed once all *in vitro* diagnostic (IVD) testing has completed and all culture, DST and NGS results have been returned for at least 284 TB-positive and 316 additional RIF-resistant clinical samples, and all user appraisal questionnaires have been completed at the three clinical sites.

### 4 Study Population and Eligibility

Individuals who have one or more risk factors for DR-TB presenting to participating centres will be screened by Xpert MTB/RIF or Ultra. Those who test MTB-positive by Xpert MTB/RIF or Ultra at the study sites will be asked to participate. Once at least 360 MTB-positive patients have been tested, an additional subset of Xpert MTB-positive, RIF-resistant patients (n=400) will also be asked to participate in this study. Individuals will be recruited at outpatient clinic settings and inpatient hospital settings. Individuals will be asked by non-study clinicians or staff if they would be interested in participating in the study. Interested individuals will be referred to study personnel for additional information and screening, if appropriate. HIV-positive individuals and HIV-negative individuals will be included in this study.

Prospective approval of protocol deviations to recruitment and enrolment criteria, also known as protocol waivers or exemptions, is not permitted.

#### 4.1 Inclusion Criteria:

Patients meeting the following criteria will be screened by Xpert MTB/RIF or Xpert MTB/RIF Ultra:

CONFIDENTIAL

- Age 18 years or above;
- Symptoms suggesting pulmonary TB, i.e. persistent cough (generally ≥3 weeks or as per local definition of TB suspect), **and** at least one of the following:
    - Previously received >1 month of treatment for a prior TB episode **or**
    - Failing TB treatment with positive sputum smear or culture after ≥3 months of a standard TB treatment **or**
    - Had close contact with a known drug-resistant TB case **or**
    - Newly diagnosed with MDR-TB within the last 30 days **or**
    - Previously diagnosed with MDR-TB and failed TB treatment with positive sputum smear or culture after ≥3 months of a standard MDR-TB treatment regimen

Patients meeting the above criteria will be screened by Xpert MTB/RIF or Xpert MTB/RIF Ultra. TB patients meeting the following criteria will be included in the study**:**

- A clear Mtb-positive and RIF-resistant or RIF-sensitive result by Xpert MTB/RIF or Xpert MTB/RIF Ultra
- Provision of informed consent;
- Production of an adequate quantity (≥3mL) of sputum

#### 4.2 Exclusion Criteria:

Participants will be excluded from the study if informed consent is not provided.

#### 4.3 Early Exclusions

Participants who have provided consent and who are enrolled, but who do not provide an adequate quantity of sputum at the start of the study, will be classified as early exclusions; each will be removed from the study and a new study subject enrolled. Participants who are classified as early exclusions will be referred to the appropriate local health service for TB evaluation and care. If positive for Mtb, results of conventional microbiological tests (e.g., sputum smear microscopy, mycobacterial cultures, and/or Xpert MTB/RIF) will be reported to appropriate local health authorities in accordance with local public health reporting regulations. Study data for participants who are classified as early exclusions will not be used for any final analyses.

#### 4.4 Participant Discontinuation/Withdrawal from Study

A participant may be withdrawn at any time at the discretion of the investigator for safety, behavioural, compliance, or administrative reasons.

CONFIDENTIAL

If the participant withdraws consent for disclosure of future information, the sponsor may retain and continue to use any data collected before such a withdrawal of consent.

If a participant withdraws from the study, he/she may request destruction of any samples taken and not tested, and the investigator must document this in the site study records.

See Schedule of Assessments for data and/or samples to be collected at the time of study discontinuation and follow-up and for any further evaluations that need to be completed.

### 5 Study Intervention

Study Intervention is defined as any investigational intervention(s), marketed product(s), or medical device(s) intended to be used with a study participant according to the study protocol.

#### 5.1 *in vitro* Diagnostics

The IVD manufactured for FIND use in this study is the Xpert MTB/XDR assay. The Investigational Use Only (IUO) product is intended for the detection of Mtb and mutations associated with INH, ETH and second-line drug resistance. Likewise, the 10-color GeneXpert system provided for this study should only be used for testing IUO cartridges and study samples. The investigational product and 10-color system will be strictly accounted for, including receipt and inventory, storage, use during the trial, and return or disposal, as detailed in the Study Manual for this study.

Other IVDs (not manufactured for FIND) obtained for use in this study are:

- Xpert MTB/RIF assay and/or the Xpert MTB/RIF Ultra assay
- Hain MTBDR*plus* assay
- Hain MTBDR*sl* assay

Instructions for use of these assays will be provided in the SOPs for this study.

Any IVD incidents, including those resulting from malfunctions of the IVD, must be detected, documented and reported by the investigator throughout the study (see section 8.1.4).

#### 5.2 Preparation/Handling/Storage/Accountability

##### 5.2.1 Acquisition

Procurement of the Xpert MTB/XDR assays will be done through FIND, which will coordinate shipments from the manufacturer. It is the responsibility of each laboratory to maintain an updated inventory of the study materials and to inform FIND immediately if additional materials are required.

CONFIDENTIAL

It is expected that the respective countries will require import permits for receiving the investigational materials. The laboratory is responsible for obtaining relevant import permits in a timely manner. The sponsor and IVD manufacturer may support the laboratory to obtain import permits. Upon arrival of each new shipment of Xpert assays or reagents, the laboratory will conduct and document an incoming quality check following the Manual of Procedures. New lots may only be used after this quality check is successfully passed.

##### 5.2.2 Handling

The investigator or designee must confirm appropriate temperature conditions have been maintained during transit for all Xpert MTB/XDR assays received and any discrepancies are reported and resolved before use of the Xpert MTB/XDR assay.

Only those participants enrolled in the study may have their samples tested by the Xpert MTB/XDR assay, and only authorized and trained staff may operate the Xpert MTB/XDR assay. Testing using the investigational product will be performed according to the manufacturer’s instructions outlined within the SOP.

##### 5.2.3 Storage

Procedures for product storage and disposal will be described in the SOP. All Xpert MTB/XDR assays must be stored in a secure, environmentally controlled, and monitored (manual or automated) area in accordance with the labeled storage conditions with access limited to the investigator and authorized site staff.

##### 5.2.4 Accountability

The investigator, institution, or the head of the medical institution (where applicable) is responsible for Xpert MTB/XDR accountability, reconciliation, and record maintenance (i.e., receipt, reconciliation, and final disposition records).

Further guidance and information for the final disposition of unused study interventions are provided in the SOP.

#### 5.3 Minimization of Error and Bias

##### 5.3.1 Patient Selection

A consecutive series of Xpert MTB/RIF MTB-positive patients (n=284) will be enrolled in this study, followed by an additional, consecutive series of Xpert MTB/RIF RIF-resistant patients (n=316) between the three countries with participating clinical sites. Enrolment will be based upon clearly defined eligibility criteria.

CONFIDENTIAL

Analyses will be conducted for relevant sub-groups of the enrolled TB patient population (i.e. smear status for TB, drug-resistance profile, mutations detected, etc.), which may be considered to be indicators of disease severity.

##### 5.3.2 Index Test

The Xpert MTB/XDR assay detects Mtb mutations associated with INH, ETH and second-line drug resistance direct from sputum. In line with this intended use, clinical samples will be directly tested by the Xpert MTB/XDR assay. However, the assay will also be used to test the culture isolate. This additional Xpert MTB/XDR assay testing will allow for a comparison of performance and non-determinant rates between testing directly from the sample and testing from the culture isolate. It will also support resolution of potential discordant LPA and NGS results from the initial Xpert MTB/XDR test result, as these tests will also be performed from the culture isolate (discordance may arise as a result of infections with a mix of different strains, with only one strain being grown out in culture). Furthermore, as all assays will be performed on the same culture, there is also no risk of any assay being tested on a different quality sample.

Review bias will be avoided in this study as the index test requires no manual interpretation by the operator.  All results are automatically generated by the GeneXpert instrument and will be directly recorded by the operator on the laboratory case report form (CRF).

##### 5.3.3 Reference Standard and Comparators

There is little risk of reference standard bias in this study as a composite reference standard of phenotypic DST and NGS will be used for resistance detection (Table 1). The use of both phenotypic and genotypic information will ensure high confidence in the reference standard, as certain mutations (e.g. many *eis* promoter mutations) are known to confer only low-levels of phenotypic drug resistance, and so they may not be detected by phenotypic DST at only one critical concentration. Furthermore, all molecular assays evaluated in this study detect resistance-mutations in similar gene regions, and so all mutations that are detectable by the Xpert MTB/XDR assay are generally detected by NGS and/or the Hain LPAs and vice versa.

**Table 1.** Composite reference standard for diagnostic performance analysis

| | Composite Reference Standard | |
|---|---|---|
| **Drug of Interest** | **NGS of Reference Gene(s)** | **MGIT960 Phenotypic DST** |
| Isoniazid | *katG*, *fabG1*, *inhA* promoter, *ahpC* promoter | Isoniazid |
| Ethionamide | *inhA* promoter | Ethionamide |

CONFIDENTIAL

| | | |
|---|---|---|
| Fluoroquinolones | | |
| Moxifloxacin | *gyrA*, *gyrB* | Moxifloxacin |
| Levofloxacin | *gyrA*, *gyrB* | Levofloxacin |
| Second-line Injectables | | |
| Amikacin | *rrs* | Amikacin |
| Kanamycin | *rrs*, *eis* promoter | Kanamycin |
| Capreomycin | *rrs* | Capreomycin |

##### 5.3.4 Laboratory Methods

The participating sites will undergo an on-site laboratory and clinical evaluation during the site initiation visit to ensure standardized and high-quality performance of molecular methods.

### 6 Study Procedures

Study procedures and their timing are summarized in the Schedule of Assessments. Protocol waivers or exemptions are not allowed.

Adherence to the study design requirements, including those specified in the Schedule of Assessments, is essential and required for study conduct.

All screening evaluations must be completed and reviewed to confirm that potential participants meet all eligibility criteria. The investigator will maintain a screening log to record details of all participants screened and to confirm eligibility or record reasons for screening failure, as applicable.

Procedures conducted as part of the participant's routine clinical management (e.g., blood count) and obtained before signing of the ICF may be utilized for screening or baseline purposes provided the procedures met the protocol-specified criteria and were performed within the time frame defined in the Schedule of Assessments.

##### 6.1.1 Sample Selection

Up to two sputum samples (≥3mL) will be collected from consecutive, prospectively-enrolled patients with defined risk factors for DR-TB and a valid Xpert MTB/RIF or Xpert MTB/RIF Ultra result. Depending upon enrolment rates, study timelines may be accelerated by also using existing, well-characterized frozen TB clinical (sputum) samples collected during the course of routine clinical care and TB surveillance in the sites of intended use. These samples will only be used if recruitment or enrolment is slow at one or more sites. At the 4-week study time point, recruitment rates will be assessed to determine whether this use of additional samples will be necessary.

CONFIDENTIAL

##### 6.1.2 Study Workflow

Study flow is shown in Figure 1. All available Xpert MTB/RIF TB-positive samples will be tested in this study (i.e. consecutive patient samples) until sample size targets are met. All molecular assays will be performed in accordance with SOPs provided by the manufacturers. Specimen handling and processing by routine tests will be carried out according to NTP local policies and standards.

Samples will first be tested by Xpert MTB/RIF or Ultra, as part of routine diagnostic procedures. This Xpert MTB/RIF step will act as a screening tool for trial enrolment. Participants testing positive for MTB by Xpert MTB/RIF will be asked to volunteer for the Xpert MTB/XDR trial. This (testing of specimens from TB-positive samples) is in line with the intended use of the XDR assay as an add-on test to the Xpert MTB/RIF assay.

A staged enrolment will proceed: Stage 1 will enrol 360 MTB-positive participants, irrespective of RIF resistance/sensitivity. Once enrolment is complete, Stage 2 will enrol 400 MTB-positive RIF-resistant participants (see section 9.1 and Appendix 4: Sample Requirement to Achieve Target Sample Size). This staging is designed to better evaluate Xpert MTB/XDR sensitivity for second-line resistance detection.

In order to efficiently assess the diagnostic performance of the Xpert MTB/XDR assay, participants will be asked to provide ≥3ml of sputum (either a single sputum or two consecutively collected and pooled sputa). Samples will be homogenized and split for testing.

The Xpert MTB/XDR assay and acid-fast bacilli smear will be performed on the direct sample. 2ml of sputum will be decontaminated and used for MGIT and LJ culture.

Following direct testing, phenotypic MGIT DST will be performed for all culture-positive samples for INH, RIF, fluoroquinolone (moxifloxacin and levofloxacin), AMK, ETH (ethionamide), KAN and CAP. Cultured samples will also undergo subsequent molecular testing by LPA and targeted NGS of relevant gene regions (*katG, inhA, fabG1, ahpC, gyrA, gyrB, rrs,* and *eis*) and another Xpert MTB/XDR assay. NGS will be performed from the culture isolate to obtain sequencing reads of high quality.

CONFIDENTIAL

**Figure 1.** Phase II sample flow

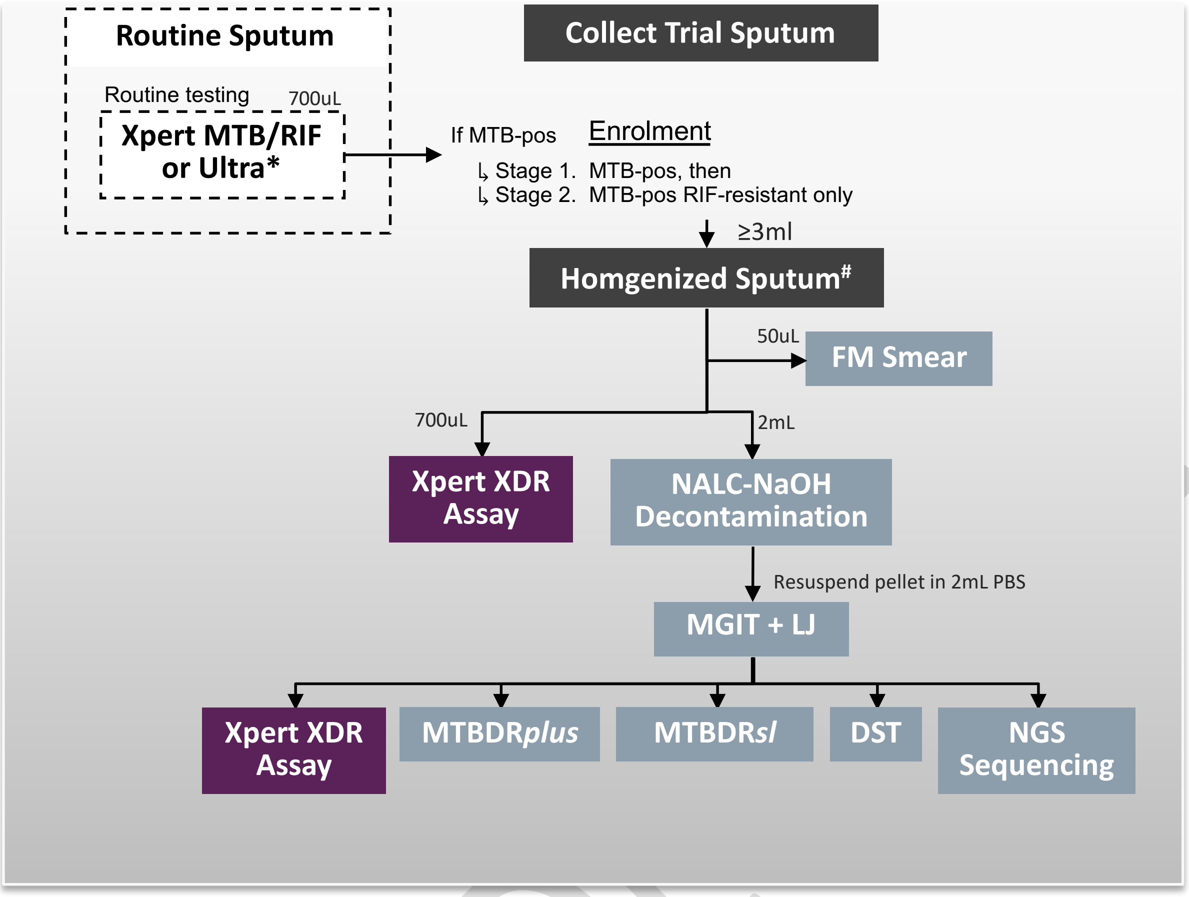

### ≥ 3ml of sputum is required for the trial. This can be achieved through an individual sputum or pooling of serially collected sputa, homogenized with glass beads.

All diagnostic test results (smear, Xpert MTB/XDR on direct sample, MGIT, LJ, Xpert MTB/XDR on culture isolate, MTBDR*plus*, MTBDR*sl*, DST and NGS results) will be recorded by the laboratory technicians for each site on the laboratory CRF.

### 7 Specimen Collection, Handling, Transport and Storage

Clinical (sputum) samples from enrolled patients will be used in this study. Patients will be asked to provide up to two sputum samples for testing after providing informed consent for the study. Sputum samples from the same patient will be pooled and used directly for acid-fast bacilli smear, Xpert MTB/XDR and MGIT and LJ culture.

Appropriate laboratory staff at each site reference laboratory are responsible for receiving the samples, storing the samples and conducting all testing in line with BSL3 requirements. All leftover samples must be stored until study conclusion and completion of all analyses, in case any additional or repeat testing is requested by the sponsor.

#### 7.1 Reference Standard Test and Index Test Procedures

A composite reference standard of phenotypic DST and NGS will be used for resistance detection to ensure high confidence in the reference standard for this study. Phenotypic MGIT DST and targeted NGS will be performed on the culture isolate for each clinical sample. MGIT DST will be done on site to determine culture drug-susceptibility to INH, ETH, moxifloxacin, levofloxacin, AMK, KAN and CAP at the revised WHO critical concentrations[15]. Targeted NGS of the gene regions included in the Xpert MTB/XDR assay will be performed on DNA extracted from the cultured sample, which will be sent to a SNRL for NGS, or sequenced locally using primers specific for the gene regions included in the assay. The output for both phenotypic DST and NGS results are automated, and require no manual operator interpretation. Testing results, when obtained, will be input onto the laboratory CRF.

The index test will be performed on both the direct clinical sample as well as the culture isolate, to ensure high Xpert MTB/XDR diagnostic performance direct from sputum and to provide a basis for comparison if any discordants are seen between the initial Xpert MTB/XDR result and the phenotypic DST and additional molecular test results (NGS and LPA). The output for the Xpert MTB/XDR assay is automated and does not require manual interpretation. As with the reference test, all Xpert test results will noted on the laboratory CRF.

#### 7.2 Genetics

No patient genetic information will be collected, analysed or recorded in this study. The proposed genetic sequencing and molecular analysis protocols use only TB pathogen-specific sequences and will be limited to analysis of *M. tuberculosis* bacteria genomes.

#### 7.3 Biomarkers

Biomarkers are not evaluated in this study.

### 8 Safety and Incident Reporting

The definitions of an adverse event (AE) and serious adverse event (SAE) can be found in Appendix 1: Safety Definitions and Reporting.

AE will be reported by the participant spontaneously (or, when appropriate, by a caregiver, surrogate, or the participant's legally authorized representative).

The investigator and any qualified designees are responsible for detecting, documenting, and recording events that meet the definition of an AE or SAE and remain responsible for following up AEs that are serious, considered related to the study intervention or study procedures, or that caused the operator to discontinue

CONFIDENTIAL

use of the Xpert MTB/XDR assay or participants to discontinue their participation in the Xpert/MTB XDR Clinical Evaluation.

##### 8.1.1 Time Period for Collecting SAE Information

All SAEs will be collected from the signing of the informed consent form (ICF) OR start of Xpert MTB/XDR testing until the end of the Xpert MTB/XDR Clinical Evaluation at the time points specified in the SOP.

All SAEs will be recorded and reported to the sponsor or designee within 24 hours, as indicated in Appendix 2: Incident Definition and Reporting. The investigator will submit any updated SAE data to the sponsor within 24 hours of it being available.

Investigators are not obligated to actively seek SAEs after conclusion of the study participation. However, if the investigator learns of any SAE, including a death, at any time after a participant has been discharged from the study, or the IVD operators have completed IVD testing, and he/she considers the event to be reasonably related to the study intervention or study participation, the investigator must promptly notify the sponsor.

The method of recording, evaluating, and assessing causality of the SAE and the procedures for completing and transmitting SAE reports are provided in Appendix 1: Safety Definitions and Reporting.

##### 8.1.2 Reporting and Follow up of SAEs

All SAEs will be followed until resolution, stabilization, the event is otherwise explained, or the participant is lost to follow-up (as defined in Section **Error! Reference source not found.**). Further information on follow-up procedures is given in Appendix 2: Incident Definition and Reporting)

##### 8.1.3 Regulatory Reporting Requirements for SAEs

Prompt notification by the investigator to the sponsor of a SAE is essential so that legal obligations and ethical responsibilities towards the safety of participants and the safety of a study intervention under clinical investigation are met.

The sponsor has a legal responsibility to notify both the local regulatory authority and other regulatory agencies about the safety of a study intervention under clinical investigation. The sponsor will comply with country-specific regulatory requirements relating to safety reporting to the regulatory authority, IRB/IEC, and investigators.

An investigator who receives an investigator safety report describing a SAE or other specific safety information (e.g., summary or listing of SAEs) from the sponsor will review and then file it in the Investigator Site File (ISF) and will notify the IRB/IEC, if appropriate according to local requirements.

CONFIDENTIAL

##### 8.1.4 IVD Incidents (including Malfunctions)

IVDs are being provided for use in this study for investigative purposes. In order to fulfil regulatory reporting obligations worldwide, the investigator is responsible for the detection and documentation of events meeting the definitions of incident or malfunction that occur during the study with such devices.

The definition of an IVD Incident can be found in Appendix 2: Incident Definition and Reporting.

NOTE: Incidents fulfilling the definition of an AE/SAE will also follow the processes outlined above and in Appendix 1: Safety Definitions and Reporting of the protocol.

##### 8.1.5 Time Period for Detecting IVD Incidents

IVD incidents or malfunctions of the device that result in an incident will be detected, documented, and reported during all periods of the study in which the IVD is used.

If the investigator learns of any incident at any time after a participant has been discharged from the study, and such incident is considered reasonably related to a IVD provided for the study, the investigator will promptly notify the sponsor.

The method of documenting IVD Incidents is provided in Appendix 2.

##### 8.1.6 Follow-up of IVD Incidents

All IVD incidents involving an AE will be followed and reported in the same manner as other AEs (see Section 8.1.2). This applies to all participants, including those who discontinue study intervention.

The investigator is responsible for ensuring that follow-up includes any supplemental investigations as indicated to elucidate the nature and/or causality of the incident.

New or updated information will be recorded on the originally completed form with all changes signed and dated by the investigator.

##### 8.1.7 Reporting of IVD Incidents to Sponsor

IVD incidents will be reported to the sponsor within 24 hours after the investigator determines that the event meets the protocol definition of a IVD incident.

The Medical Device Incident Report Form will be sent to the sponsor by email. If email is unavailable, then the form should be faxed to the sponsor at +41 (22) 710 05 99.

The Trial Manager will be the contact for the receipt of IVD and SAE.

CONFIDENTIAL

##### 8.1.8 Regulatory Reporting Requirements for Medical Device Incidents

The investigator will promptly report all incidents occurring with any IVD provided for use in the study in order for the sponsor to fulfil the legal responsibility to notify appropriate regulatory authorities and other entities about certain safety information relating to medical devices being used in clinical studies.

The investigator, or responsible person according to local requirements (e.g., the head of the medical institution), will comply with the applicable local regulatory requirements relating to the reporting of incidents to the IRB/IEC.

### 9 Statistical Considerations

A detailed statistical analysis plan will be prepared separately and only some key elements are presented here.

#### 9.1 Sample Size Determination

To achieve the targeted precision for accuracy estimates, we estimate that 760 samples be tested for this study, with a final 284 MTB-positive and an additional 316 RIF-resistant samples tested by Xpert MTB/XDR and all reference standards and comparators (assumptions provided in Appendix 4), i.e. a total of 600 specimen results available for analysis. The desired precision for the accuracy estimates was chosen to achieve high confidence in the estimates for Xpert MTB/XDR resistance detection when considering evidence from both the ‘external laboratory validation’ (phase I) and the clinical trial (phase II). Sample size is detailed below in Table 2.

**Table 2. Sample size**

| | | **Clinical trial** (TB samples) | | |
|---|---|---|---|---|
| | | n | Point estimate (95%CI)* | Total width of CI* |
| **INH** | Sensitivity | 395 | 90% (86, 93) | 7% |
| | Specificity | 205 | 98% (95, 99) | 4% |
| **FQs** | Sensitivity | 93 | 90% (82, 95) | 13% |
| | Specificity | 507 | 98% (96, 99) | 3% |

CONFIDENTIAL

| | | | | |
|---|---|---|---|---|
| **AMK** | Sensitivity | 34 | 90%<br>(74, 97) | 23% |
| | Specificity | 566 | 98%<br>(96, 99) | 3% |
| **KAN** | Sensitivity | 71 | 90%<br>(80, 96) | 16% |
| | Specificity | 529 | 98%<br>(96, 99) | 3% |
| **CAP** | Sensitivity | 34 | 90%<br>(74, 97) | 23% |
| | Specificity | 566 | 98%<br>(96, 99) | 3% |

* 95%CI based on Wilson's score method (as recommended by Newcombe et al. as well as CLSI/FDA) using continuity correction.

#### 9.2 Statistical Analysis Plan

The statistical analysis plan (SAP) will be developed and finalized before database lock and will describe the samples to be included in the analyses and procedures for accounting for missing, unused, and spurious data. This section is a summary of the planned statistical analyses of the primary and secondary endpoints.

Primary analyses of outcome data will be done using the composite reference standard. Primary analyses will be focused on estimating accuracy (clinical sensitivity and specificity) for INH and second-line drug resistance detection:

- **Clinical Sensitivity:** proportion positive by composite reference standard (phenotypic DST and NGS) that are detected as positive by index test (Xpert MTB/XDR)
- **Clinical Specificity:** proportion negative by composite reference standard (phenotypic DST and NGS) that are detected as negative by index test (Xpert MTB/XDR)

For simple proportions (sensitivity and specificity), 95% confidence intervals will be computed using the Wilson score method. Primary diagnostic accuracy analyses will be carried out across all study sites. All TB samples that generate both Xpert MTB/XDR and composite reference standard results will contribute to the analysis of the performance characteristics for INH- and second-line resistance detection. If discordances are observed between the Xpert MTB/XDR results from testing sputa and the culture isolate, results from the culture isolate will be considered in these analyses.

Primary analyses will also evaluate assay operational characteristics, which will be captured through direct observation and user-appraisal questionnaires. Primary assessments of Xpert MTB/XDR operational characteristics will evaluate assay characteristics compared to Hain MTBDR*plus*/*sl* assays, including:

CONFIDENTIAL

- Time to first result
- Ease of use
- Failure and invalid rate

These assessments of Xpert and LPA characteristics will be descriptive, and any differences will be further explored by sample type (i.e. smear- and culture-result, patient HIV- and treatment statues, etc.). Assessments of platform operational characteristics will be based upon information gathered during the clinical study, as well as information provided by the manufacturers.

##### 9.2.1 Exclusion Criteria for Diagnostic Performance Analysis

Samples meeting any of the following criteria in the course of the study will be excluded from the primary analyses of Xpert MTB/XDR accuracy:

- no valid result for NGS for relevant gene region or Xpert MTB/XDR,
- smear-positive/culture negative,
- single positive culture with ≤20 colonies (LJ) or time to positivity >28 days (MGIT),
- culture positive but no MTB complex identification available,
- specimens with growth of mycobacteria other than MTB complex only

##### 9.2.2 Definitions for Diagnostic Performance Analysis

| **TEST RESULT** | **DESCRIPTION** |
|---|---|
| Phenotypic Drug*-resistant | Culture-positive and growth for Drug* in conventional DST testing. |
| Phenotypic Drug*-sensitive | Culture-positive and no growth for Drug* in conventional DST testing |
| Genotypic Drug*-resistant | NGS identifies mutations recognized to be associated with resistance (defined based on consultation with WHO prior to analysis) |
| Genotypic Drug*-sensitive | NGS identifies no mutations recognized to be associated with resistance (defined based on consultation with WHO prior to analysis) |
| Composite reference standard Drug*-resistant | If Phenotypic Drug*-sensitive but NGS identifies mutations recognized to be associated with Drug* resistance for the respective gene regions, the composite reference standard will be considered Drug*-resistant.<br>If Phenotypic Drug*-resistant but NGS does not identify mutations recognized to be associated with Drug* resistance for the respective gene regions, the composite reference standard will be considered Drug*-resistant (as mutations will be assumed outside of the region sequenced). |

CONFIDENTIAL

| | |
|---|---|
| Composite reference standard Drug*-sensitive | If Phenotypic Drug*-sensitive <u>and</u> NGS shows either no mutations or only mutations that are not associated with Drug* resistance for the respective gene regions. |

* Drug: INH, ETH, FQ (moxifloxacin and levofloxacin for phenotypic DST), AMK, KAN or CAP

##### 9.2.3 Additional Characteristics Assessed

Secondary analyses will investigate assay diagnostic performance between important subgroups, including: Xpert MTB/XDR direct versus indirect performance (on direct versus cultured samples), performance between sites, by smear result, by gene target, performance compared to Hain MTBDR*plus* and MTBDR*sl* assays, performance by reflex test to Xpert MTB/RIF or Ultra result, and performance by patient HIV and pre-treatment status. Detailed definitions for diagnostic test results are given in Appendix 5: Definitions of Diagnostic Test Results.

##### 9.2.4 Efficacy Analyses

| Endpoint | Statistical Analysis Methods |
|---|---|
| Primary | • Sensitivity<br>• Specificity<br>• Data summaries |
| Secondary | • Chi-squared<br>• Sensitivity<br>• Specificity<br>• Positive Predictive Value<br>• Negative Predictive Value |

#### 9.3 Planned Interim Analyses

Interim analyses for the phase II clinical trial will be led by the Trial Manager and conducted by the Trial Statistician. Preliminary data will be analysed once for every 200 clinical samples tested. Briefly, once 200 clinical samples have been tested by Xpert MTB/XDR both directly on the clinical sample and on the culture isolate and have returned NGS results for all drugs tested, an interim analysis will be conducted. The diagnostic sensitivity and specificity of the Xpert MTB/XDR assay will be estimated against NGS for each drug evaluated, including for important sub-groups as detailed in section 2.

Data will be unmasked for this analysis, and results will serve as an early indicator of Xpert MTB/XDR accuracy in the clinical trial. If performance estimates fall outside of the presented confidence intervals for resistance detection (Table 2), the Data Monitoring Committee (DMC) will review all data for accuracy and consistency.

CONFIDENTIAL

Additionally, the sponsor may conduct a monitoring visit to one or more sites to ensure adherence to SOPs and effective data management.

##### 9.3.1 Data Monitoring Committee

The DMC for Phase II of the Xpert MTB/XDR evaluation will consist of the Head of TB Programme, Head of Clinical & Regulatory Affairs, Trial Manager, Data Manager, and the Data Entry Clerks at each study site. The head of the DMC will be the Data Manager, who will help to train site staff on the use of the database system and perform periodic data checks to ensure that data is collected, managed and reported clearly, securely and accurately throughout the project. The Data Entry Clerks at each site will perform first- and second-data entry and report any issues to the Trial Manager and Data Manager, who will work together with the Data Entry Clerks to resolve any data issues. The Head of Clinical & Regulatory Affairs will serve as an advisor to the DMC.

### 10 Regulatory and Ethical Considerations

#### 10.1 Regulatory and Ethics Approvals

This study will be conducted in accordance with the protocol and with the following:

- Consensus ethical principles derived from international guidelines including the Declaration of Helsinki
- Applicable ICH Good Clinical Practice (GCP) Guidelines
- Applicable laws and regulations

The protocol, protocol amendments, ICF and other relevant documents (eg, advertisements) must be submitted to an IRB/IEC by the investigator and reviewed and approved by the IRB/IEC before the study is initiated.

- Any amendments to the protocol will require IRB/IEC approval before implementation of changes made to the study design, except for changes necessary to eliminate an immediate hazard to study participants.
- The investigator will be responsible for the following:
  - Providing written summaries of the status of the study to the IRB/IEC annually or more frequently in accordance with the requirements, policies, and procedures established by the IRB/IEC
  - Notifying the IRB/IEC of SAEs or other significant safety findings as required by IRB/IEC procedures
  - Providing oversight of the conduct of the study at the site and adherence to requirements of ICH guidelines, the IRB/IEC, the WHO Good Clinical Laboratory Practice (GCLP), European regulation 536/2014 for clinical studies (if applicable), and with applicable national regulations.

CONFIDENTIAL

#### 10.2 Informed Consent Process

The investigator or his/her representative will explain the nature of the study to the participant or his/her legally authorized representative and answer all questions regarding the study.

Participants must be informed that their participation is voluntary. Participants or their legally authorized representative will be required to sign and date a statement of informed consent that meets the requirements of the IRB/IEC or study centre.

There must be evidence that written informed consent was obtained before the participant was enrolled in the study and ample time was given to participant to consent. The date the written consent was obtained (as well as the time, ideally) must be recorded. The authorized person obtaining the informed consent must also sign and date the ICF.

Illiterate participants must provide a thumbprint on the ICF and the ICF signed and dated by an impartial witness.

Participants must be re-consented to the most current version of the ICF(s) during their participation in the study.

A copy of the ICF(s) must be provided to the participant or the participant's legally authorized representative.

#### 10.3 Data Protection

Participants will be assigned a unique identifier by the sponsor. Any participant records or datasets that are transferred to the sponsor will contain the identifier only; participant names or any information which would make the participant identifiable will not be transferred.

The participant must be informed that his/her personal study-related data will be used by the sponsor in accordance with local data protection law. The level of disclosure must also be explained to the participant.

The participant must be informed that his/her medical records may be examined by Clinical Quality Assurance auditors or other authorized personnel appointed by the sponsor, by appropriate IRB/IEC members, and by inspectors from regulatory authorities.

#### 10.4 Other Ethical Considerations

Following completion of study procedures, any remaining samples will be stored temporarily in case further testing for this study is deemed to be necessary. Samples will be labelled with a study identifier and date of collection; the specimens will be linked to results of other mycobacteriology tests, the final study diagnosis for each

CONFIDENTIAL

sample (e.g. drug-resistant, drug-sensitive, indeterminant for each drug compound). Samples will be stored temporarily (up to 2 years) at the sites. Further testing may be required at a Reference Laboratory or specialized research laboratories abroad. An export permit will be obtained accordingly when possible.

In the event that one site does not meet sample processing targets within the study timelines, the study will go forward at the other sites and the investigators will proceed with data submission as per study plan.

### 11 Data Handling and Record Keeping

All participant data relating to the study will be recorded on printed or electronic CRF unless transmitted to the sponsor or designee electronically (e.g., laboratory data). The investigator is responsible for verifying that data entries are accurate and correct by physically or electronically signing the CRF.

The investigator must maintain accurate documentation (source data) that supports the information entered in the CRF.

The investigator must permit study-related monitoring, audits, IRB/IEC review, and regulatory agency inspections and provide direct access to source data documents.

The sponsor or designee is responsible for the data management of this study including quality checking of the data.

Study monitors will perform source data verification to confirm that data entered into the CRF by authorized site personnel are accurate, complete, and verifiable from source documents; that the safety and rights of participants are being protected; and that the study is being conducted in accordance with the currently approved protocol and any other study agreements, ICH GCP, and all applicable regulatory requirements.

Records and documents, including signed ICFs, pertaining to the conduct of this study must be retained by the investigator for 2 years after study completion unless local regulations or institutional policies require a longer retention period. No records may be destroyed during the retention period without the written approval of the sponsor. No records may be transferred to another location or party without written notification to the sponsor.

#### 11.1 Source Data and Source Documents

Source documents provide evidence for the existence of the participant and substantiate the integrity of the data collected. Source documents are filed at the investigator's site and may include:

- Xpert MTB/XDR test results (electronic)
- Xpert MTB/RIF test results (electronic)

CONFIDENTIAL

- Xpert MTB/RIF Ultra test results (electronic)
- Hain MTBDR*plus* and MTBDR*sl* test strips
- NGS results (electronic records)
- Manually-collected data such as lab requisitions, lab registers, and laboratory worksheets (AFB smear log, DST worksheets, QC records, etc.)
- Data generated from additional, automated instruments; e.g., MGIT unloaded positive, negative, and DST reports

Data reported on the CRF or entered in the eCRF that are transcribed from source documents must be consistent with the source documents or the discrepancies must be explained. The investigator may need to request previous medical records or reports (if available), depending on the study.

The investigator/institution should maintain adequate and accurate source documents and trial records that include all pertinent observations on each of the site's trials subjects. Source data should be attributable, legible, contemporaneous, original, accurate and complete. Changes to source data should be traceable, should not obscure the original entry and should be explained if necessary.

#### 11.2 Data Management

For the clinical study, de-identified sample CRF data will consist of participant medical history, clinical examination, and prior laboratory testing results when available. All clinical CRF notes will be transferred to the eCRF by Data Entry Clerks.

All laboratory results (smear, culture, DST, molecular results) will be recorded by laboratory technicians directly on the laboratory CRF. All Xpert testing will generate electronic records of test results. All electronic results will be stored electronically on password-protected computers at the site. Data will also be transcribed into the CRF. LPA testing will generate results on paper test strips. These test strips will be interpreted by the operator and input into the laboratory CRF. All LPA test strips will be retained by the site as source documents. Laboratory CRFs will be transferred from paper CRFs to eCRFs by Data Entry Clerks at the sites and uploaded to the Openclinica study database. The OpenClinica Community v3.13 database is password controlled and restricted. FIND will ultimately be responsible for data management.

Electronic double-data entry must be conducted by two on-site Data Entry Clerks. Data entry training will be provided by FIND, either on site or remotely. Sites will use a password-protected web-based data-entry tool which has the advantage that study monitors will have continuous access to the electronic data. Electronic data will be backed up daily.

Where possible, first data entry will be done as soon as a result becomes available (real-time). Second data entry will only be done by study staff after all sample results

CONFIDENTIAL

(culture, NGS, Xpert) are available. The Principal Investigator of each site will review all data entered for the site's samples.

Details relating to data entry procedures and timelines will be provided to the sites in a data management user manual.

Study monitors will conduct regular checks to see that all paper and electronic records are consistent and accurate. A priority during monitoring visits will be to ensure correctness of electronic data entries in comparison to source data.

Additional details relating to data entry procedures and timelines will be provided to the sites in the data management SOP.

### 12 Quality Management

Quality Management for this study consists of Quality Control activities, training and capacity building provided by FIND (or designee) to the investigational sites and laboratories, as well as the use of Standard Operating Procedures, Work Instructions, Tools and Templates.

Training on the protocol, GCP and the use of the IVDs and laboratory tests will be provided by FIND. A Laboratory Manual which describes all of the sample testing procedures will be provided by FIND prior to the commencement of the study. Training on the EDC system will be provided by FIND Data Management prior to first participant enrolment.

#### 12.1 Quality Control

Quality control should be applied to each stage of data handling to ensure that all data are reliable and have been processed correctly. The investigational site is responsible for performing regular Quality Control checks on the data they generate. Additionally, the sponsor will perform risk based monitoring of this study, and associated Quality Control checks, as described in the Monitoring Plan.

QC will be applied to each stage of data handling to ensure that all data are reliable and have been processed correctly:

- Laboratory staff will be trained in data collection and entry
- All source documentation will be filed and preserved
- All assay results will be recorded onto paper laboratory CRFs immediately following testing
- All data will be double entered from the paper CRF to the eCRF by two on-site staff
- Electronic data will be backed up daily
- The Co-investigator will review all data entered for the site's samples

CONFIDENTIAL

- The Trial Manager and Data Manager will perform periodic data checks to ensure all paper and electronic records are complete and input data are consistent and accurate

#### 12.2 Quality assurance

As part of routine QA, the sponsor (FIND) or designee may conduct an audit of the investigational site. Monitoring visits may include, but are not limited to, review of regulatory files, laboratory reports, and protocol compliance. Study monitors will meet with co-investigators to discuss any problems and actions to be taken and document visit findings and discussions.

FIND will provide training on data collection and entry, implement quality control procedures related to the data entry system and generate data quality control checks that will be run on the database. Any missing data or data anomalies will be communicated to the laboratory for clarification and resolution. The laboratory will provide direct access to all source data/documents for the purpose of monitoring and auditing by the sponsor by local and regulatory authorities, if requested.

#### 12.3 Study and Site Closure

The sponsor designee reserves the right to close the study site or terminate the study at any time for any reason at the sole discretion of the sponsor. Study sites will be closed upon study completion. A study site is considered closed when all required documents and study supplies have been collected and a study-site closure visit has been performed.

The investigator may initiate study-site closure at any time, provided there is reasonable cause and sufficient notice is given in advance of the intended termination.

Reasons for the early closure of a study site by the sponsor or investigator may include but are not limited to:

- Failure of the investigator to comply with the protocol, the requirements of the IRB/IEC or local health authorities, the sponsor's procedures, or GCP guidelines
- Inadequate recruitment of participants by the investigator
- Discontinuation of further study intervention development

### 13 Publication Policy

Authorship will be determined by mutual agreement and in line with International Committee of Medical Journal Editors authorship requirements, as described in the

CONFIDENTIAL

publication policy section of the contractual agreement. Briefly, results of this study will be submitted for publication in peer reviewed journals as soon as feasible upon completion of the study. Appropriate attributions (i.e. co-authorship or acknowledgement regarding the generation of data as the case may be) will be made in all publications related to this study. Authorship determination will be in accordance with the guidelines provided by the International Committee of Medical Journal Editors.

Before a paper or abstract is submitted for publication, the other party shall be provided thirty (30) days to review the proposed publication or disclosure to assure that Proprietary/Confidential Information is protected and to prevent, at its sole discretion, any publication of the party's own Proprietary/Confidential Information.  In the event that Cepheid desires to protect any of the information disclosed in the proposed publication, then FIND agrees to delay publication by an additional 60 days or such other reasonable amount of time as required to allow Cepheid to make any applicable filings.

CONFIDENTIAL

15 Technical Expert Group Meeting Report: critical concentrations for drug susceptibility testing for TB medicines. Geneva: World Health Organization; 2017 (WHO/HTM/TB/2017.22). Licence: CC BY-NC-SA 3.0 IGO.

CONFIDENTIAL

### 15 Appendices

**Appendix 1: Safety Definitions and Reporting**

**Appendix 2: Incident Definition and Reporting**

**Appendix 3: Protocol Amendment Summary Table**

**Appendix 4: Sample Requirement to Achieve Target Sample Size**

**Appendix 5: Definitions of Diagnostic Test Results**

CONFIDENTIAL

### Appendix 1: Safety Definitions and Reporting

| Adverse Event (AE) Definition |
|---|
| • An AE is any unfavorable and unintended sign (including abnormal laboratory finding), symptom or disease temporally associated with the use of an investigational product, whether or not related to the investigational product. |

| Serious Adverse Event (SAE) Definition: |
|---|
| **a. Results in death** |
| **b. Is life-threatening**<br>The term 'life-threatening' in the definition of 'serious' refers to an event in which the participant was at risk of death at the time of the event. It does not refer to an event, which hypothetically might have caused death, if it were more severe. |
| **c. Requires inpatient hospitalization or prolongation of existing hospitalization**<br>In general, hospitalization signifies that the participant has been detained (usually involving at least an overnight stay) at the hospital or emergency ward for observation and/or treatment that would not have been appropriate in the physician's office or outpatient setting. Complications that occur during hospitalization are AEs. If a complication prolongs hospitalization or fulfils any other serious criteria, the event is serious. When in doubt as to whether "hospitalization" occurred or was necessary, the AE should be considered serious.<br>Hospitalization for elective treatment of a pre-existing condition that did not worsen from baseline is not considered an AE. |
| **d. Results in persistent disability/incapacity**<br>• The term disability means a substantial disruption of a person's ability to conduct normal life functions.<br>• This definition is not intended to include experiences of relatively minor medical significance such as uncomplicated headache, nausea, vomiting, diarrhea, influenza, and accidental trauma (e.g., sprained ankle) which may interfere with or prevent everyday life functions but do not constitute a substantial disruption. |
| **e. Is a congenital anomaly/birth defect** |
| **f. Other situations:**<br>• Medical or scientific judgment should be exercised in deciding whether SAE reporting is appropriate in other situations such as important medical events that may not be immediately life-threatening or result in death or hospitalization but may jeopardize the participant or may require medical or surgical intervention to prevent one of the other outcomes listed in the above definition. These events should usually be considered serious.<br>Examples of such events include intensive treatment in an emergency room or at home for allergic bronchospasm or convulsions that do not result in hospitalization, or development of drug dependency or drug abuse. |

CONFIDENTIAL

| SAE Reporting to FIND |
|---|
| • The SAE Report must be sent to the FIND Head of Program and Trial Manager via e-mail, marked High Priority, with a follow up call to ensure receipt.<br>• Initial notification via telephone does not replace the need for the investigator to complete and sign the SAE Report within the designated reporting time frames.<br>• Contacts for SAE reporting can be found on the front of the protocol. |

CONFIDENTIAL

### Appendix 2:  Incident Definition and Reporting

| Medical Device/IVD Incident Definition |
|---|
| • A medical device incident is any malfunction or deterioration in the characteristics and/or performance of a device or IVD as well as any inadequacy in the labeling or the instructions for use which, directly or indirectly, might lead to or might have led to the death of a participant/user/other person or to a serious deterioration in his/her state of health.<br>• Not all incidents lead to death or serious deterioration in health. The nonoccurrence of such a result might have been due to other fortunate circumstances or to the intervention of health care personnel. |

**It is sufficient that:**

- An **incident** associated with a device happened.

  AND

- The **incident** was such that, if it occurred again, might lead to death or a serious deterioration in health.

A serious deterioration in state of health can include any of the following:

- Life-threatening illness
- Permanent impairment of body function or permanent damage to body structure
- Condition necessitating medical or surgical intervention to prevent one of the above
- Fetal distress, fetal death, or any congenital abnormality or birth defects

| Medical Device Incident Documenting |
|---|
| • Any medical device incident occurring during the study will be documented in the participant's medical records, in accordance with the investigator's normal clinical practice, and on the appropriate form of the CRF.<br>• For medical device incidents fulfilling the definition above, complete the SAE Report Form.<br>• It is very important that the investigator provides his/her assessment of causality (relationship to the medical device provided by the sponsor) and describes any corrective or remedial actions taken to prevent recurrence of the incident.<br>• A remedial action is any action other than routine maintenance or servicing of a medical device where such action is necessary to prevent recurrence of an incident. This includes any amendment to the device design to prevent recurrence. |

CONFIDENTIAL

### Appendix 3: Protocol Amendment Summary Table

The Protocol Amendment Summary of Changes Table for the current amendment is located directly before the Table of Contents (TOC).

Amendment [amendment number]: ([date])

#### Overall Rationale for the Amendment

[Rationale]

| Section # and Name | Description of Change | Brief Rationale |
|---|---|---|

CONFIDENTIAL

### Appendix 4: Sample Requirement to Achieve Target Sample Size

| | MTB+ group* | | Additional RIF-R group | | TOTAL |
|---|---|---|---|---|---|
| Samples Tested | 100% | 360 | 100% | 400 | **760** |
| Invalid Xpert Ultra results | 1% | 4 | 1% | 4 | **8** |
| Culture-contaminated | 10% | 36 | 10% | 40 | **76** |
| Molecular-test non-determinant* | 10% | 36 | 10% | 40 | **76** |
| Included** | 79% | 284 | 79% | 316 | **600** |
| INH-resistant | 29% | 82 | 99% | 313 | **395** |
| INH-sensitive | 71% | 202 | 1% | 3 | **205** |
| FQ-resistant | 5% | 14 | 25% | 79 | **93** |
| FQ-sensitive | 95% | 270 | 75% | 237 | **507** |
| AMK/CAP-resistant | 2% | 6 | 9% | 28 | **34** |
| AMK/CAP-sensitive | 98% | 278 | 91% | 288 | **566** |
| KAN-resistant | 1% | 3 | 21.5% | 68 | **71** |
| KAN-sensitive | 99% | 281 | 78.5% | 248 | **529** |

*Assume 23% of TB-positive samples are RIF-R based upon previous studies and data from these regions[11,13].

**We assume 29% INH resistance, 5% FQ resistance and 1-2% second-line injectable (AMK, KAN and CAP) resistance among TB-positive patients based upon previous studies and data from these regions[11,13]. Testing 284 TB-positive specimens (based on Xpert MTB/RIF and Ultra-testing) would then be expected to yield 82 INH-resistant but only 14 FQ-resistant specimens and 3-6 second-line injectable-resistant specimens (limiting factor in sample size calculations). This allows for determination of Xpert MTB/XDR performance for INH resistance detection as a reflex test to any TB-positive result, but is not sufficient for determination for determination of Xpert MTB/XDR performance for second-line resistance detection. To efficiently get to a sufficient number of second-line resistant samples, we will supplement this by testing 316 additional RIF-resistant samples. To account for contaminated cultures and invalid/non-determinant Xpert MTB/RIF or Ultra, NGS and Xpert MTB/XDR results, we would inflate these numbers by 21%, leading to a total of 360 TB-positive and 400 additional RIF-resistant samples, i.e. a total of 760 patient samples tested in this study.

CONFIDENTIAL

### Appendix 5: Definitions of Diagnostic Test Results

| TEST RESULT | DESCRIPTION |
|---|---|
| Smear-positive | ≥ 1 positive FM smear (inclusive of scanty positive smears) using WHO/IUATLD grading. |
| Culture-positive | LJ and/or MGIT culture growth-confirmed MTB complex. |
| Culture-negative | LJ or MGIT have no culture growth after >56 days and >42 days |
| Contaminated culture | LJ: Culture completely overgrown by bacterial or fungal contaminations within 3 weeks (discarded). In case of mixed cultures, isolated MTB colonies transferred to new LJ tube (repeat culture). |
| NGS INH-sensitive | Targeted NGS of *katG, fabG1, ahpC* and *inhA* does NOT detect any mutations associated with INH resistance. |
| NGS INH-resistant | Targeted NGS of *katG, fabG1, ahpC* and *inhA* detects at least one mutation associated with INH resistance. |
| NGS ETH-sensitive | Targeted NGS of *inhA* does NOT detect any mutations associated with INH resistance. |
| NGS ETH-resistant | Targeted NGS of *inhA* detects at least one mutation associated with INH resistance. |
| NGS FQ-sensitive | Targeted NGS of *gyrA* and *gyrB* does NOT detect any mutations associated with FQ resistance. |
| NGS FQ-resistant | Targeted NGS of *gyrA* and *gyrB* detects at least one mutation associated with FQ resistance. |
| NGS AMK-sensitive | Targeted NGS of *rrs* does NOT detect any mutations associated with AMK resistance. |
| NGS AMK-resistant | Targeted NGS of *rrs* detects at least one mutation associated with AMK resistance. |
| NGS KAN-sensitive | Targeted NGS of *rrs* and *eis* does NOT detect any mutations associated with KAN resistance. |
| NGS KAN-resistant | Targeted NGS of *rrs* and *eis* detects at least one mutation associated with KAN resistance. |
| NGS CAP-sensitive | Targeted NGS of *rrs* does NOT detect any mutations associated with CAP resistance. |
| NGS CAP-resistant | Targeted NGS of *rrs* detects at least one mutation associated with CAP resistance. |
| MTBDR*plus* INH-sensitive | No detection of *katG* or *inhA* mutations associated with INH resistance by line probe assay (WT probe hybridization, no MUT probes hybridized). |
| MTBDR*plus* INH-resistant | Detection of *katG* or *inhA* mutations associated with INH resistance by line probe assay (MUT probe hybridization or absence of WT probe hybridization). |

CONFIDENTIAL

| | |
|---|---|
| MTBDR*sl* FQ-sensitive | No detection of *gyrA* or *gyrB* mutations associated with FQ resistance by line probe assay (WT probe hybridization, no MUT probes hybridized). |
| MTBDR*sl* FQ-resistant | Detection of *gyrA* or *gyrB* mutations associated with FQ resistance by line probe assay (MUT probe hybridization or absence of WT probe hybridization). |
| MTBDR*sl* AMK-sensitive | No detection of *rrs* mutations associated with AMK resistance by line probe assay (WT probe hybridization, no MUT probes hybridized). |
| MTBDR*sl* AMK-resistant | Detection of *rrs* mutations associated with AMK resistance by line probe assay (MUT probe hybridization or absence of WT probe hybridization). |
| MTBDR*sl* KAN-sensitive | No detection of *rrs* or *eis* mutations associated with KAN resistance by line probe assay (WT probe hybridization, no MUT probes hybridized). |
| MTBDR*sl* KAN-resistant | Detection of *rrs* or *eis* mutations associated with KAN resistance by line probe assay (MUT probe hybridization or absence of WT probe hybridization). |
| MTBDR*sl* CAP-sensitive | No detection of *rrs* mutations associated with CAP resistance by line probe assay (WT probe hybridization, no MUT probes hybridized). |
| MTBDR*sl* CAP-resistant | Detection of *rrs* mutations associated with CAP resistance by line probe assay (MUT probe hybridization or absence of WT probe hybridization). |
| Xpert MTB/XDR Drug*-sensitive | Valid result with Drug* resistance NOT detected by Xpert MTB/XDR. |
| Xpert MTB/XDR Drug*-resistant | Valid result with Drug* resistance detected by Xpert MTB/XDR. |
| Xpert MTB/XDR Drug*-indeterminant | Valid result with Drug* resistance indeterminant. |
| Xpert MTB/XDR non-determinant** | No valid run result by Xpert MTB/XDR. |

* Drug: INH, ETH, FQ (moxifloxacin and levofloxacin for phenotypic DST), AMK, KAN or CAP
**Xpert non-determinants: result during initial run or after repeat following an invalid result

CONFIDENTIAL
