## Supplement 2 - WGS protocol for "Clinical evaluation of the Xpert MTB/XDR assay for rapid detection of isoniazid, fluoroquinolone, ethionamide and second-line drug resistance: A cross-sectional multicentre diagnostic accuracy study"

### Whole Genome Sequencing Pipeline (Procedures and Analysis)

#### DNA Extraction

After MGIT cultures were flagged as positive and volumes were allocated for other testing and/or storage (i.e. phenotypic DST, Xpert MTB/XDR, MTBDRplus and MTBDRsl), the remaining ~300uL in the MGIT tube was grown an additional 3-5 days. Past this time, the MGIT tube was placed in the fridge at 4 $^{o}$C until DNA extractions were performed.

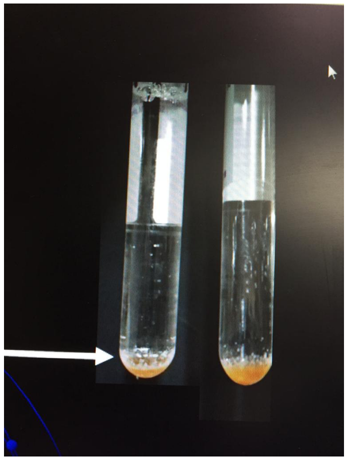

*Figure 1. Cells/clumps at bottom of MGIT tube were transferred in volume for DNA extraction*

DNA extractions were performed using Ultra-Deep Microbiome Prep kit (Molzym GmbH & Co. KG, Bremen, Germany). For the extraction, 300uL of each culture to be extracted was pipetted into the provided, sterile 2mL ST tube, ensuring that any visible cells or clumps were transferred, as per Figure 1. Following this step, steps 3-19 were followed as per the package insert (Protocol 1; page 16-17). 50uL of DNA eluate from the ET tube was sent to Medgenome (Bangalore, India) for whole genome sequencing.

If any DNA aliquots failed QA/QC checks prior to sequencing, these samples were re-cultured by MGIT and if positive were re-extracted, using 1mL of positive MGIT culture, grown an additional 3-5 days following MGIT positivity.  Otherwise, all DNA extraction procedures were followed (starting from step 3 of Protocol 1).

#### Whole Genome Sequencing and Analysis

Whole genome sequencing was conducted using the Illumina Hiseq X Ten platform at Medgenome, Bangalore, India. All details relating to the whole genome sequencing pipeline are detailed within Soundarajan et al.(1) Raw sequencing data was file transferred through FIND-established sFTP. Drug-resistance mutation calls for 11 gene targets of interest (katG, inhA promoter, oxyR-ahpC intergenic region, fabG1, rpoB, gyrA, gyrB, rrs, eis promoter, ethA, tlyA) were reported according to % of resistance alleles at loci of interest. All mutations detected by WGS (see “Relevant resistance mutations,” below) were considered for the analysis, even if the frequency of allele/mutation was a low percentage of the WGS reads and/or disagreed with the phenotypic DST result.

### QC Criteria

**QC criteria**

| Analysis category | QC metrics Description | PASS condition | ALERT condition | FAIL condition |
|---|---|---|---|---|
| Read metrics | Total read >=Q30 (%) | >=85% | <= 80% and <95% | <80% |
| Read metrics | GC content (%) | >50% and <70% | - | <=50% or >=70% |
| Alignment metrics | MTB genome read depth (X) | >= 75 | >=50 and <75 | <50 |
| Alignment metrics | MTB genome coverage (>=1X) | >=85% | - | <85% |
| Alignment metrics | rpoB gene read depth (X) | >= 75 | >=50 and <75 | <50 |
| Alignment metrics | rpoB gene coverage (>=1X) | >=85% | - | <85% |
| Alignment metrics | mpt64 gene read depth (X) | >= 75 | >=50 and <75 | <50 |
| Alignment metrics | mpt64 gene coverage (>=1X) | >=85% | - | <85% |
| Alignment metrics | hsp gene read depth (X) | >= 75 | >=50 and <75 | <50 |
| Alignment metrics | hsp gene coverage (>=1X) | >=85% | - | <85% |

### Results output format

| Non-NGS results | | | Metadata | | | NGS Read metrics | | | | Coverage metrics | | | | | | | | | Lineage | | | Bioinformatic |
|---|---|---|---|---|---|---|---|---|---|---|---|---|---|---|---|---|---|---|---|---|---|---|
| Sample name | Smear score | Xpert result | Library ID | MG sample ID | Sample type | Raw data (Gbps) | No. of reads | Percent of reads (>Q30) | Mean read quality | Genome read-depth(X) | MTB genome coverage (1X) | MTB genome coverage (10X) | rpoB read-depth(X) | rpoB gene coverage | mpt64 read-depth(X) | mpt64 gene coverage | hsp read-depth(X) | hsp gene coverage | Species | Lineage (name, sublineage) | Identity (%) | NGS Bioinformatics Acceptance |
| TB034010001 | #N/A | #N/A | LIB182292 | 398653 | Culture_DNA | 2.19 | 14511084 | 89.31 | 37.54 | 233.71 | 99.61 | 99.36 | 227.14 | 100.00 | 488.61 | 100.00 | 232.90 | 100.00 | M. tuberculosis | Lineage 3 ( N/A ) | 100.0% | PASS |
| TB034010005 | #N/A | #N/A | LIB182293 | 398654 | Culture_DNA | 2.11 | 13965124 | 88.66 | 37.36 | 280.74 | 99.28 | 99.23 | 271.91 | 100.00 | 414.51 | 100.00 | 303.08 | 100.00 | M. tuberculosis | Lineage 2 ( N/A ) | 100.0% | PASS |
| TB034010007 | #N/A | #N/A | LIB182294 | 398655 | Culture_DNA | 1.28 | 8476666 | 88.67 | 37.36 | 147.24 | 99.28 | 99.17 | 147.88 | 100.00 | 221.03 | 100.00 | 152.57 | 100.00 | M. tuberculosis | Lineage 2 ( N/A ) | 99.7% | PASS |

### Relevant resistance mutations

In lieu of a widely accepted mutation manual, resistance mutations were characterized according to a selection of research studies, systematic reviews and meta-analyses of mutations that confer resistance to isoniazid, ethionamide, fluoroquinolone, amikacin, kanamycin and capreomycin.(2–6)

| gene | mutation | drug | resistance | level | note | reference |
|---|---|---|---|---|---|---|
| katG | S315T | INH | R | | | WHO CC report; Miotto et al. |
| katG | S315N | INH | R | | | WHO CC report; Miotto et al. |
| katG | S315G | INH | R | | | WHO CC report |
| katG | S315I | INH | R | | | WHO CC report |
| katG | S315R | INH | R | | | WHO CC report |
| katG | Frameshifts and premature stop codons | INH | R | | | Miotto et al. |
| inhA pt | t-8a | INH | R | low | (in absence of other INH resistance mutations) | WHO CC report |
| inhA pt | t-8c | INH | R | low | (in absence of other INH resistance mutations) | WHO CC report |
| inhA pt | t-8g | INH | R | low | (in absence of other INH resistance mutations) | WHO CC report |
| inhA pt | c-15t | INH | R | low | (in absence of other INH resistance mutations) | WHO CC report; Miotto et al. |
| inhA pt | g-17t | INH | R | low | (in absence of other INH resistance mutations) | WHO CC report |
| fabG1 | L203L | INH | R | | | WHO CC report |
| oxyR-ahpC | (any except g-46a) | INH | R | | | WHO CC report |
| inhA pt | (any) | ETH | R | | | Miotto et al.; Morlock et al. |
| gyrA | G88C | FQ | R | | | WHO CC report; Miotto et al. |
| gyrA | G88A | FQ | R | | | WHO CC report; Miotto et al. |
| gyrA | D89G | FQ | R | | | WHO CC report |
| gyrA | D89N | FQ | R | | | WHO CC report |
| gyrA | A90V | FQ | R | low | (in absence of other FQ resistance mutations) | WHO CC report; Miotto et al. |
| gyrA | S91P | FQ | R | low | (in absence of other FQ resistance mutations) | WHO CC report; Miotto et al. |
| gyrA | D94A | FQ | R | low | (in absence of other FQ resistance mutations) | WHO CC report; Miotto et al. |
| gyrA | D94G | FQ | R | | | WHO CC report; Miotto et al. |
| gyrA | D94H | FQ | R | | | WHO CC report; Miotto et al. |
| gyrA | D94N | FQ | R | | | WHO CC report; Miotto et al. |
| gyrA | D94Y | FQ | R | | | WHO CC report; Miotto et al. |
| gyrB | E501D | FQ | R | | | WHO CC report |
| gyrB | E501V | FQ | R | | | WHO CC report |
| gyrB | A504T | FQ | R | | | WHO CC report |
| gyrB | A504V | FQ | R | | | WHO CC report; Miotto et al. |
| rrs | a1401g | AMK | R | | | WHO CC report; Miotto et al.; Georghiou et al. |
| rrs | g1484t | AMK | R | | | WHO CC report; Miotto et al.; Georghiou et al. |
| eis pt | c-14t | AMK | R | | | WHO CC report |
| rrs | a1401g | KAN | R | | | WHO CC report; Miotto et al.; Georghiou et al. |
| rrs | c1402t | KAN | R | | | WHO CC report; Miotto et al. |
| rrs | c1402a | KAN | R | | | Georghiou et al. |
| rrs | g1484t | KAN | R | | | WHO CC report; Georghiou et al. |
| eis pt | g-10a | KAN | R | | | WHO CC report; Miotto et al. |
| eis pt | g-10c | KAN | R | | | Georghiou et al. |
| eis pt | g-37t | KAN | R | | | WHO CC report; Miotto et al. |
| eis pt | c-12t | KAN | R | | | WHO CC report; Miotto et al. |
| eis pt | c-14t | KAN | R | | | WHO CC report |
| rrs | a1401g | CAP | R | | | WHO CC report; Miotto et al.; Georghiou et al. |
| rrs | c1402t | CAP | R | | | WHO CC report; Miotto et al.; Georghiou et al. |
| rrs | c1402a | CAP | R | | | Georghiou et al. |
| rrs | g1484t | CAP | R | | | WHO CC report; Miotto et al.; Georghiou et al. |
| tlyA | frameshifts and premature stop codons | CAP | R | | | Miotto et al. |
| tlyA | N236K | CAP | R | | | Miotto et al. |
