## Supplement 3 for "Clinical evaluation of the Xpert MTB/XDR assay for rapid detection of isoniazid, fluoroquinolone, ethionamide and second-line drug resistance: A cross-sectional multicentre diagnostic accuracy study"

### Supplement S3.

#### Table 1. Diagnostic test result definitions

| Diagnostic test result | Definition |
|---|---|
| Smear-positive | $\geq$1 positive FM smear (inclusive of scanty positive smears) using WHO/IUATLD grading. |
| Culture-positive | LJ and/or MGIT culture growth-confirmed MTB complex. |
| Culture-negative | LJ and MGIT have no culture growth after >56 days and >42 days, respectively. |
| Contaminated culture | LJ: Culture completely overgrown by bacterial or fungal contaminations within 3 weeks (discarded). In case of mixed cultures, isolated MTB colonies transferred to a new LJ tube (repeat culture).<br>MGIT: Instrument positivity without detection of AFB. |
| WGS INH-sensitive | WGS detects at least one mutation associated with INH resistance. |
| WGS INH-resistant | WGS does NOT detect any mutation associated with INH resistance. |
| WGS ETH-sensitive | WGS detects at least one mutation associated with ETH resistance. |
| WGS ETH-resistant | WGS does NOT detect any mutation associated with ETH resistance. |
| WGS FQ-sensitive | WGS detects at least one mutation associated with FQ resistance. |
| WGS FQ-resistant | WGS does NOT detect any mutation associated with FQ resistance. |
| WGS AMK-sensitive | WGS detects at least one mutation associated with AMK resistance. |
| WGS AMK-resistant | WGS does NOT detect any mutation associated with AMK resistance. |
| WGS KAN-sensitive | WGS detects at least one mutation associated with KAN resistance. |
| WGS KAN-resistant | WGS does NOT detect any mutation associated with KAN resistance. |
| WGS CAP-sensitive | WGS detects at least one mutation associated with CAP resistance. |
| WGS CAP-resistant | WGS does NOT detect any mutation associated with CAP resistance. |
| MTBDRplus INH-sensitive | No detection of katG or inhA mutations associated with INH resistance by line probe assay (WT probe hybridization, no MUT probes hybridized). |
| MTBDRplus INH-resistant | Detection of katG or inhA mutations associated with INH resistance by line probe assay (MUT probe hybridization or absence of WT probe hybridization). |
| MTBDRsl FQ-sensitive | No detection of gyrA or gyrB mutations associated with FQ resistance by line probe assay (WT probe hybridization, no MUT probes hybridized). |
| MTBDRsl FQ-resistant | Detection of gyrA or gyrB mutations associated with FQ resistance by line probe assay (MUT probe hybridization or absence of WT probe hybridization). |
| MTBDRsl AMK-sensitive | No detection of rrs mutations associated with AMK resistance by line probe assay (WT probe hybridization, no MUT probes hybridized). |
| MTBDRsl AMK-resistant | Detection of rrs mutations associated with AMK resistance by line probe assay (MUT probe hybridization or absence of WT probe hybridization). |
| MTBDRsl KAN-sensitive | No detection of rrs or eis mutations associated with KAN resistance by line probe assay (WT probe hybridization, no MUT probes hybridized). |
| MTBDRsl KAN-resistant | Detection of rrs or eis mutations associated with KAN resistance by line probe assay (MUT probe hybridization or absence of WT probe hybridization). |
| MTBDRsl CAP-sensitive | No detection of rrs mutations associated with CAP resistance by line probe assay (WT probe hybridization, no MUT probes hybridized). |
| MTBDRsl CAP-resistant | Detection of rrs mutations associated with CAP resistance by line probe assay (MUT probe hybridization or absence of WT probe hybridization). |
| Xpert MTB/XDR drug*-sensitive | Valid result with drug* resistance NOT detected by Xpert MTB/XDR. |
| Xpert MTB/XDR drug*-resistant | Valid result with drug* resistance detected by Xpert MTB/XDR. |

| | |
|---|---|
| Xpert MTB/XDR drug*-indeterminate | Valid result with drug* resistance indeterminate by Xpert MTB/XDR. |
| Xpert MTB/XDR invalid | No valid result by Xpert MTB/XDR. |
| **Reference standard - test result** | **Definition** |
| Phenotypic drug*-resistant | Culture-positive and growth for drug* in conventional DST testing. |
| Phenotypic drug*-sensitive | Culture-positive and no growth for drug* in conventional DST testing. |
| Genotypic drug*-resistant | WGS identifies mutations recognized to be associated with resistance (defined based on consultation with WHO prior to analysis) |
| Genotypic drug*-sensitive | WGS identifies no mutations recognized to be associated with resistance (defined based on consultation with WHO prior to analysis) |
| Composite reference standard drug*-resistant | If Phenotypic drug*-sensitive but WGS identifies mutations recognized to be associated with drug* resistance for the respective gene regions, the composite reference standard will be considered drug*-resistant.<br>If Phenotypic drug*-resistant but WGS does not identify mutations recognized to be associated with drug* resistance for the respective gene regions, the composite reference standard will be considered drug*-resistant (as unknown resistance mutations will be assumed). |
| Composite reference standard drug*-sensitive | If Phenotypic drug*-sensitive and NGS shows either no mutations or only mutations that are not associated with drug* resistance for the respective gene regions. |

AFB: acid-fast bacilli; AMK: amikacin; CAP: capreomycin; DST: drug susceptibility testing; ETH: ethionamide; IUATLD: International Union Against Tuberculosis and Lung Disease; FM: fluorescence microscopy; FQ: fluoroquinolones; INH: isoniazid; KAN: kanamycin; LJ: Löwenstein–Jensen; MGIT: Mycobacteria Growth Indicator Tube; MTB: *Mycobacterium tuberculosis*; MUT: mutation; NGS: next-generation sequencing; WGS: whole genome sequencing; WHO: World Health Organization; WT: wild type.

*Drug: INH, ETH, FQ (moxifloxacin and levofloxacin for phenotypic DST), AMK, KAN or CAP.
Cases without valid (either negative or positive) smear assessment do not contribute to the calculation of subgroup sensitivity. Evaluation is related to phenotypic reference standard evaluations.

**Table 2. Overall sensitivity and specificity against pDST reference standard**

| All participants | N | TP | FP | FN | TN | Sensitivity % (95% CI) | Specificity % (95% CI) |
|---|---|---|---|---|---|---|---|
| INH resistance detection | 592 | 455 | 6 | 24 | 107 | 95.0 (92.5, 96.7) | 94.7 (88.3, 97.8) |
| ETH resistance detection | 593 | 170 | 13 | 148 | 262 | 53.5 (47.8, 59.0) | 95.3 (91.9, 97.4) |
| FQ resistance detection | 588 | 204 | 20 | 13 | 351 | 94.0 (89.7, 96.6) | 94.6 (91.7, 96.6) |
| AMK resistance detection | 576 | 54 | 8 | 9 | 505 | 85.7 (74.1, 92.9) | 98.4 (96.8, 99.3) |
| KAN resistance detection | 578 | 155 | 32 | 14 | 377 | 91.7 (86.2, 95.2) | 92.2 (89.0, 94.5) |
| CAP resistance detection | 578 | 50 | 3 | 17 | 508 | 74.6 (62.3, 84.1) | 99.4 (98.1, 99.8) |

AMK: amikacin; CAP: capreomycin; CI: confidence interval; ETH: ethionamide; FN: false negatives; FP: false positives; FQ: fluoroquinolones; INH: isoniazid; KAN: kanamycin; N: number; TN: true negatives; TP: true positives.
Cases without valid (either negative or positive) smear assessment do not contribute to the calculation of subgroup sensitivity. Evaluation is related to phenotypic reference standard evaluations.

**Table 3. Sensitivity and specificity by site against pDST as reference standard**

| | N | TP | FP | FN | TN | Sensitivity % (95% CI) | Specificity % (95% CI) |
|---|---|---|---|---|---|---|---|
| **INH-R detection** | | | | | | | |
| All sites | 605 | 464 | 6 | 25 | 110 | 94.9 (92.4, 96.6) | 94.8 (88.6, 97.9) |
| Hinduja | 178 | 143 | 0 | 2 | 33 | 98.6 (94.6, 99.8) | 100.0 (87.0, 100.0) |
| NITRD | 116 | 63 | 5 | 15 | 33 | 80.8 (70.0, 88.5) | 86.8 (71.1, 95.1) |
| Moldova | 230 | 213 | 0 | 3 | 14 | 98.6 (95.7, 99.6) | 100.0 (73.2, 100.0) |
| South Africa | 81 | 45 | 1 | 5 | 30 | 90.0 (77.4, 96.3) | 96.8 (81.5, 99.8) |
| **ETH-R detection** | | | | | | | |
| All sites | 605 | 176 | 13 | 148 | 268 | 54.3 (48.7, 59.8) | 95.4 (92.0, 97.4) |
| Hinduja | 178 | 39 | 2 | 66 | 71 | 37.1 (28.1, 47.2) | 97.3 (89.6, 99.5) |
| NITRD | 116 | 12 | 0 | 19 | 85 | 38.7 (22.4, 57.7) | 100.0 (94.6, 100.0) |
| Moldova | 230 | 101 | 8 | 57 | 64 | 63.9 (55.9, 71.3) | 88.9 (78.7, 94.7) |
| South Africa | 81 | 24 | 3 | 6 | 48 | 80.0 (60.9, 91.6) | 94.1 (82.8, 98.5) |
| **FQ-R detection** | | | | | | | |
| All sites | 604 | 207 | 20 | 15 | 362 | 93.2 (88.9, 96.0) | 94.8 (91.9, 96.7) |
| Hinduja | 178 | 102 | 12 | 2 | 62 | 98.1 (92.5, 99.7) | 83.8 (73.0, 91.0) |
| NITRD | 116 | 38 | 6 | 8 | 64 | 82.6 (68.0, 91.7) | 91.4 (81.6, 96.5) |
| Moldova | 230 | 52 | 2 | 4 | 172 | 92.9 (81.9, 97.7) | 98.9 (95.5, 99.8) |
| South Africa | 80 | 15 | 0 | 1 | 64 | 93.8 (67.7, 99.7) | 100.0 (92.9, 100.0) |
| **AMK-R detection** | | | | | | | |
| All sites | 603 | 56 | 9 | 9 | 529 | 86.2 (74.8, 93.1) | 98.3 (96.7, 99.2) |
| Hinduja | 177 | 19 | 1 | 4 | 153 | 82.6 (60.5, 94.3) | 99.4 (95.9, 100.0) |
| NITRD | 115 | 6 | 0 | 2 | 107 | 75.0 (35.6, 95.5) | 100.0 (95.7, 100.0) |
| Moldova | 230 | 10 | 8 | 2 | 210 | 83.3 (50.9, 97.1) | 96.3 (92.6, 98.3) |
| South Africa | 81 | 21 | 0 | 1 | 59 | 95.5 (75.1, 99.8) | 100.0 (92.4, 100.0) |
| **KAN-R detection** | | | | | | | |
| All sites | 604 | 162 | 34 | 14 | 394 | 92.0 (86.8, 95.4) | 92.1 (89.0, 94.4) |
| Hinduja | 177 | 24 | 14 | 4 | 135 | 85.7 (66.4, 95.3) | 90.6 (84.4, 94.6) |
| NITRD | 116 | 6 | 1 | 2 | 107 | 75.0 (35.6, 95.5) | 99.1 (94.2, 100.0) |
| Moldova | 230 | 111 | 19 | 7 | 93 | 94.1 (87.7, 97.4) | 83.0 (74.5, 89.2) |
| South Africa | 81 | 21 | 0 | 1 | 59 | 95.5 (75.1, 99.8) | 100.0 (92.4, 100.0) |
| **CAP-R Detection** | | | | | | | |
| All sites | 604 | 52 | 3 | 17 | 532 | 75.4 (63.3, 84.6) | 99.4 (98.2, 99.9) |
| Hinduja | 178 | 18 | 1 | 6 | 153 | 75.0 (52.9, 89.4) | 99.4 (95.9, 100.0) |
| NITRD | 116 | 4 | 1 | 2 | 109 | 66.7 (24.1, 94.0) | 99.1 (94.3, 100.0) |
| Moldova | 230 | 10 | 1 | 8 | 211 | 55.6 (31.3, 77.6) | 99.5 (97.0, 100.0) |
| South Africa | 80 | 20 | 0 | 1 | 59 | 95.2 (74.1, 99.8) | 100.0 (92.4, 100.0) |

AMK-R: amikacin-resistant; CAP-R: capreomycin-resistant; CI: confidence interval; ETH-R: ethionamide-resistant; FN: false negatives; FP: false positives; FQ-R: fluoroquinolone-resistant; INH-R: isoniazid-resistant; KAN-R: kanamycin-resistant; N: number; NITRD: National Institute of TB and Respiratory Diseases; TN: true negatives; TP: true positives.

Cases without valid (either negative or positive) smear assessment do not contribute to the calculation of subgroup sensitivity. Presented Xpert MTB/XDR performance primarily relates to “direct sample” test results (missing/invalid “direct” results are replaced by “culture sample” test results, if available). Evaluation is related to phenotypic reference standard evaluations.

**Table 4. Overall sensitivity and specificity by sample type (sputum and culture) against composite reference standard**

| All participants | N | TP | FP | FN | TN | Sensitivity % (95% CI) | Specificity % (95% CI) |
|---|---|---|---|---|---|---|---|
| ***Xpert XDR Direct*** | | | | | | | |
| INH resistance detection | 565 | 460 | 0 | 28 | 77 | 94.3 (91.7, 96.1) | 100.0 (94.1, 100.0) |
| ETH resistance detection | 541 | 178 | 1 | 150 | 212 | 54.3 (48.7, 59.7) | 99.5 (97.0, 100.0) |
| FQ resistance detection | 532 | 222 | 2 | 13 | 295 | 94.5 (90.5, 96.9) | 99.3 (97.3, 99.9) |
| AMK resistance detection | 511 | 60 | 2 | 22 | 427 | 73.2 (62.1, 82.1) | 99.5 (98.1, 99.9) |
| KAN resistance detection | 515 | 181 | 5 | 29 | 300 | 86.2 (80.6, 90.4) | 98.4 (96.0, 99.4) |
| CAP resistance detection | 513 | 53 | 1 | 34 | 425 | 60.9 (49.8, 71.0) | 99.8 (98.5, 100.0) |
| ***Xpert XDR Culture*** | | | | | | | |
| INH resistance detection | 576 | 470 | 0 | 27 | 79 | 94.6 (92.1, 96.3) | 100.0 (94.2, 100.0) |
| ETH resistance detection | 551 | 188 | 4 | 146 | 213 | 56.3 (50.8, 61.7) | 98.2 (95.0, 99.4) |
| FQ resistance detection | 544 | 227 | 0 | 12 | 305 | 95.0 (91.2, 97.3) | 100.0 (98.4, 100.0) |
| AMK resistance detection | 534 | 64 | 1 | 23 | 446 | 73.6 (62.8, 82.2) | 99.8 (98.6, 100.0) |
| KAN resistance detection | 537 | 192 | 3 | 27 | 315 | 87.7 (82.4, 91.6) | 99.1 (97.0, 99.8) |
| CAP resistance detection | 536 | 55 | 0 | 37 | 444 | 59.8 (49.0, 69.7) | 100.0 (98.9, 100.0) |

AMK: amikacin; CAP: capreomycin; CI: confidence interval; ETH: ethionamide; FN: false negatives; FP: false positives; FQ: fluoroquinolones; INH: isoniazid; KAN: kanamycin; N: number; TN: true negatives; TP: true positives.

Cases without valid (either negative or positive) smear assessment do not contribute to the calculation of subgroup sensitivity. Evaluation is related to composite reference standard evaluations.

**Table 5. Overall sensitivity and specificity by smear against composite reference standard**

| All participants | N | TP | FP | FN | TN | Sensitivity % (95% CI) | Sensitivity % Smear Pos (95% CI) - N(all) | Sensitivity % Smear Neg (95% CI) - N(all) | Specificity % (95% CI) |
|---|---|---|---|---|---|---|---|---|---|
| INH resistance detection | 565 | 460 | 0 | 28 | 77 | 94.3 (91.7, 96.1) | 94.3 (91.3, 96.3) 437 | 94.1 (87.1, 97.6) 126 | 100.0 (94.1, 100.0) |
| ETH resistance detection | 541 | 178 | 1 | 150 | 212 | 54.3 (48.7, 59.7) | 54.8 (48.5, 60.9) 417 | 50.8 (38.2, 63.3) 122 | 99.5 (97.0, 100.0) |
| FQ resistance detection | 532 | 222 | 2 | 13 | 295 | 94.5 (90.5, 96.9) | 96.1 (91.7, 98.3) 409 | 89.1 (77.1, 95.5) 121 | 99.3 (97.3, 99.9) |
| AMK resistance detection | 511 | 60 | 2 | 22 | 427 | 73.2 (62.1, 82.1) | 76.1 (63.9, 85.3) 400 | 53.8 (26.1, 79.6) 109 | 99.5 (98.1, 99.9) |
| KAN resistance detection | 515 | 181 | 5 | 29 | 300 | 86.2 (80.6, 90.4) | 87.4 (81.4, 91.8) 403 | 78.8 (60.6, 90.4) 110 | 98.4 (96.0, 99.4) |
| CAP resistance detection | 513 | 53 | 1 | 34 | 425 | 60.9 (49.8, 71.0) | 60.8 (48.7, 71.7) 402 | 54.5 (24.6, 81.9) 109 | 99.8 (98.5, 100.0) |

AMK: amikacin; CAP: capreomycin; CI: confidence interval; ETH: ethionamide; FN: false negatives; FP: false positives; FQ: fluoroquinolones; INH: isoniazid; KAN: kanamycin; N: number; Neg: negative; Pos: positive; TN: true negatives; TP: true positives.

Cases without valid (either negative or positive) smear assessment do not contribute to the calculation of subgroup sensitivity. Evaluation is related to composite reference standard evaluations.

**Table 6. Overall sensitivity and specificity by HIV status against composite reference standard**

| | N | TP | FP | FN | TN | Sensitivity % (95% CI) | Sensitivity % HIV Pos (95% CI) - N(all) | Sensitivity % HIV Neg (95% CI) - N(all) | Specificity % (95% CI) |
|---|---|---|---|---|---|---|---|---|---|
| **INH-R detection** | | | | | | | | | |
| Xpert MTB/XDR Culture | 564 | 461 | 0 | 26 | 77 | 94.7 (92.2, 96.4) | 96.4 (86.4, 99.4) 60 | 93.9 (90.3, 96.2) 340 | 100.0 (94.1, 100.0) |
| Xpert MTB/XDR Direct | 564 | 459 | 0 | 28 | 77 | 94.3 (91.7, 96.1) | 96.4 (86.4, 99.4) 60 | 93.5 (89.9, 96.0) 340 | 100.0 (94.1, 100.0) |
| Diff. [Culture-Direct] | 564 | - | - | - | - | 0.4 (−0.4, 1.5) | 0.0 (−6.5, 6.5) 60 | 0.3 (−1.0, 1.9) 340 | 0.0 (−4.8, 4.8) |
| **ETH-R detection** | | | | | | | | | |
| Xpert MTB/XDR Culture | 541 | 182 | 4 | 146 | 209 | 55.5 (49.9, 60.9) | 79.5 (63.1, 90.1) 53 | 57.1 (49.9, 64.1) 332 | 98.1 (94.9, 99.4) |
| Xpert MTB/XDR Direct | 541 | 178 | 1 | 150 | 212 | 54.3 (48.7, 59.7) | 79.5 (63.1, 90.1) 53 | 55.6 (48.4, 62.6) 332 | 99.5 (97.0, 100.0) |
| Diff. [Culture-Direct] | 541 | - | - | - | - | 1.2 (0.05, 3.1) | 0.0 (−9.0, 9.0) 53 | 1.5 (−0.4, 4.4) 332 | −1.4 (−4.1, 0.4) |
| **FQ-R detection** | | | | | | | | | |
| Xpert MTB/XDR Culture | 530 | 224 | 0 | 10 | 296 | 95.7 (92.0, 97.8) | 100.0 (74.7, 100.0) 45 | 92.6 (86.1, 96.4) 333 | 100.0 (98.4, 100.0) |
| Xpert MTB/XDR Direct | 530 | 222 | 2 | 12 | 294 | 94.9 (91.0, 97.2) | 100.0 (74.7, 100.0) 45 | 91.0 (84.1, 95.2) 333 | 99.3 (97.3, 99.9) |
| Diff. [Culture-Direct] | 530 | - | - | - | - | 0.9 (−1.2, 3.3) | 0.0 (-20.4, 20.4) 45 | 1.6 (−2.3, 6.3) 333 | 0.7 (−0.6, 2.4) |
| **AMK-R detection** | | | | | | | | | |
| Xpert MTB/XDR Culture | 509 | 61 | 1 | 21 | 426 | 74.4 (63.4, 83.1) | 100.0 (74.7, 100.0) 44 | 62.2 (46.5, 75.8) 317 | 99.8 (98.5, 100.0) |
| Xpert MTB/XDR Direct | 509 | 60 | 2 | 22 | 425 | 73.2 (62.1, 82.1) | 100.0 (74.7, 100.0) 44 | 60.0 (44.4, 73.9) 317 | 99.5 (98.1, 99.9) |
| Diff. [Culture-Direct] | 509 | - | - | - | - | 1.2 (−3.3, 6.6) | 0.0 (−20.4, 20.4) 44 | 2.2 (-5.8, 11.6) 317 | 0.2 (−0.7, 1.3) |
| **KAN-R detection** | | | | | | | | | |
| Xpert MTB/XDR Culture | 513 | 183 | 3 | 26 | 301 | 87.6 (82.1, 91.6) | 96.3 (79.1, 99.8) 46 | 86.5 (79.7, 91.4) 319 | 99.0 (96.9, 99.7) |
| Xpert MTB/XDR Direct | 513 | 180 | 5 | 29 | 299 | 86.1 (80.5, 90.4) | 96.3 (79.1, 99.8) 46 | 84.5 (77.4, 89.7) 319 | 98.4 (96.0, 99.4) |
| Diff. [Culture-Direct] | 513 | - | - | - | - | 1.4 (−0.4, 4.1) | 0.0 (−12.5, 12.5) 46 | 2.0 (−0.6, 5.8) 319 | 0.7 (−0.6, 2.4) |
| **CAP-R detection** | | | | | | | | | |
| Xpert MTB/XDR Culture | 512 | 53 | 0 | 34 | 425 | 60.9 (49.8, 71.0) | 88.2 (62.3, 97.9) 44 | 45.7 (31.2, 60.8) 320 | 100.0 (98.9, 100.0) |
| Xpert MTB/XDR Direct | 512 | 53 | 1 | 34 | 424 | 60.9 (49.8, 71.0) | 88.2 (62.3, 97.9) 44 | 43.5 (29.2, 58.8) 320 | 99.8 (98.5, 100.0) |
| Diff. [Culture-Direct] | 512 | - | - | - | - | 0.0 (−5.2, 5.2) | 0.0 (−18.4, 18.4) 44 | 2.2 (−5.7, 11.3) 320 | 0.2 (−0.7, 1.3) |

AMK-R: amikacin-resistant; CAP-R: capreomycin-resistant; CI: confidence interval; Diff: difference; ETH-R: ethionamide-resistant; FN: false negatives; FP: false positives; FQ-R: fluoroquinolone-resistant; INH-R: isoniazid-resistant; KAN-R: kanamycin-resistant; N: number; NC: not calculable; Neg: negative; Pos: positive; TN: true negatives; TP: true positives.

Presented confidence intervals for difference between sensitivities {specificities} are calculated according to Tango. Comparative analysis of "culture vs. direct" testing performances is based on paired sample data. Cases without valid (either negative or positive) HIV status do not contribute to the calculation of HIV subgroup sensitivity.

**Table 7. Overall sensitivity and specificity by pre-treatment status against composite reference standard**

| | N | TP | FP | FN | TN | Sensitivity % (95% CI) | Pre-Treated Sensitivity % (95% CI) - N(all) | Not Pre-Treated Sensitivity % (95% CI) - N(all) | Specificity % (95% CI) | Pre-Treated Specificity % (95% CI) | Not Pre-Treated Specificity % (95% CI) |
|---|---|---|---|---|---|---|---|---|---|---|---|
| **INH-R detection** | | | | | | | | | | | |
| Xpert MTB/XDR Culture | 564 | 461 | 0 | 26 | 77 | 94.7 (92.2, 96.4) | 97.9 (92.0, 99.6) 105 | 93.5 (90.3, 95.8) 418 | 100.0 (94.1, 100.0) | 100.0 (62.9, 100.0) | 100.0 (92.7, 100.0) |
| Xpert MTB/XDR Direct | 564 | 459 | 0 | 28 | 77 | 94.3 (91.7, 96.1) | 95.8 (89.1, 98.7) 105 | 93.5 (90.3, 95.8) 418 | 100.0 (94.1, 100.0) | 100.0 (62.9, 100.0) | 100.0 (92.7, 100.0) |
| Diff. [Culture-Direct] | 564 | - | - | - | - | 0.4 (-0.4, 1.5) | 2.1 (−1.8, 7.3) 105 | 0.0 (−1.1, 1.1) 418 | 0.0 (−4.8, 4.8) | 0.0 (−29.9, 29.9) | 0.0 (−5.8, 5.8) |
| **ETH-R detection** | | | | | | | | | | | |
| Xpert MTB/XDR Culture | 541 | 182 | 4 | 146 | 209 | 55.5 (49.9, 60.9) | 56.1 (43.3, 68.1) 102 | 58.0 (51.4, 64.3) 398 | 98.1 (94.9, 99.4) | 100.0 (88.0, 100.0) | 97.5 (93.3, 99.2) |
| Xpert MTB/XDR Direct | 541 | 178 | 1 | 150 | 212 | 54.3 (48.7, 59.7) | 53.0 (40.4, 65.3) 102 | 57.1 (50.6, 63.5) 398 | 99.5 (97.0, 100.0) | 100.0 (88.0, 100.0) | 99.4 (96.0, 100.0) |
| Diff. [Culture-Direct] | 541 | - | - | - | - | 1.2 (0.05, 3.1) | 3.0 (−2.6, 10.4) 102 | 0.8 (−0.8, 3.0) 398 | −1.4 (−4.1, 0.4) | 0.0 (−9.6, 9.6) | −1.9 (−5.4, 0.5) |
| **FQ-R detection** | | | | | | | | | | | |
| Xpert MTB/XDR Culture | 530 | 224 | 0 | 10 | 296 | 95.7 (92.0, 97.8) | 98.1 (88.8, 99.9) 100 | 94.1 (88.7, 97.1) 391 | 100.0 (98.4, 100.0) | 100.0 (90.4, 100.0) | 100.0 (98.0, 100.0) |
| Xpert MTB/XDR Direct | 530 | 222 | 2 | 12 | 294 | 94.9 (91.0, 97.2) | 98.1 ( 88.8, 99.9) 100 | 92.8 (87.1, 96.2) 391 | 99.3 (97.3, 99.9) | 100.0 (90.4, 100.0) | 99.2 (96.7, 99.9) |
| Diff. [Culture-Direct] | 530 | - | - | - | - | 0.9 (−1.2, 3.3) | 0.0 (−8.2, 8.2) 100 | 1.3 (−1.2, 4.7) 391 | 0.7 (−0.6, 2.4) | 0.0 (−7.7, 7.7) | 0.8 (−0.8, 3.0) |
| **AMK-R detection** | | | | | | | | | | | |
| Xpert MTB/XDR Culture | 509 | 61 | 1 | 21 | 426 | 74.4 (63.4, 83.1) | 76.9 (46.0, 93.8) 94 | 74.2 (61.3, 84.1) 378 | 99.8 (98.5, 100.0) | 100.0 (94.4, 100.0) | 99.7 (98.0, 100.0) |
| Xpert MTB/XDR Direct | 509 | 60 | 2 | 22 | 425 | 73.2 (62.1, 82.1) | 76.9 (46.0, 93.8) 94 | 72.6 (59.6, 82.8) 378 | 99.5 (98.1, 99.9) | 100.0 (94.4, 100.0) | 99.4 (97.5, 99.9) |
| Diff. [Culture-Direct] | 509 | - | - | - | - | 1.2 (-3.3, 6.6) | 0.0 (−22.8, 22.8) 94 | 1.6 (−4.3, 8.6) 378 | 0.2 (−0.7, 1.3) | 0.0 (−4.5, 4.5) | 0.3 (−0.9, 1.8) |
| **KAN-R detection** | | | | | | | | | | | |
| Xpert MTB/XDR Culture | 513 | 183 | 3 | 26 | 301 | 87.6 (82.1, 91.6) | 95.1 (82.2, 99.2) 95 | 86.6 (80.1, 91.3) 381 | 99.0 (96.9, 99.7) | 100.0 (91.7, 100.0) | 98.7 (95.8, 99.7) |
| Xpert MTB/XDR Direct | 513 | 180 | 5 | 29 | 299 | 86.1 (80.5, 90.4) | 92.7 (79.0, 98.1) 95 | 85.4 (78.6, 90.3) 381 | 98.4 (96.0, 99.4) | 100.0 (91.7, 100.0) | 97.8 (94.6, 99.2) |
| Diff. [Culture-Direct] | 513 | - | - | - | - | 1.4 (−0.4, 4.1) | 2.4 (−6.3, 12.6) 95 | 1.3 (−1.1, 4.5) 381 | 0.7 (−0.6, 2.4) | 0.0 (−6.6, 6.6) | 0.9 (−0.8, 3.2) |
| **CAP-R detection** | | | | | | | | | | | |

| | | | | | | | | | | | |
|---|---|---|---|---|---|---|---|---|---|---|---|
| Xpert MTB/XDR Culture | 512 | 53 | 0 | 34 | 425 | 60.9 (49.8, 71.0) | 60.0 (32.9, 82.5) 96 | 61.5 (48.6, 73.1) 379 | 100.0 (98.9, 100.0) | 100.0 (94.4, 100.0) | 100.0 (98.5, 100.0) |
| Xpert MTB/XDR Direct | 512 | 53 | 1 | 34 | 424 | 60.9 (49.8, 71.0) | 60.0 (32.9, 82.5) 96 | 60.0 (47.1, 71.7) 379 | 99.8 (98.5, 100.0) | 100.0 (94.4, 100.0) | 99.7 (98.0, 100.0) |
| Diff. [Culture-Direct] | 512 | - | - | - | - | 0.0 (−5.2, 5.2) | 0.0 (−20.4, 20.4) 96 | 1.5 (−4.1, 8.2) 379 | 0.2 (−0.7, 1.3) | 0.0 (−4.5, 4.5) | 0.3 (−0.9, 1.8) |

AMK-R: amikacin-resistant; CAP-R: capreomycin-resistant; CI: confidence interval; Diff: difference; ETH-R: ethionamide-resistant; FN: false negatives; FP: false positives; FQ-R: fluoroquinolone-resistant; INH-R: isoniazid-resistant; KAN-R: kanamycin-resistant; N: number; NC: not calculable; Neg: negative; Pos: positive; TN: true negatives; TP: true positives.

Presented confidence intervals for difference between sensitivities {specificities} are calculated according to Tango. Comparative analysis of "culture vs. direct" testing performances is based on paired sample data. Cases without information on previous history of TB status do not contribute to the calculation of pre-treatment-subgroup sensitivity. Pre-treated signifies previous treatment failures i.e. these are the cases with documented history of TB where the treatment has not been completed or the patient was NOT considered being cured by completed treatment.

**Table 8. Overall sensitivity and specificity by Xpert reflex test against composite reference standard**

| | N | TP | FP | FN | TN | Sensitivity % (95% CI) | Specificity % (95% CI) |
|---|---|---|---|---|---|---|---|
| **INH-R detection** | | | | | | | |
| *'High/Medium'* | 347 | 280 | 0 | 15 | 52 | 94.9 (91.6, 97.0) | 100.0 (91.4, 100.0) |
| *'Low/Very low'* | 152 | 127 | 0 | 7 | 18 | 94.8 (89.1, 97.7) | 100.0 (78.1, 100.0) |
| Xpert Ultra | 234 | 198 | 0 | 8 | 28 | 96.1 (92.2, 98.2) | 100.0 (85.0, 100.0) |
| Xpert MTB/RIF | 331 | 262 | 0 | 20 | 49 | 92.9 (89.1, 95.5) | 100.0 (90.9, 100.0) |
| 'Difference ('Xpert Ultra' - 'Xpert MTB/RIF') | - | - | - | - | - | 3.2 (−1.2, 7.6) | 0.0 (nc, nc) |
| **ETH-R detection** | | | | | | | |
| *'High/Medium'* | 339 | 101 | 0 | 99 | 139 | 50.5 (43.4, 57.6) | 100.0 (96.6, 100.0) |
| *'Low/Very low'* | 148 | 49 | 1 | 41 | 57 | 54.4 (43.6, 64.9) | 98.3 (89.5, 99.9) |
| Xpert Ultra | 219 | 67 | 0 | 76 | 76 | 46.9 (38.5, 55.4) | 100.0 (94.0, 100.0) |
| Xpert MTB/RIF | 322 | 111 | 1 | 74 | 136 | 60.0 (52.5, 67.0) | 99.3 (95.4, 100.0) |
| 'Difference ('Xpert Ultra' - 'Xpert MTB/RIF') | - | - | - | - | - | −13.1 (−24.6, −1.7) | 0.7 (−1.4, 2.9) |
| **FQ-R detection** | | | | | | | |
| *'High/Medium'* | 338 | 163 | 0 | 5 | 170 | 97.0 (92.8, 98.9) | 100.0 (97.2, 100.0) |
| *'Low/Very low'* | 146 | 42 | 2 | 7 | 95 | 85.7 (72.1, 93.6) | 97.9 (92.0, 99.6) |
| Xpert Ultra | 212 | 125 | 0 | 1 | 86 | 99.2 (95.0, 100.0) | 100.0 (94.7, 100.0) |
| Xpert MTB/RIF | 320 | 97 | 2 | 12 | 209 | 89.0 (81.2, 93.9) | 99.1 (96.3, 99.8) |
| 'Difference ('Xpert Ultra' - 'Xpert MTB/RIF') | - | - | - | - | - | 10.2 (3.3, 17.1) | 0.9 (−1.2, 3.1) |
| **AMK-R detection** | | | | | | | |
| *'High/Medium'* | 330 | 32 | 1 | 18 | 279 | 64.0 (49.1, 76.7) | 99.6 (97.7, 100.0) |
| *'Low/Very low'* | 134 | 8 | 1 | 3 | 122 | 72.7 (39.3, 92.7) | 99.2 (94.9, 100.0) |
| Xpert Ultra | 201 | 36 | 1 | 6 | 158 | 85.7 (70.8, 94.1) | 99.4 (96.0, 100.0) |
| Xpert MTB/RIF | 310 | 24 | 1 | 16 | 269 | 60.0 (43.4, 74.7) | 99.6 (97.6, 100.0) |
| 'Difference ('Xpert Ultra' - 'Xpert MTB/RIF') | - | - | - | - | - | 25.7 (4.8, 46.7) | −0.3 (−1.9, 1.4) |
| **KAN-R detection** | | | | | | | |
| *'High/Medium'* | 331 | 99 | 1 | 20 | 211 | 83.2 (75.0, 89.2) | 99.5 (97.0, 100.0) |
| *'Low/Very low'* | 137 | 58 | 4 | 7 | 68 | 89.2 (78.5, 95.2) | 94.4 (85.7, 98.2) |
| Xpert Ultra | 203 | 61 | 1 | 9 | 132 | 87.1 (76.5, 93.6) | 99.2 (95.3, 100.0) |
| Xpert MTB/RIF | 312 | 120 | 4 | 20 | 168 | 85.7 (78.6, 90.8) | 97.7 (93.8, 99.3) |
| 'Difference ('Xpert Ultra' - 'Xpert MTB/RIF') | - | - | - | - | - | 1.4 (−9.4, 12.3) | 1.6 (−1.8, 4.9) |
| **CAP-R detection** | | | | | | | |
| *'High/Medium'* | 331 | 27 | 0 | 26 | 278 | 50.9 (37.0, 64.7) | 100.0 (98.3, 100.0) |
| *'Low/Very low'* | 135 | 6 | 1 | 7 | 121 | 46.2 (20.4, 73.9) | 99.2 (94.8, 100.0) |
| Xpert Ultra | 202 | 35 | 0 | 9 | 158 | 79.5 (64.2, 89.7) | 100.0 (97.0, 100.0) |

| | | | | | | | |
|---|---|---|---|---|---|---|---|
| Xpert MTB/RIF | 311 | 18 | 1 | 25 | 267 | 41.9 (27.4, 57.8) | 99.6 (97.6, 100.0) |
| 'Difference ('Xpert Ultra' - 'Xpert MTB/RIF') | - | - | - | - | - | 37.7 (16.4, 58.9) | 0.4 (−0.7, 1.5) |

AMK-R: amikacin-resistant; CAP-R: capreomycin-resistant; CI: confidence interval; ETH-R: ethionamide-resistant; FN: false negatives; FP: false positives; FQ-R: fluoroquinolone-resistant; INH-R: isoniazid-resistant; KAN-R: kanamycin-resistant; N: number; NC: not calculable; Neg: negative; Pos: positive; TN: true negatives; TP: true positives.

Presented measures of diagnostic accuracy refer to the performance of Xpert MTB/XDR direct sample test results by pre-test method. Cases without semi-quantitative pre-test result being either of “Very low”, “Low”, “Medium” or “High” are considered in the referring overall pre-test method groups, but are not referred to any of the distinct pre-test result sub-categories. Presented “Differences” estimates were rounded after calculation therefore deviations may occur in the last digit compared with the intuitive difference when looking at (also rounded) group-percentages. Presented differences in confidence intervals denoted as “nc” refer to cases without variation in observed data (i.e. in both methods, binary proportion estimates are consistently 100% {or 0%}).

**Table 9. Overall sensitivity and specificity by Xpert reflex test against composite reference standard**

| INH-R DETECTION | | | | | | | | | | | |
|---|---|---|---|---|---|---|---|---|---|---|---|
| **Participant ID** | **Site** | **MGIT Culture Result** | **LJ Culture Result** | **Smear Result** | **Speciation Result** | **MGIT INH-R Result** | **WGS INH-R Result** | **Xpert MTB/XDR INH-R Result** | **Hain MTBDRplus INH-R Result** | **TB History** | **WGS Resistance Mutation** |
| TB034010069 | Hinduja | Pos | Pos | Pos | MTB Complex | Resistant | R | Not det | Not Det. | No | katG E607* |
| TB034010135 | Hinduja | Pos | Pos | Pos | MTB Complex | Resistant | S | Not det | Not Det. | Don't Know | no |
| TB034020016 | NITRD | Pos | Pos | Pos | MTB Complex | Sensitive | R | Not det | Not Det. | No | oxyR-ahpC g-45a |
| TB034020033 | NITRD | Pos | Pos | Neg | MTB Complex | Resistant | S | Not det | Not Det. | No | no |
| TB034020044 | NITRD | Pos | Pos | Pos | MTB Complex | Resistant | S | Not det | Not Det. | No | no |
| TB034020054 | NITRD | Pos | Pos | Pos | MTB Complex | Resistant | R | Not det | Not Det. | No | oxyR-ahpC t-34a; t-35a |
| TB034020058 | NITRD | Pos | Pos | Pos | MTB Complex | Sensitive | R | Not det | Not Det. | No | katG S315T |
| TB034020069 | NITRD | Pos | Neg | Neg | MTB Complex | Resistant | R | Not det | | No | katG S315T |
| TB034020082 | NITRD | Pos | Pos | Pos | MTB Complex | Resistant | S | Not det | Not Det. | No | no |
| TB034020087 | NITRD | Pos | Pos | Neg | MTB Complex | Resistant | R | Not det | Not Det. | No | katG W135* |
| TB034020092 | NITRD | Pos | Neg | Pos | MTB Complex | Resistant | S | Not det | Not Det. | No | no |
| TB034020093 | NITRD | Pos | Neg | Neg | MTB Complex | Resistant | S | Not det | Not Det. | Yes | no |
| TB034020114 | NITRD | Pos | Neg | Pos | MTB Complex | Resistant | R | Not det | Not Det. | Yes | oxyR-ahpC t-34a |
| TB034020118 | NITRD | Pos | Neg | Pos | MTB Complex | Resistant | R | Not det | Not Det. | No | oxyR-ahpC t-34a |
| TB034020122 | NITRD | Pos | Neg | Neg | MTB Complex | Resistant | S | Not det | Not Det. | No | no |
| TB034020125 | NITRD | Pos | Neg | Neg | MTB Complex | Resistant | | Not det | Not Det. | No | |
| TB034020143 | NITRD | Pos | Pos | Pos | MTB Complex | Resistant | S | Not det | Not Det. | Yes | no |
| TB034020152 | NITRD | Pos | Pos | Pos | MTB Complex | Resistant | R | Not det | Not Det. | Yes | oxyR-ahpC t-35g |
| TB034030033 | Moldova | Pos | Pos | Pos | MTB Complex | Resistant | R | Not det | Not Det. | Yes | oxyR-ahpC g-32a |
| TB034030035 | Moldova | Pos | Pos | Pos | MTB Complex | Resistant | R | Not det | Detected | Yes | katG S315T; inhA c-15t; oxyR-ahpC g-32a |
| TB034030085 | Moldova | Pos | Pos | Pos | MTB Complex | Sensitive | R | Not det | Not Det. | Yes | katG S315T |

| | | | | | | | | | | | |
|---|---|---|---|---|---|---|---|---|---|---|---|
| TB034030107 | Moldova | Pos | Pos | Pos | MTB Complex | Resistant | R | Not det | Not Det. | No | katG S315T; inhA c-15t |
| TB034040044 | S Africa | Pos | Pos | Pos | MTB Complex | Resistant | S | Not det | Not Det. | No | no |
| TB034040056 | S Africa | Pos | Pos | Pos | MTB Complex | Resistant | | Not det | Not Det. | No | |
| TB034040058 | S Africa | Pos | Neg | Pos | MTB Complex | Resistant | | Not det | Not Det. | Yes | |
| TB034040067 | S Africa | Pos | Pos | Pos | MTB Complex | Resistant | R | Not det | Detected | Yes | inhA c-15t |
| TB034040088 | S Africa | Pos | Neg | Pos | MTB Complex | Resistant | R | Not det | Detected | Yes | inhA t-8c |
| TB034040092 | S Africa | Pos | Neg | Pos | MTB Complex | Sensitive | R | Not det | Not Det. | Yes | katG S315T |
| **ETH-R DETECTION** | | | | | | | | | | | |
| **Participant ID** | **Site** | **MGIT Culture Result** | **LJ Culture Result** | **Smear Result** | **Speciation Result** | **MGIT ETH-R Result** | **WGS ETH-R Result** | **Xpert MTB/XDR ETH-R Result** | **Hain MTBDRplus ETH-R Result** | **TB History** | **WGS Resistance Mutation** |
| TB034010006 | Hinduja | Pos | Pos | Pos | MTB Complex | Resistant | S | Not det | Not Det. | Yes | no |
| TB034010008 | Hinduja | Pos | Neg | Neg | MTB Complex | Resistant | S | Not det | Not Det. | Don't Know | no |
| TB034010014 | Hinduja | Pos | Pos | Neg | MTB Complex | Resistant | S | Not det | Not Det. | No | no |
| TB034010016 | Hinduja | Pos | Contaminated | Pos | MTB Complex | Resistant | | Not det | Not Det. | No | |
| TB034010019 | Hinduja | Pos | Pos | Pos | MTB Complex | Resistant | S | Not det | Not Det. | No | no |
| TB034010021 | Hinduja | Pos | Neg | Neg | MTB Complex | Resistant | S | Not det | Not Det. | Yes | no |
| TB034010022 | Hinduja | Pos | Neg | Neg | MTB Complex | Resistant | S | Not det | Not Det. | Yes | no |
| TB034010026 | Hinduja | Pos | Pos | Pos | MTB Complex | Resistant | S | Not det | Not Det. | Yes | no |
| TB034010027 | Hinduja | Pos | Pos | Pos | MTB Complex | Resistant | S | Not det | Not Det. | No | no |
| TB034010031 | Hinduja | Pos | Pos | Pos | MTB Complex | Resistant | S | Not det | Not Det. | Yes | no |
| TB034010033 | Hinduja | Pos | Neg | Pos | MTB Complex | Resistant | S | Not det | Not Det. | No | no |
| TB034010034 | Hinduja | Pos | Neg | Pos | MTB Complex | Resistant | S | Not det | Not Det. | No | no |
| TB034010037 | Hinduja | Pos | Pos | Pos | MTB Complex | Resistant | S | Not det | Not Det. | Yes | no |
| TB034010042 | Hinduja | Pos | Pos | Pos | MTB Complex | Resistant | S | Not det | Not Det. | Yes | no |
| TB034010047 | Hinduja | Pos | Pos | Pos | MTB Complex | Resistant | S | Not det | Not Det. | Don't Know | no |

| | | | | | | | | | | | |
|---|---|---|---|---|---|---|---|---|---|---|---|
| TB034010051 | Hinduja | Pos | Pos | Neg | MTB Complex | Resistant | S | Not det | Not Det. | No | no |
| TB034010055 | Hinduja | Pos | Pos | Pos | MTB Complex | Resistant | S | Not det | Not Det. | Don't Know | no |
| TB034010063 | Hinduja | Pos | Pos | Pos | MTB Complex | Resistant | S | Not det | Not Det. | No | no |
| TB034010065 | Hinduja | Pos | Pos | Pos | MTB Complex | Resistant | S | Not det | Not Det. | Don't Know | no |
| TB034010071 | Hinduja | Pos | Pos | Pos | MTB Complex | Resistant | S | Not det | Not Det. | Yes | no |
| TB034010074 | Hinduja | Pos | Pos | Pos | MTB Complex | Resistant | S | Not det | Not Det. | Yes | no |
| TB034010078 | Hinduja | Pos | Pos | Pos | MTB Complex | Resistant | S | Not det | Not Det. | Yes | no |
| TB034010079 | Hinduja | Pos | Pos | Pos | MTB Complex | Resistant | S | Not det | Not Det. | No | no |
| TB034010080 | Hinduja | Pos | Neg | Pos | MTB Complex | Resistant | | Not det | Not Det. | Don't Know | |
| TB034010087 | Hinduja | Pos | Pos | Pos | MTB Complex | Resistant | S | Not det | Not Det. | Yes | no |
| TB034010088 | Hinduja | Pos | Pos | Pos | MTB Complex | Resistant | S | Not det | Not Det. | Yes | no |
| TB034010090 | Hinduja | Pos | Pos | Neg | MTB Complex | Resistant | S | Not det | Not Det. | No | no |
| TB034010093 | Hinduja | Pos | Pos | Pos | MTB Complex | Resistant | S | Not det | Not Det. | Yes | no |
| TB034010097 | Hinduja | Pos | Pos | Pos | MTB Complex | Resistant | S | Not det | Not Det. | Yes | no |
| TB034010101 | Hinduja | Pos | Neg | Pos | MTB Complex | Resistant | S | Not det | Not Det. | Yes | no |
| TB034010108 | Hinduja | Pos | Pos | Pos | MTB Complex | Resistant | S | Not det | Not Det. | Don't Know | no |
| TB034010110 | Hinduja | Pos | Neg | Pos | MTB Complex | Resistant | S | Not det | Not Det. | Yes | no |
| TB034010114 | Hinduja | Pos | Pos | Neg | MTB Complex | Resistant | S | Not det | Not Det. | Yes | no |
| TB034010115 | Hinduja | Pos | Pos | Neg | MTB Complex | Resistant | S | Not det | Not Det. | Yes | no |
| TB034010120 | Hinduja | Pos | Pos | Pos | MTB Complex | Resistant | S | Not det | Not Det. | Don't Know | no |
| TB034010125 | Hinduja | Pos | Pos | Pos | MTB Complex | Resistant | S | Not det | Not Det. | Yes | no |
| TB034010126 | Hinduja | Pos | Pos | Pos | MTB Complex | Resistant | S | Not det | Not Det. | Yes | no |
| TB034010132 | Hinduja | Pos | Pos | Pos | MTB Complex | Resistant | S | Not det | Not Det. | No | no |
| TB034010133 | Hinduja | Pos | Pos | Pos | MTB Complex | Resistant | S | Not det | Not Det. | No | no |
| TB034010134 | Hinduja | Pos | Pos | Pos | MTB Complex | Resistant | S | Not det | Not Det. | No | no |
| TB034010140 | Hinduja | Pos | Neg | Pos | MTB Complex | Resistant | | Not det | Not Det. | Yes | |
| TB034010142 | Hinduja | Pos | Pos | Neg | MTB Complex | Resistant | S | Not det | Not Det. | Don't Know | no |
| TB034010144 | Hinduja | Pos | Neg | Pos | MTB Complex | Resistant | | Not det | Not Det. | Don't Know | |

| | | | | | | | | | | | |
|---|---|---|---|---|---|---|---|---|---|---|---|
| TB034010147 | Hinduja | Pos | Contaminated | Pos | MTB Complex | Resistant | S | Not det | Not Det. | Don't Know | no |
| TB034010149 | Hinduja | Pos | Contaminated | Pos | MTB Complex | Resistant | | Not det | Not Det. | Don't Know | |
| TB034010151 | Hinduja | Pos | Pos | Pos | MTB Complex | Resistant | S | Not det | Not Det. | Don't Know | no |
| TB034010153 | Hinduja | Pos | Neg | Pos | MTB Complex | Resistant | S | Not det | Not Det. | Don't Know | no |
| TB034010154 | Hinduja | Pos | Pos | Pos | MTB Complex | Resistant | S | Not det | Not Det. | No | no |
| TB034010155 | Hinduja | Pos | Neg | Neg | MTB Complex | Resistant | S | Not det | Not Det. | No | no |
| TB034010159 | Hinduja | Pos | Pos | Pos | MTB Complex | Resistant | S | Not det | Not Det. | Don't Know | no |
| TB034010160 | Hinduja | Pos | Pos | Pos | MTB Complex | Resistant | S | Not det | Not Det. | Don't Know | no |
| TB034010163 | Hinduja | Pos | Pos | Pos | MTB Complex | Resistant | S | Not det | Not Det. | No | no |
| TB034010166 | Hinduja | Pos | Neg | Pos | MTB Complex | Resistant | S | Not det | Not Det. | Yes | no |
| TB034010167 | Hinduja | Pos | Pos | Pos | MTB Complex | Resistant | S | Not det | Not Det. | Don't Know | no |
| TB034010172 | Hinduja | Pos | Neg | Pos | MTB Complex | Resistant | | Not det | Not Det. | Don't Know | |
| TB034010173 | Hinduja | Pos | Neg | Neg | MTB Complex | Resistant | S | Not det | Not Det. | Yes | no |
| TB034010176 | Hinduja | Pos | Pos | Pos | MTB Complex | Resistant | S | Not det | Not Det. | No | no |
| TB034010180 | Hinduja | Pos | Neg | Neg | MTB Complex | Resistant | S | Not det | Not Det. | No | no |
| TB034010181 | Hinduja | Pos | Pos | Pos | MTB Complex | Resistant | S | Not det | Not Det. | No | no |
| TB034010182 | Hinduja | Pos | Pos | Pos | MTB Complex | Resistant | S | Not det | Not Det. | Yes | no |
| TB034010187 | Hinduja | Pos | Pos | Pos | MTB Complex | Resistant | S | Not det | Not Det. | No | no |
| TB034010188 | Hinduja | Pos | Pos | Pos | MTB Complex | Resistant | S | Not det | Not Det. | Yes | no |
| TB034010190 | Hinduja | Pos | Pos | Neg | MTB Complex | Resistant | S | Not det | Not Det. | No | no |
| TB034010197 | Hinduja | Pos | Pos | Pos | MTB Complex | Resistant | S | Not det | Not Det. | No | no |
| TB034010198 | Hinduja | Pos | Contaminated | Pos | MTB Complex | Resistant | S | Not det | Not Det. | Yes | no |
| TB034010203 | Hinduja | Pos | Contaminated | Pos | MTB Complex | Resistant | S | Not det | Not Det. | Yes | no |
| TB034020021 | NITRD | Pos | Pos | Pos | MTB Complex | Resistant | S | Not det | Not Det. | Yes | no |
| TB034020032 | NITRD | Pos | Pos | Neg | MTB Complex | Sensitive | R | Not det | Not Det. | No | inhA c-15t |

| | | | | | | | | | | | |
|---|---|---|---|---|---|---|---|---|---|---|---|
| TB034020041 | NITRD | Pos | Neg | Neg | MTB Complex | Resistant | S | Not det | Not Det. | No | no |
| TB034020044 | NITRD | Pos | Pos | Pos | MTB Complex | Resistant | S | Not det | Not Det. | No | no |
| TB034020046 | NITRD | Pos | Pos | Pos | MTB Complex | Resistant | S | Not det | Not Det. | No | no |
| TB034020068 | NITRD | Pos | Pos | Neg | MTB Complex | Resistant | S | Not det | Not Det. | No | no |
| TB034020076 | NITRD | Pos | Pos | Pos | MTB Complex | Resistant | S | Not det | Not Det. | Yes | no |
| TB034020081 | NITRD | Pos | Pos | Neg | MTB Complex | Resistant | S | Not det | Not Det. | Yes | no |
| TB034020086 | NITRD | Pos | Pos | Pos | MTB Complex | Resistant | S | Not det | Not Det. | No | no |
| TB034020092 | NITRD | Pos | Neg | Pos | MTB Complex | Resistant | S | Not det | Not Det. | No | no |
| TB034020106 | NITRD | Pos | Pos | Neg | MTB Complex | Resistant | S | Not det | Not Det. | No | no |
| TB034020108 | NITRD | Pos | Pos | Pos | MTB Complex | Resistant | S | Not det | Not Det. | Yes | no |
| TB034020113 | NITRD | Pos | Neg | Neg | MTB Complex | Resistant | S | Not det | Not Det. | No | no |
| TB034020120 | NITRD | Pos | Pos | Pos | MTB Complex | Resistant | S | Not det | Not Det. | No | no |
| TB034020125 | NITRD | Pos | Neg | Neg | MTB Complex | Resistant | | Not det | Not Det. | No | |
| TB034020127 | NITRD | Pos | Neg | Neg | MTB Complex | Resistant | S | Not det | Not Det. | No | no |
| TB034020132 | NITRD | Pos | Pos | Pos | MTB Complex | Resistant | R | Not det | Detected | No | inhA t-8c |
| TB034020139 | NITRD | Pos | Neg | Pos | MTB Complex | Resistant | S | Not det | Not Det. | No | no |
| TB034020140 | NITRD | Pos | Neg | Pos | MTB Complex | Resistant | S | Not det | Not Det. | No | no |
| TB034020148 | NITRD | Pos | Pos | Neg | MTB Complex | Resistant | | Not det | Detected | No | |
| TB034030001 | Moldova | Pos | Pos | Neg | MTB Complex | Resistant | S | Not det | Not Det. | No | no |
| TB034030007 | Moldova | Pos | Pos | Pos | MTB Complex | Resistant | S | Not det | Not Det. | No | no |
| TB034030008 | Moldova | Pos | Pos | Pos | MTB Complex | Resistant | S | Not det | Not Det. | No | no |
| TB034030011 | Moldova | Pos | Pos | Neg | MTB Complex | Resistant | S | Not det | Not Det. | Yes | no |
| TB034030016 | Moldova | Pos | Pos | Pos | MTB Complex | Resistant | S | Not det | Not Det. | Yes | no |
| TB034030018 | Moldova | Pos | Neg | Neg | MTB Complex | Resistant | S | Not det | Not Det. | No | no |
| TB034030020 | Moldova | Pos | Pos | Neg | MTB Complex | Resistant | S | Not det | Not Det. | No | no |
| TB034030023 | Moldova | Pos | Pos | Pos | MTB Complex | Resistant | S | Not det | Not Det. | Yes | no |
| TB034030027 | Moldova | Pos | Pos | Pos | MTB Complex | Resistant | S | Not det | Not Det. | Yes | no |
| TB034030035 | Moldova | Pos | Pos | Pos | MTB Complex | Resistant | R | Not det | Detected | Yes | inhA c-15t |

| | | | | | | | | | | | |
|---|---|---|---|---|---|---|---|---|---|---|---|
| TB034030038 | Moldova | Pos | Pos | Pos | MTB Complex | Resistant | S | Not det | Not Det. | Yes | no |
| TB034030042 | Moldova | Pos | Pos | Pos | MTB Complex | Resistant | S | Not det | Not Det. | Yes | no |
| TB034030043 | Moldova | Pos | Pos | Pos | MTB Complex | Resistant | S | Not det | Not Det. | Yes | no |
| TB034030060 | Moldova | Pos | Neg | Pos | MTB Complex | Resistant | S | Not det | Not Det. | No | no |
| TB034030071 | Moldova | Pos | Pos | Pos | MTB Complex | Resistant | S | Not det | Not Det. | No | no |
| TB034030075 | Moldova | Pos | Pos | Pos | MTB Complex | Resistant | S | Not det | Not Det. | Yes | no |
| TB034030076 | Moldova | Pos | Pos | Pos | MTB Complex | Resistant | S | Not det | Not Det. | No | no |
| TB034030080 | Moldova | Pos | Pos | Pos | MTB Complex | Resistant | S | Not det | Not Det. | No | no |
| TB034030096 | Moldova | Pos | Pos | Neg | MTB Complex | Resistant | S | Not det | Not Det. | No | no |
| TB034030100 | Moldova | Pos | Pos | Pos | MTB Complex | Resistant | S | Not det | Not Det. | No | no |
| TB034030105 | Moldova | Pos | Pos | Pos | MTB Complex | Resistant | S | Not det | Not Det. | Yes | no |
| TB034030107 | Moldova | Pos | Pos | Pos | MTB Complex | Resistant | R | Not det | Not Det. | No | inhA c-15t |
| TB034030114 | Moldova | Pos | Pos | Pos | MTB Complex | Resistant | S | Not det | Not Det. | Yes | no |
| TB034030117 | Moldova | Pos | Pos | Pos | MTB Complex | Resistant | S | Not det | Not Det. | No | no |
| TB034030123 | Moldova | Pos | Pos | Pos | MTB Complex | Resistant | S | Not det | Not Det. | No | no |
| TB034030129 | Moldova | Pos | Pos | Pos | MTB Complex | Resistant | S | Not det | Detected | Yes | no |
| TB034030130 | Moldova | Pos | Pos | Pos | MTB Complex | Resistant | S | Not det | Detected | Yes | no |
| TB034030135 | Moldova | Pos | Pos | Pos | MTB Complex | Resistant | S | Not det | Not Det. | No | no |
| TB034030142 | Moldova | Pos | Pos | Pos | MTB Complex | Resistant | S | Not det | Not Det. | No | no |
| TB034030147 | Moldova | Pos | Pos | Neg | MTB Complex | Resistant | S | Not det | Not Det. | Yes | no |
| TB034030148 | Moldova | Pos | Pos | Neg | MTB Complex | Resistant | S | Not det | Not Det. | Yes | no |
| TB034030154 | Moldova | Pos | Pos | Neg | MTB Complex | Resistant | S | Not det | Not Det. | Yes | no |
| TB034030155 | Moldova | Pos | Pos | Pos | MTB Complex | Resistant | S | Not det | Not Det. | No | no |
| TB034030157 | Moldova | Pos | Pos | Pos | MTB Complex | Resistant | S | Not det | Not Det. | Yes | no |
| TB034030161 | Moldova | Pos | Pos | Pos | MTB Complex | Resistant | S | Not det | Not Det. | No | no |
| TB034030164 | Moldova | Pos | Pos | Pos | MTB Complex | Resistant | S | Not det | Not Det. | Yes | no |
| TB034030167 | Moldova | Pos | Pos | Pos | MTB Complex | Resistant | S | Not det | Not Det. | No | no |
| TB034030168 | Moldova | Pos | Pos | Pos | MTB Complex | Resistant | S | Not det | Not Det. | No | no |

| | | | | | | | | | | | |
|---|---|---|---|---|---|---|---|---|---|---|---|
| TB034030171 | Moldova | Pos | Pos | Pos | MTB Complex | Resistant | S | Not det | Not Det. | No | no |
| TB034030179 | Moldova | Pos | Pos | Neg | MTB Complex | Sensitive | S | Det | Detected | Yes | no |
| TB034030184 | Moldova | Pos | Pos | Pos | MTB Complex | Resistant | S | Not det | Not Det. | Yes | no |
| TB034030188 | Moldova | Pos | Pos | Pos | MTB Complex | Resistant | S | Not det | Detected | No | no |
| TB034030189 | Moldova | Pos | Pos | Pos | MTB Complex | Resistant | S | Not det | Not Det. | Yes | no |
| TB034030194 | Moldova | Pos | Pos | Neg | MTB Complex | Resistant | S | Not det | Not Det. | No | no |
| TB034030204 | Moldova | Pos | Pos | Pos | MTB Complex | Resistant | S | Not det | Not Det. | Yes | no |
| TB034030210 | Moldova | Pos | Pos | Pos | MTB Complex | Resistant | S | Not det | Not Det. | No | no |
| TB034030212 | Moldova | Pos | Pos | Pos | MTB Complex | Resistant | S | Not det | Not Det. | No | no |
| TB034030214 | Moldova | Pos | Pos | Pos | MTB Complex | Resistant | S | Not det | Not Det. | No | no |
| TB034030215 | Moldova | Pos | Pos | Pos | MTB Complex | Resistant | S | Not det | Not Det. | Yes | no |
| TB034030216 | Moldova | Pos | Pos | Pos | MTB Complex | Resistant | S | Not det | Not Det. | No | no |
| TB034030217 | Moldova | Pos | Pos | Pos | MTB Complex | Resistant | S | Not det | Not Det. | Yes | no |
| TB034030218 | Moldova | Pos | Pos | Pos | MTB Complex | Resistant | S | Not det | Not Det. | Yes | no |
| TB034030222 | Moldova | Pos | Pos | Pos | MTB Complex | Resistant | S | Not det | Not Det. | No | no |
| TB034030228 | Moldova | Pos | Pos | Pos | MTB Complex | Resistant | S | Not det | Not Det. | No | no |
| TB034030235 | Moldova | Pos | Pos | Pos | MTB Complex | Resistant | S | Not det | Not Det. | Yes | no |
| TB034030236 | Moldova | Pos | Pos | Pos | MTB Complex | Resistant | S | Not det | Not Det. | Yes | no |
| TB034030244 | Moldova | Pos | Pos | Pos | MTB Complex | Resistant | S | Not det | Not Det. | Yes | no |
| TB034030245 | Moldova | Pos | Pos | Neg | MTB Complex | Resistant | S | Not det | Not Det. | Yes | no |
| TB034040012 | S Africa | Pos | Pos | Pos | MTB Complex | Resistant | | Not det | Not Det. | Yes | |
| TB034040036 | S Africa | Pos | Neg | Pos | MTB Complex | Resistant | | Not det | Not Det. | Yes | |
| TB034040044 | S Africa | Pos | Pos | Pos | MTB Complex | Resistant | S | Not det | Not Det. | No | no |
| TB034040048 | S Africa | Pos | Pos | Pos | MTB Complex | Resistant | S | Not det | Not Det. | No | no |
| TB034040067 | S Africa | Pos | Pos | Pos | MTB Complex | Resistant | R | Not det | Detected | Yes | inhA c-15t |
| TB034040088 | S Africa | Pos | Neg | Pos | MTB Complex | Sensitive | R | Not det | Detected | Yes | inhA t-8c |
| TB034040095 | S Africa | Pos | Neg | Pos | MTB Complex | Resistant | | Not det | Not Det. | Yes | |
| **FQ-R DETECTION** | | | | | | | | | | | |

| Participant ID | Site | MGIT Culture Result | LJ Culture Result | Smear Result | Speciation Result | MGIT FQ-R Result | WGS FQ-R Result | Xpert MTB/XDR FQ-R Result | Hain MTBDRsl FQ-R Result | TB History | WGS Resistance Mutation |
|---|---|---|---|---|---|---|---|---|---|---|---|
| TB034010005 | Hinduja | Pos | Pos | Neg | MTB Complex | LVX:Resistant\|MXF:Resistant | R | Not det | Detected | Don't Know | gyrA D94N |
| TB034020006 | NITRD | Pos | Contaminated | Neg | MTB Complex | LVX:Resistant\|MXF:Sensitive | S | Not det | Not Det. | No | no |
| TB034020041 | NITRD | Pos | Neg | Neg | MTB Complex | LVX:Resistant\|MXF:Resistant | S | Not det | Detected | No | no |
| TB034020066 | NITRD | Pos | Pos | Pos | MTB Complex | LVX:Resistant\|MXF:Resistant | R | Not det | Not Det. | No | gyrA D94G |
| TB034020069 | NITRD | Pos | Neg | Neg | MTB Complex | LVX:Resistant\|MXF:Resistant | R | Not det | Not Det. | No | gyrB E501D |
| TB034020118 | NITRD | Pos | Neg | Pos | MTB Complex | LVX:Resistant\|MXF:Resistant | R | Not det | Detected | No | gyrA D94G |
| TB034020148 | NITRD | Pos | Pos | Neg | MTB Complex | LVX:Resistant\|MXF:Resistant | | Not det | Detected | No | |
| TB034020151 | NITRD | Pos | Pos | Pos | MTB Complex | LVX:Resistant\|MXF:Resistant | R | Not det | Detected | Yes | gyrA A90V; D94G |
| TB034030079 | Moldova | Pos | Pos | Pos | MTB Complex | LVX:Sensitive\|MXF:Resistant | S | Not det | Not Det. | No | no |
| TB034030107 | Moldova | Pos | Pos | Pos | MTB Complex | LVX:Resistant\|MXF:Resistant | R | Not det | Not Det. | No | gyrA A90V |
| TB034030127 | Moldova | Pos | Neg | Neg | MTB Complex | LVX:Sensitive\|MXF:Sensitive | S | Low-level R det | Not Det. | Yes | no |
| TB034030131 | Moldova | Pos | Pos | Pos | MTB Complex | LVX:Sensitive\|MXF:Sensitive | S | Low-level R det | Detected | No | no |
| TB034030165 | Moldova | Pos | Neg | Neg | MTB Complex | LVX:Resistant\|MXF:Sensitive | S | Not det | Detected | No | no |
| TB034030176 | Moldova | Pos | Pos | Pos | MTB Complex | LVX:Resistant\|MXF:Resistant | S | Not det | Detected | No | no |

| | | | | | | | | | | | |
|---|---|---|---|---|---|---|---|---|---|---|---|
| TB034040044 | S Africa | Pos | Pos | Pos | MTB Complex | LVX:Resistant\|MXF:Resistant | S | Not det | Not Det. | No | no |
| **AMK-R DETECTION** | | | | | | | | | | | |
| **Participant ID** | **Site** | **MGIT Culture Result** | **LJ Culture Result** | **Smear Result** | **Speciation Result** | **MGIT AMK-R Result** | **WGS AMK-R Result** | **Xpert MTB/XDR AMK-R Result** | **Hain MTBDRsl AMK-R Result** | **TB History** | **WGS Resistance Mutation** |
| TB034010076 | Hinduja | Pos | Pos | Pos | MTB Complex | Sensitive | S | Det | Not Det. | Yes | no |
| TB034010088 | Hinduja | Pos | Pos | Pos | MTB Complex | Resistant | R | Not det | Detected | Yes | rrs a1401g |
| TB034010116 | Hinduja | Pos | Contaminated | Neg | MTB Complex | Resistant | S | Not det | Not Det. | Don't Know | no |
| TB034010125 | Hinduja | Pos | Pos | Pos | MTB Complex | Resistant | R | Not det | Detected | Yes | rrs a1401g |
| TB034010185 | Hinduja | Pos | Pos | Pos | MTB Complex | Resistant | R | Not det | Detected | Don't Know | rrs a1401g |
| TB034020040 | NITRD | Pos | Neg | Pos | MTB Complex | Sensitive | R | Not det | Not Det. | No | rrs g1484t |
| TB034020041 | NITRD | Pos | Neg | Neg | MTB Complex | Sensitive | R | Not det | Not Det. | No | rrs g1484t |
| TB034020048 | NITRD | Pos | Pos | Pos | MTB Complex | Sensitive | R | Not det | Not Det. | No | rrs g1484t |
| TB034020052 | NITRD | Pos | Pos | Neg | MTB Complex | Sensitive | R | Not det | Not Det. | No | rrs g1484t |
| TB034020055 | NITRD | Pos | Pos | Pos | MTB Complex | Sensitive | R | Not det | Not Det. | No | rrs g1484t |
| TB034020063 | NITRD | Pos | Pos | Pos | MTB Complex | Sensitive | R | Not det | Not Det. | No | rrs g1484t |
| TB034020070 | NITRD | Pos | Pos | Pos | MTB Complex | Sensitive | R | Not det | Not Det. | No | rrs g1484t |
| TB034020089 | NITRD | Pos | Neg | Pos | MTB Complex | Sensitive | R | Not det | Not Det. | No | rrs g1484t |
| TB034020091 | NITRD | Pos | Pos | Pos | MTB Complex | Sensitive | R | Not det | Not Det. | No | rrs g1484t |
| TB034020119 | NITRD | Pos | Neg | Neg | MTB Complex | Contaminated | R | Not det | Not Det. | No | rrs g1484t |
| TB034020121 | NITRD | Pos | Neg | Pos | MTB Complex | Sensitive | R | Not det | Not Det. | No | rrs g1484t |
| TB034020122 | NITRD | Pos | Neg | Neg | MTB Complex | Resistant | S | Not det | Not Det. | No | no |
| TB034020148 | NITRD | Pos | Pos | Neg | MTB Complex | Resistant | | Not det | Detected | No | |
| TB034030022 | Moldova | Pos | Pos | Neg | MTB Complex | Sensitive | S | Det | Not Det. | No | no |
| TB034030034 | Moldova | Pos | Pos | Pos | MTB Complex | Sensitive | R | Not det | Not Det. | No | rrs g1484t |

| | | | | | | | | | | | |
|---|---|---|---|---|---|---|---|---|---|---|---|
| TB034030041 | Moldova | Pos | Pos | Pos | MTB Complex | Sensitive | R | Not det | Not Det. | Yes | rrs g1484t |
| TB034030104 | Moldova | Pos | Pos | Pos | MTB Complex | Resistant | R | Not det | Detected | Yes | rrs g1484t |
| TB034030224 | Moldova | Pos | Pos | Pos | MTB Complex | Resistant | R | Not det | Detected | No | rrs a1401g |
| TB034040044 | S Africa | Pos | Pos | Pos | MTB Complex | Resistant | S | Not det | Not Det. | No | no |
| **KAN-R DETECTION** | | | | | | | | | | | |
| **Participant ID** | **Site** | **MGIT Culture Result** | **LJ Culture Result** | **Smear Result** | **Speciation Result** | **MGIT KAN-R Result** | **WGS KAN-R Result** | **Xpert MTB/XDR KAN-R Result** | **Hain MTBDRsl KAN-R Result** | **TB History** | **WGS Result** |
| TB034010042 | Hinduja | Pos | Pos | Pos | MTB Complex | Sensitive | S | Det | Not Det. | Yes | no |
| TB034010088 | Hinduja | Pos | Pos | Pos | MTB Complex | Resistant | R | Not det | Detected | Yes | rrs a1401g |
| TB034010116 | Hinduja | Pos | Contaminated | Neg | MTB Complex | Resistant | S | Not det | Not Det. | Don't Know | no |
| TB034010125 | Hinduja | Pos | Pos | Pos | MTB Complex | Resistant | R | Not det | Detected | Yes | rrs a1401g; eis g-10a |
| TB034010153 | Hinduja | Pos | Neg | Pos | MTB Complex | Sensitive | R | Not det | Not Det. | Don't Know | rrs c1402a |
| TB034010185 | Hinduja | Pos | Pos | Pos | MTB Complex | Resistant | R | Not det | Detected | Don't Know | rrs a1401g |
| TB034020040 | NITRD | Pos | Neg | Pos | MTB Complex | Sensitive | R | Not det | Not Det. | No | rrs c1402a; g1484t |
| TB034020041 | NITRD | Pos | Neg | Neg | MTB Complex | Sensitive | R | Not det | Not Det. | No | rrs c1402a; g1484t |
| TB034020048 | NITRD | Pos | Pos | Pos | MTB Complex | Sensitive | R | Not det | Not Det. | No | rrs c1402a; g1484t |
| TB034020052 | NITRD | Pos | Pos | Neg | MTB Complex | Sensitive | R | Not det | Not Det. | No | rrs c1402a; g1484t |
| TB034020055 | NITRD | Pos | Pos | Pos | MTB Complex | Sensitive | R | Not det | Not Det. | No | rrs c1402a; g1484t |
| TB034020063 | NITRD | Pos | Pos | Pos | MTB Complex | Sensitive | R | Not det | Not Det. | No | rrs c1402a; g1484t |
| TB034020070 | NITRD | Pos | Pos | Pos | MTB Complex | Sensitive | R | Not det | Not Det. | No | rrs c1402a; g1484t |
| TB034020089 | NITRD | Pos | Neg | Pos | MTB Complex | Sensitive | R | Not det | Not Det. | No | rrs c1402a; g1484t |
| TB034020091 | NITRD | Pos | Pos | Pos | MTB Complex | Sensitive | R | Not det | Not Det. | No | rrs c1402a; g1484t |
| TB034020119 | NITRD | Pos | Neg | Neg | MTB Complex | Contaminated | R | Not det | Not Det. | No | rrs c1402a; g1484t |
| TB034020121 | NITRD | Pos | Neg | Pos | MTB Complex | Sensitive | R | Not det | Not Det. | No | rrs c1402a; g1484t |

| | | | | | | | | | | | |
|---|---|---|---|---|---|---|---|---|---|---|---|
| TB034020122 | NITRD | Pos | Neg | Neg | MTB Complex | Resistant | S | Not det | Not Det. | No | no |
| TB034020148 | NITRD | Pos | Pos | Neg | MTB Complex | Resistant | | Not det | Detected | No | |
| TB034030005 | Moldova | Pos | Pos | Neg | MTB Complex | Sensitive | S | Det | Not Det. | No | no |
| TB034030035 | Moldova | Pos | Pos | Pos | MTB Complex | Resistant | R | Not det | Not Det. | Yes | rrs c1402a; eis c-12t |
| TB034030049 | Moldova | Pos | Pos | Pos | MTB Complex | Sensitive | S | Det | Not Det. | No | no |
| TB034030085 | Moldova | Pos | Pos | Pos | MTB Complex | Sensitive | R | Not det | Not Det. | Yes | eis g-37t |
| TB034030107 | Moldova | Pos | Pos | Pos | MTB Complex | Resistant | R | Not det | Not Det. | No | eis c-12t |
| TB034030127 | Moldova | Pos | Neg | Neg | MTB Complex | Sensitive | S | Det | Not Det. | Yes | no |
| TB034030165 | Moldova | Pos | Neg | Neg | MTB Complex | Resistant | S | Not det | Not Det. | No | no |
| TB034030176 | Moldova | Pos | Pos | Pos | MTB Complex | Resistant | S | Not det | Not Det. | No | no |
| TB034030179 | Moldova | Pos | Pos | Neg | MTB Complex | Sensitive | S | Det | Not Det. | Yes | no |
| TB034030185 | Moldova | Pos | Pos | Pos | MTB Complex | Resistant | S | Not det | Not Det. | No | no |
| TB034030188 | Moldova | Pos | Pos | Pos | MTB Complex | Resistant | S | Not det | Not Det. | No | no |
| TB034030223 | Moldova | Pos | Pos | Pos | MTB Complex | Resistant | S | Not det | Not Det. | No | no |
| TB034030224 | Moldova | Pos | Pos | Pos | MTB Complex | Sensitive | R | Not det | Detected | No | rrs a1401g |
| TB034040044 | S Africa | Pos | Pos | Pos | MTB Complex | Resistant | S | Not det | Not Det. | No | no |
| TB034040092 | S Africa | Pos | Neg | Pos | MTB Complex | Sensitive | R | Not det | Not Det. | Yes | eis g-10a |
| **CAP-R DETECTION** | | | | | | | | | | | |
| **Participant ID** | **Site** | **MGIT Culture Result** | **LJ Culture Result** | **Smear Result** | **Speciation Result** | **MGIT CAP-R Result** | **WGS CAP-R Result** | **Xpert MTB/XDR CAP-R Result** | **Hain MTBDRsl CAP-R Result** | **TB History** | **WGS Result** |
| TB034010049 | Hinduja | Pos | Pos | Pos | MTB Complex | Resistant | S | Not det | Not Det. | No | no |
| TB034010063 | Hinduja | Pos | Pos | Pos | MTB Complex | Resistant | S | Not det | Not Det. | No | no |
| TB034010088 | Hinduja | Pos | Pos | Pos | MTB Complex | Resistant | R | Not det | Detected | Yes | rrs a1401g |
| TB034010125 | Hinduja | Pos | Pos | Pos | MTB Complex | Resistant | R | Not det | Detected | Yes | rrs a1401g |
| TB034010137 | Hinduja | Pos | Pos | Pos | MTB Complex | Resistant | S | Not det | Not Det. | Yes | no |

| | | | | | | | | | | | |
|---|---|---|---|---|---|---|---|---|---|---|---|
| TB034010153 | Hinduja | Pos | Neg | Pos | MTB Complex | Sensitive | R | Not det | Not Det. | Don't Know | rrs c1402a |
| TB034010185 | Hinduja | Pos | Pos | Pos | MTB Complex | Resistant | R | Not det | Detected | Don't Know | rrs a1401g |
| TB034020040 | NITRD | Pos | Neg | Pos | MTB Complex | Sensitive | R | Not det | Not Det. | No | rrs c1402a; g1484t |
| TB034020041 | NITRD | Pos | Neg | Neg | MTB Complex | Sensitive | R | Not det | Not Det. | No | rrs c1402a; g1484t |
| TB034020048 | NITRD | Pos | Pos | Pos | MTB Complex | Sensitive | R | Not det | Not Det. | No | rrs c1402a; g1484t |
| TB034020052 | NITRD | Pos | Pos | Neg | MTB Complex | Sensitive | R | Not det | Not Det. | No | rrs c1402a; g1484t |
| TB034020055 | NITRD | Pos | Pos | Pos | MTB Complex | Sensitive | R | Not det | Not Det. | No | rrs c1402a; g1484t |
| TB034020063 | NITRD | Pos | Pos | Pos | MTB Complex | Sensitive | R | Not det | Not Det. | No | rrs c1402a; g1484t |
| TB034020070 | NITRD | Pos | Pos | Pos | MTB Complex | Sensitive | R | Not det | Not Det. | No | rrs c1402a; g1484t |
| TB034020089 | NITRD | Pos | Neg | Pos | MTB Complex | Sensitive | R | Not det | Not Det. | No | rrs c1402a; g1484t |
| TB034020091 | NITRD | Pos | Pos | Pos | MTB Complex | Sensitive | R | Not det | Not Det. | No | rrs c1402a; g1484t |
| TB034020119 | NITRD | Pos | Neg | Neg | MTB Complex | Contaminated | R | Not det | Not Det. | No | rrs c1402a; g1484t |
| TB034020121 | NITRD | Pos | Neg | Pos | MTB Complex | Sensitive | R | Not det | Not Det. | No | rrs c1402a; g1484t |
| TB034020122 | NITRD | Pos | Neg | Neg | MTB Complex | Resistant | S | Not det | Not Det. | No | no |
| TB034020148 | NITRD | Pos | Pos | Neg | MTB Complex | Resistant | | Not det | Detected | No | |
| TB034030010 | Moldova | Pos | Pos | Pos | MTB Complex | Sensitive | R | Not det | Not Det. | Yes | rrs c1402a |
| TB034030022 | Moldova | Pos | Pos | Neg | MTB Complex | Sensitive | S | Det | Not Det. | No | no |
| TB034030034 | Moldova | Pos | Pos | Pos | MTB Complex | Sensitive | R | Not det | Not Det. | No | rrs c1402a; g1484t |
| TB034030035 | Moldova | Pos | Pos | Pos | MTB Complex | Sensitive | R | Not det | Not Det. | Yes | rrs c1402a |
| TB034030041 | Moldova | Pos | Pos | Pos | MTB Complex | Sensitive | R | Not det | Not Det. | Yes | rrs c1402a; g1484t |
| TB034030065 | Moldova | Pos | Pos | Pos | MTB Complex | Resistant | S | Not det | Not Det. | No | no |
| TB034030070 | Moldova | Pos | Pos | Pos | MTB Complex | Sensitive | R | Not det | Not Det. | No | rrs c1402a |
| TB034030072 | Moldova | Pos | Pos | Pos | MTB Complex | Resistant | R | Not det | Not Det. | Yes | tlyA S159* |
| TB034030073 | Moldova | Pos | Pos | Pos | MTB Complex | Resistant | S | Not det | Not Det. | No | no |
| TB034030075 | Moldova | Pos | Pos | Pos | MTB Complex | Resistant | S | Not det | Not Det. | Yes | no |
| TB034030104 | Moldova | Pos | Pos | Pos | MTB Complex | Resistant | R | Not det | Detected | Yes | rrs g1484t |
| TB034030138 | Moldova | Pos | Pos | Pos | MTB Complex | Resistant | S | Not det | Not Det. | Yes | no |
| TB034030176 | Moldova | Pos | Pos | Pos | MTB Complex | Resistant | S | Not det | Not Det. | No | no |

| | | | | | | | | | | | |
|---|---|---|---|---|---|---|---|---|---|---|---|
| TB034030224 | Moldova | Pos | Pos | Pos | MTB Complex | Resistant | R | Not det | Detected | No | rrs a1401g |
| TB034040044 | S Africa | Pos | Pos | Pos | MTB Complex | Resistant | S | Not det | Not Det. | No | no |

AMK-R: amikacin-resistant; CAP-R: capreomycin-resistant; CI: confidence interval; Det: detected; ETH-R: ethionamide-resistant; FQ-R: fluoroquinolone-resistant; ID: identification; INH-R: isoniazid-resistant; KAN-R: kanamycin-resistant; ; LJ: Löwenstein–Jensen; MGIT: Mycobacteria Growth Indicator Tube; MTB: *Mycobacterium tuberculosis*; Neg: negative; Pos: positive; R: resistant; S: sensitive; TB: tuberculosis; WGS: whole genome sequencing.
